# Determinants of functional burden pleiotropy and gene dosage responses across human traits

**DOI:** 10.1101/2025.02.25.25322833

**Authors:** Sayeh Kazem, Kuldeep Kumar, Jane Yang, Florian Benitiere, Guillaume Huguet, Josephine Mollon, Thomas Renne, Laura M. Schultz, Emma E.M. Knowles, Worrawat Engchuan, Omar Shanta, Bhooma Thiruvahindrapuram, Jeffrey R. MacDonald, Celia M. T. Greenwood, Stephen W. Scherer, Laura Almasy, Jonathan Sebat, David C. Glahn, Guillaume Dumas, Sébastien Jacquemont

## Abstract

Pleiotropic and monotonic effects of gene dosage are central to understanding comorbidities in developmental pediatric and psychiatric disorders, yet the underlying biological processes are unknown. We developed Functional Burden analysis (FunBurd) to investigate the association of all protein-coding copy-number-variants (CNVs), genome-wide, with 43 complex traits in ∼500,000 UK-Biobank participants. We tested CNV associations disrupting 172 tissue or cell-type gene-sets. We observed associations for all traits and replicated these associations in the All of Us cohort. Functional burden pleiotropy, defined as the number of traits significantly associated with a gene set, was correlated with genetic constraint and was higher for brain compared to non-brain functions, even after normalizing for genetic constraint. Levels of pleiotropy, measured by burden correlation, were similar in deletions and loss-of-function SNVs, and higher compared to common variants and duplications. Most gene dosage responses were non-monotonic, with deletions and duplications showing same-direction effects, and the proportions of monotonic responses decreased with genetic constraint. Consistent with this paucity of monotonicity, associations between functional gene sets and traits were observed for either deletions or duplications, but rarely both. In addition, deletion and duplication effect sizes were negatively correlated, demonstrating that associations between traits and functional gene sets are variant-specific. Our results highlight the key role of genetic constraint and brain-specific mechanisms in shaping monotonicity and pleiotropy, providing a mechanistic basis for the whole-body multimorbidity observed in neurodevelopmental and psychiatric conditions.

## MAIN

Deciphering functions at the cell and tissue level mediating genetic associations with complex traits and conditions is a central question in human genetics^1–4^. It can offer insights into the molecular understanding of causal processes of diseases^1^. Genome-wide association studies (GWAS) have identified thousands of common variants associated with complex traits^1,2^. While functional genomic studies have shed light on the tissues and cell types disrupted by these variants and how they impact gene expression levels, it remains challenging to link common non-coding variants to genes and corresponding functions^1,4–8^.

In contrast, copy number variants (CNVs) – deletions or duplications of DNA segments – invariably lead to a large decrease or increase in gene expression when a gene is fully encompassed in a deletion or a duplication, respectively ^9–12^. Therefore, they provide direct insight into the effects of large changes in gene expression on complex traits. However, studies have been limited to a few of the most recurrent CNVs ^11,13,14^ due to statistical power. As a result, our knowledge of the CNV architecture of complex traits and biological functions that are sensitive to gene dosage remains limited.

CNVs are commonly screened in individuals referred to developmental pediatric clinics for complex neurodevelopmental disorders and congenital malformations^15–17^. Since most clinical studies have focused on pediatric populations, there is limited understanding of the medical issues that may arise later in life. Coding CNVs are associated with broad pleiotropic effects^10– 12,18,19^, yet it remains unclear whether these pleiotropic effects are driven by the multigenic nature of CNVs or by particular genes (and corresponding functions) encompassed in the CNVs. Due to statistical power, previous studies have repeatedly analyzed a small set of the recurrent CNVs^17,19,20^, which collectively affect only approximately 2% of the coding genome^21^. As a result, our understanding of gene functions sensitive to gene dosage is highly biased.

Previous studies have shown that measures of evolutionary/genetic constraint (such as LOEUF score^22^) explain a significant proportion of the effects of coding CNVs on cognitive and behavioral traits^23–26^. It is, however, unclear if the biological function of genes can inform the effects of CNVs beyond constraint metrics. In particular, it is unknown if the effects of CNVs on traits that are not under genetic constraint can be explained by gene function.

Our overarching aim is to systematically characterize the CNV architecture of complex traits (both brain and non-brain) across different levels of biological function, encompassing tissue and cell types, and compare it with the common variant architecture. To study all rare coding CNVs (beyond those sufficiently frequent to establish locus-level association), we implemented the functional burden association test (FunBurd)^26^, which aggregated CNVs that disrupt genes with similar functions, defined through shared expression patterns across tissues and cell types (**Fig. 1**).

**Fig. 1:**
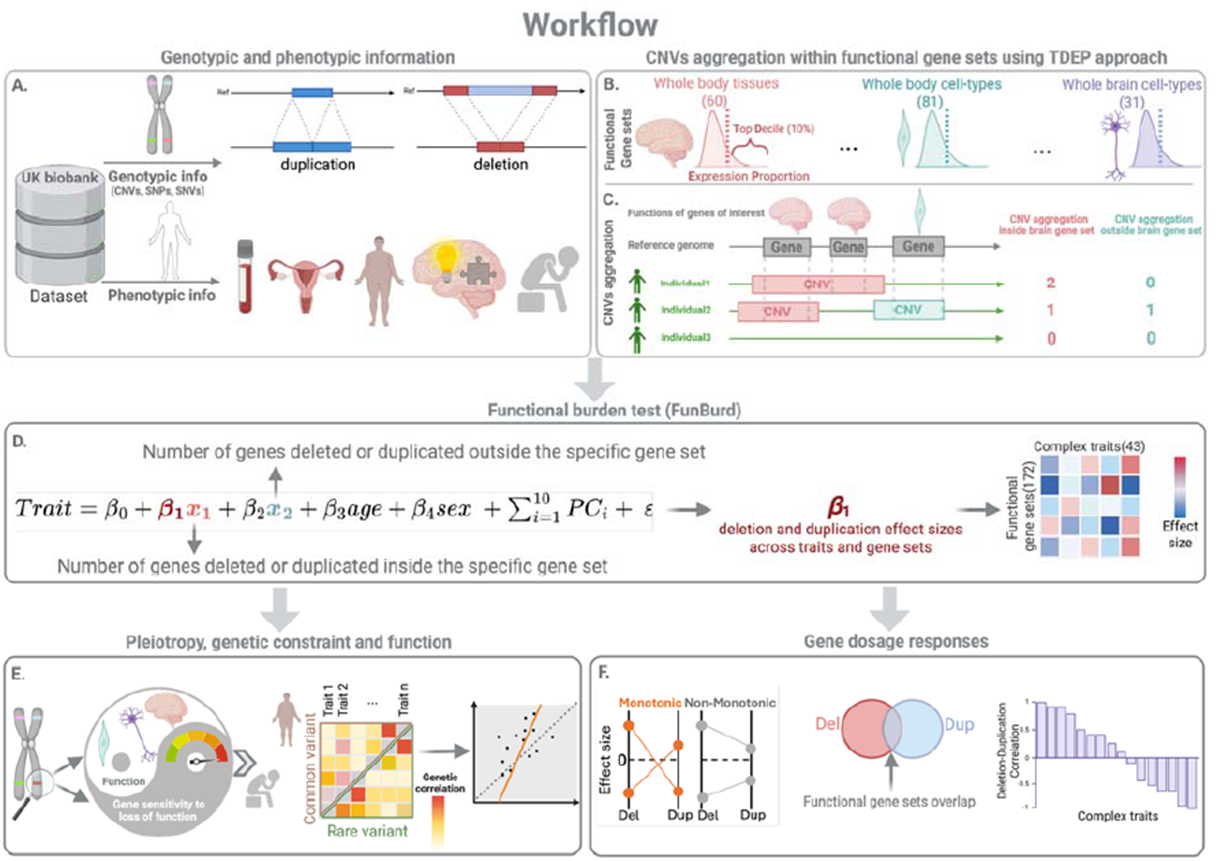
Estimating CNV effects on complex traits and downstream comparative analysis. **A)** We used CNVs and 43 phenotypes from the UK Biobank. **B)** We defined partially overlapping functional gene sets for 60 tissues, 81, and 31 whole body and brain cell types using TDEP (Top Decile Expression Proportion). C) CNVs were aggregated within each of the 172 gene sets. D) We performed functional burden tests to estimate the associations between CNVs disrupting 172 gene sets and 43 complex traits (IRNT scaled). Tests were performed separately for deletions and duplications. Downstream analyses systematically investigated (E) the relationship between gene constraint and function, and rare and common variant architectures of complex traits, and (F) monotonicity, correlation, and functional overlap in deletion and duplication effect size profiles. *Figure created with BioRender*.

We quantified the effect size of biological functions disrupted by CNVs for each of 43 complex traits—categorized into brain-related (e.g., cognitive metrics, mental health) and non-brain-related (e.g., blood assays, physical measures, reproductive factors)—in ∼500,000 UK Biobank participants. We compared these CNV burden associations to those of common variants and loss-of-function single nucleotide variants (SNVs)^18,20,27–30^. Furthermore, we systematically characterize the differences between deletions and duplications by quantifying gene dosage responses and performing CNV-burden correlations across traits. This is, to our knowledge, the first large-scale investigation of gene dosage across multiple categories of traits and biological functions.

## RESULTS

### Functional burden pleiotropy is higher for genes assigned to brain tissue

To identify gene-dosage-sensitive functions relevant to tissues across the human body, we performed functional burden analyses to test the association between 43 complex traits (blood assays, physical measures, cognitive, mental health measures, and reproductive traits, **Table ST1**) and 60 gene sets assigned to 60 whole-body tissues (**Table ST2**) which on average showed an overlap of 7.9% (**Methods, Fig. S1, Table ST3**). We chose these 43 phenotypes (**Table ST1**) relevant to a broad spectrum of medical conditions, because their associations with common variants and rare SNVs have been previously studied ^20,27,29^. Out of the 2580 associations (43 traits by 60 gene sets for deletions and duplications separately, **Table ST4**), CNVs showed FDR-significant association in all 5 trait categories (**Figs. 2A, S2-S4**). Deletions showed a higher proportion of FDR significant associations than duplications (n-del=568 [22%], n-dup=352 [13.6%]; proportion test p-value=1.2e-14, **Fig. 2B, Table ST5**).

**Fig. 2:**
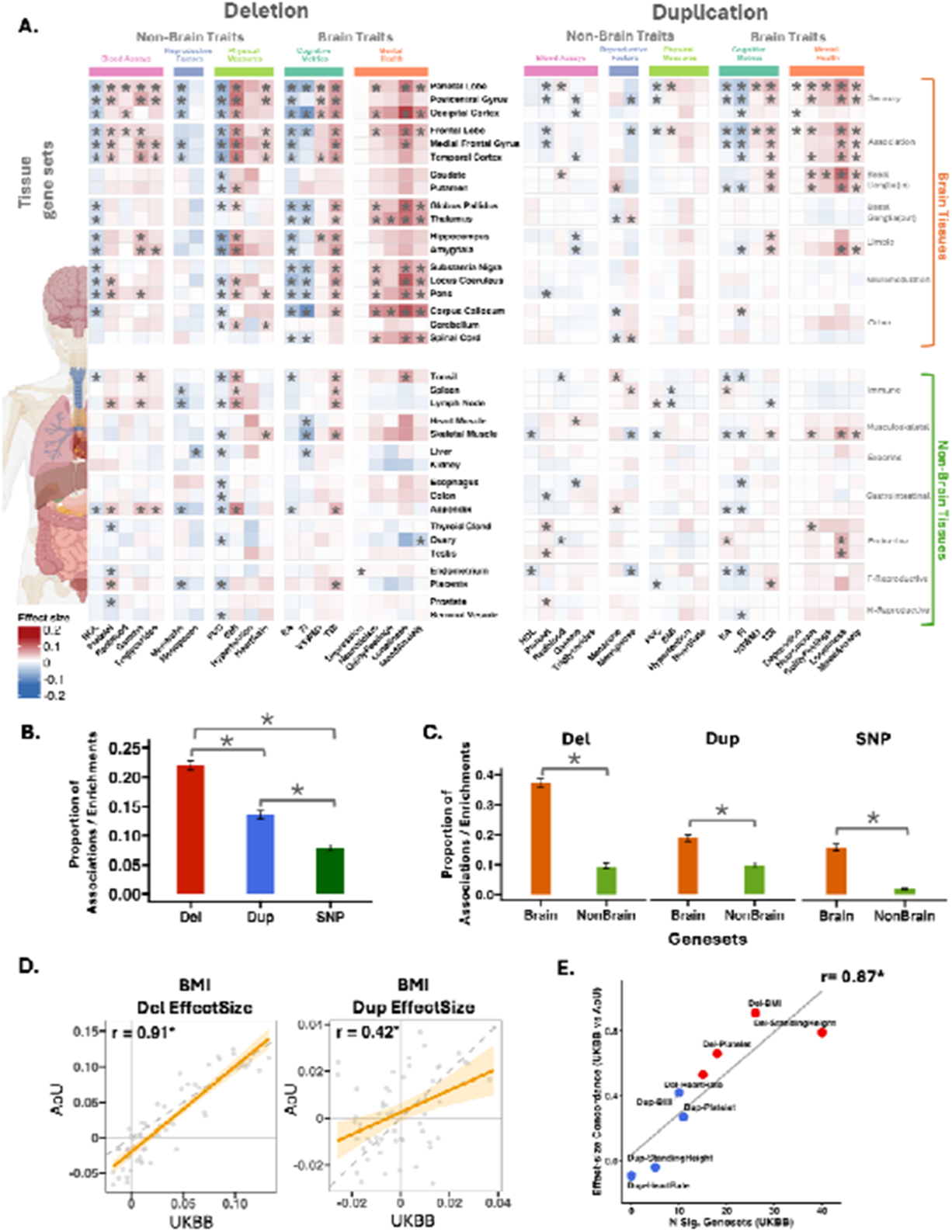
Tissue-specific associations of CNVs with complex traits. **A)** The heatmap displays association effect sizes between five categories of traits (x-axis) and tissue-specific gene sets (y-axis) for deletions (left) and duplications (right). The five trait categories are shown along the x-axis (top), and gene sets are listed on the y-axis, with their categories/annotations indicated on the right side. The color intensity reflects the direction and magnitude of the association (blue = negative effect size; red = positive effect size). Black asterisks (*) indicate statistically significant associations between traits and genes (FDR correction across all gene sets and traits, 172 ⍰ 43 ⍰ 2 =14,792 tests). Only a representative subset of traits is represented (the complete heatmaps are shown in **Figs. S2-S3). B-C)** Summary of CNV association differences. Bar plots (with standard error) summarizing the differences in the level of association/enrichment between the type of variants for **(B)** Whole-Body Comparison: Differences in association/enrichment levels across type of variants for all tissue gene sets; **(C)** Higher pleiotropy in brain tissue gene sets. : Bar plots represent the proportion of significant trait associations for brain and non-brain functional gene sets. **D)** The relationship between geneset effect sizes across the AoU and UKBB cohorts measured by the Pearson correlation; asterisks denote significant P-Jaccard. **E)** Relationship between discovery power and cross-cohort replication. Level of replication (correlation) across BMI, Heart Rate, Platelet Count, and Standing Height. Per-trait cross-cohort concordance of 60 gene-set effect sizes is plotted against the number of FDR-significant gene sets identified in the UKBB discovery analysis. Note: Townsend Deprivation Index (TDI) is an area-level measure of material deprivation, grouped with brain-related traits based on its established genetic and phenotypic correlations with cognitive and mental health outcomes^89,90^. **Abbreviations:** HDL: high-density lipoprotein; HbA1c: glycated haemoglobin; BMD: bone mineral density; BMI: body mass index; EA: educational attainment; FI: fluid intelligence; TDI: Townsend deprivation index; VSWM: visuospatial working memory; FVC: Forced vital capacity; Del: deletion; Dup: duplication; SNP: single-nucleotide polymorphism; AoU: All of Us.

Because CNVs are strong contributors to neurodevelopmental and psychiatric disorders^15,16,31^, we stratified functions into brain and non-brain gene sets. We define functional burden pleiotropy as the number of traits significantly associated with a gene set through CNV burden, extending the classical gene-level concept of pleiotropy to the gene-set level (**Methods**). Brain tissue gene sets showed higher functional burden pleiotropy, a greater proportion of significant associations, compared to non-brain gene sets, for both brain and non-brain traits (FDR-significant proportion test, Del p-value<1e-16, Dup p-value=2.2e-10, **Figs. 2C and S5A, Table ST6**). Sensitivity analyses (**Fig. S6, Table ST7**) showed that results were not influenced by i) removing correlated traits (**Fig. S7**); ii) different RNA-seq datasets (GTEx^32^ (**Figs. S8-S9**) and Human Protein Atlas^33^ (**Figs. S10-S11**); iii) phenotype measures methods (**Fig. S6E**), iv) age (<60 versus > 60 years), and sex (Female and Male, **Fig. S6**); v) ancestries (**Fig. S6B**); and vi) after restricting to a filtered set of 15 independent traits and gene sets with pairwise Jaccard overlap ≤ 20% (**Fig. S12**). Results were unchanged after removing recurrent CNVs carried by more than 20 individuals (population frequency of 1/20000; effect size r = 0.88 for deletions and 0.96 for duplications; pleiotropy r = 0.81 and 0.91; **Fig. S13**) and after excluding CNVs encompassing ≥ 5 genes (**Fig. S14**). Differences between deletion and duplication association patterns were not explained by carrier-count imbalance (**Fig. S15**), and carrier counts did not differ between brain and non-brain gene sets (Wilcoxon p = 0.15 for deletions, p = 0.69 for duplications; **Fig. S16**).

Replication in an independent cohort and orthogonal validation against rare loss-of-function variants further supported these findings. In the All of Us cohort (**Methods**), effect size and p-value concordance with UKBB was high for deletions (Pearson r up to 0.91, **Figs. 2D-E, S17, S18, Tables ST8-ST9**). Duplication effect size and p-value concordance was weaker (Pearson r up to 0.42). The level of concordance was directly related to the strength of CNV-trait association: traits with more FDR-significant gene sets in UKBB showed higher cross-cohort correlations (**Fig. 2E**). Deletion burden also showed 88.6% mean sign concordance with aggregated predicted loss-of-function (pLoF) burden from GeneBass ^34^ exome sequencing (binomial test, FDR q < 0.05 for 21 of 24 traits; **Methods, Fig. S19, Table ST10**).

We observed specific dosage-dependent effects across distinct brain structures and functions (functional gene set categories/annotations, **Fig. S4**). For duplications, cortical effects were stronger in association cortex than in sensorimotor cortex, consistent with the cortical brain organization hierarchy, previously characterized at the molecular, cellular, and functional levels^35–37^, suggesting an interplay between gene dosage and cortical organization hierachy^36,38^. At the subcortical level, duplications and deletions showed effects on brain traits through the input and output (respectively) of the basal ganglia system^39^, a group of nuclei that modulate cortical activity to influence motor control, as well as decision-making and executive functions. Finally, dosage specificity was also evident in regions implicated in neuromodulation (neurotransmitter release, such as locus coeruleus (norepinephrine)^40^, substantia nigra (dopamine)^41^, and brain stem structures of pons and medulla oblongata (acetylcholine)^42^) that were predominantly sensitive to deletions.

Sex differences impact the genetic architecture of the complex traits^43^, including mental health^44^. To assess this, we ran sex-stratified CNV burden associations across functional gene sets and trait categories (**Figs. S20-S25, Table ST11**). Although overall male and female CNV burden association patterns were similar to the main analysis, for mental health measures, a higher proportion of deletion compared to duplication associations were observed for females only (proportion test, FDR corrected p-value=2.4e-8, for females, **Figs. S22, S25**).

Direct comparison between sexes confirmed that females showed a higher proportion of significant deletion associations for mental health traits than males (15.3% vs. 6.7%, FDR q < 0.05), driven primarily by Mood Swings and Mood/Anxiety phenotypes (**Table ST12**). These female-specific gene dosage sensitive associations for mental health were notable for sensorimotor, association, and basal-ganglia gene-sets.

Finally, we asked if these observations were similar for common variants, by performing S-LDSC^5,28^ using the same traits in the same dataset with the same gene sets (**Methods, Fig. S26, Table ST13**). While the proportion of significant gene sets was lower (**Fig. 2B**), common variants showed higher levels of association/pleiotropy for the brain compared to non-brain gene sets (**Fig. 2C**), and this remained true when accounting for correlated traits (**Fig. S12B**).

### Gene dosage effects across whole-body and whole-brain cell types

We then asked if differences in pleiotropy observed for tissues were also observed at the cell type level. To do so, we defined 81 gene sets assigned to 81 whole-body single-cell clusters (**Table ST2**, from Human Protein Atlas, HPA)^45^, with an average overlap of 4.6% (**Fig. S27, Table ST3**). Functional burden analyses showed that 13% of the 6,966 trait / cell-type associations were FDR significant for deletions and duplications (**Figs. 3A-3B, S28-S29**, and **Table ST4**). These associations replicated in the All of Us cohort, with effect size concordance for deletions reaching r = 0.7 (**Fig. 3D-3E, Table ST8**). Significant associations were observed across HPA cell type groupings and all 5 trait categories (**Fig. S30**). Deletions showed a higher proportion of significant association than duplications (n-del=494 [14.2%], n-dup=421 [12%]; proportion test p-value=4.8e-3, **Fig. 3B**).

**Fig. 3:**
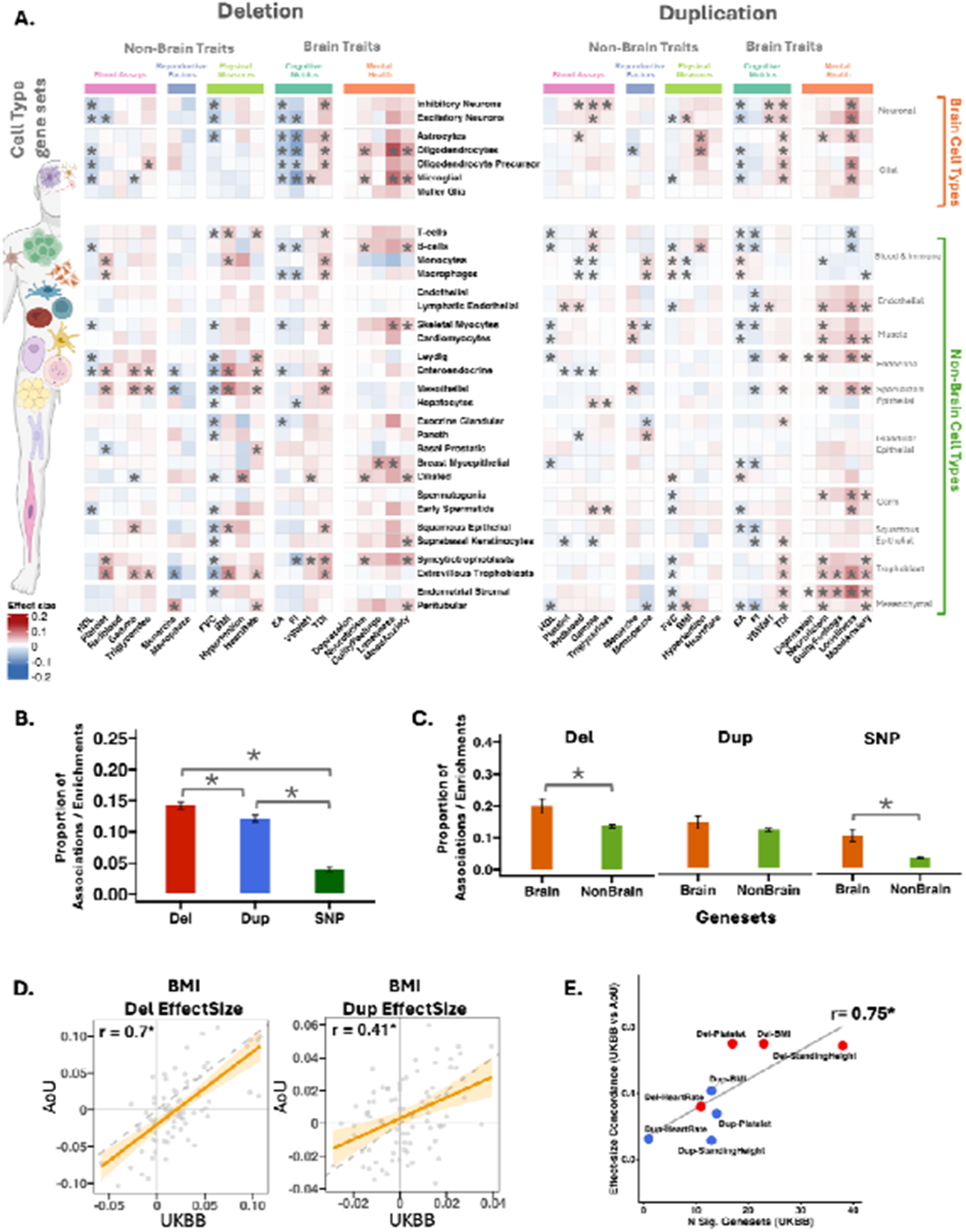
Whole-body cell type-specific associations of CNVs with complex traits. **A)** Heatmap displays a representative set of association effect sizes between five categories of traits (x-axis) and deletions and duplications aggregated by whole-body cell types (y-axis). Traits are categorized and shown along the x-axis, while the y-axis lists gene sets, with their categories/annotations indicated on the right side. The blue and red intensity color scale reflects the negative and positive effect sizes. Black asterisks (*) indicate statistically significant associations between traits and genes (FDR correction across 172 ⍰ 43 ⍰2 =14,792 tests). Only a representative subset of traits is represented (the complete heatmaps are shown in **Figs. S28-S29). B-C)** Summary of CNV association differences. Bar plots (with standard error) summarizing the differences in the level of association/enrichment between the type of variants for **(B)** Whole-Body Comparison: Differences in association/enrichment levels across type of variants for all whole-body cell type gene sets; **(C)** Higher pleiotropy in brain cell types gene sets. : Bar plots represent the proportion of significant trait associations for brain and non-brain functional gene sets. **D)** The relationship between geneset effect sizes across the AoU and UKBB cohorts measured by the Pearson correlation; asterisks denote significant P-Jaccard. **E)** Relationship between discovery power and cross-cohort replication. Level of replication (correlation) across BMI, Heart Rate, Platelet Count, and Standing Height. Per-trait cross-cohort concordance of 81 gene-set effect sizes is plotted against the number of FDR-significant gene sets identified in the UKBB discovery analysis. **Abbreviations:** HDL: high-density lipoprotein; HbA1c: glycated haemoglobin; BMD: bone mineral density; BMI: body mass index; EA: educational attainment; FI: fluid intelligence; TDI: Townsend deprivation index; VSWM: visuospatial working memory; FVC: Forced vital capacity; Del: deletion; Dup: duplication; SNP: single nucleotide polymorphism; AoU: All of Us.

Similar to tissue gene sets, the brain, compared to non-brain cell type gene sets, exhibited a higher proportion of significant associations (Deletion, p-value=5.5e-03, **Fig. 3C, Table ST6**). Beyond pleiotropy, we also observe some level of specificity with brain and non-brain cell type gene sets showing a higher proportion of associations with brain (Deletion, FDR corrected p-value=1.8e-2) and non-brain (Deletion, FDR corrected p-value=2.2e-7) traits, respectively (**Fig. S5, Table ST14**). Extending the systematic comparison to common variants using the same cell-type gene-sets showed higher levels of association for the brain compared to non-brain gene sets (**Figs. 3C, S31, Table ST6**).

We also observed distinct dosage-dependent effects across cell-type groupings (functional gene set categories/annotations, **Figs. 3D, S30**). Specifically, deletions and duplications effects on cognitive measures were primarily driven by glial and neuronal gene sets, respectively. For mental health measures, while deletions showed higher overall associations compared to duplications for glial gene sets, duplications exhibited an overall higher proportion of significant associations than deletions across all cell type gene sets (**Fig. S30**). Finally, when assessing the impact of sex, sex-stratified CNV burden associations across functional gene sets and trait categories showed that overall male and female CNV burden association patterns were similar to the main analysis (**Figs. S32-S37**).

Building on these observations, we focused on human brain cell types, using single-cell RNA-seq data delineating 31 adult brain cell type clusters^46^ (**Table ST2**) with an average overlap of 7.5% (**Fig. S38, Table ST3**). Both neuronal and non-neuronal cell types exhibited pleiotropic effects when disrupted by CNVs (**Figs. S39-S41, Table ST6**). These associations extended to both brain and non-brain traits, with neuronal cell types showing a higher proportion of associations with brain traits than non-neuronal ones, specifically for deletions, suggesting stronger brain-specific relevance of neuronal CNV burden (**Fig. S5C, Table ST14**). For common variants, neuronal gene sets show higher levels of pleiotropy than non-neuronal ones (**Fig. S5, Table ST6**).

### Dissecting functional burden pleiotropy, gene function, and genetic constraint

Previous studies have shown that common and rare variant heritability enrichments are strongest in constrained genes across complex traits ^22,27,29,47,48^. We, therefore, asked if the levels of pleiotropy observed above were related to genetic constraint. We observed that functional gene sets with higher proportions of genes under constraint (LOEUF top-decile, **Methods**) showed a higher level of pleiotropy for deletion (r=0.39, FDR corrected p-Jaccard=7.5e-3, **Fig. 4A**), and duplication (r=0.44, FDR corrected p-Jaccard=3e-3, **Fig. 4A**). This was also the case for functional enrichment analysis computed for the same gene sets and traits using common variant GWAS summary statistics (r=0.52, FDR corrected p-Jaccard=2.4e-2, **Fig. 4A**). This remained true regardless of the constraint metric (**Fig. 4C, Tables ST15-ST16**). Overall, brain gene sets showed significantly higher proportions of genes under constraint compared to non-brain gene sets (Wilcox Ranksum test p-value=3.5e-12, **Fig. 4B**). Using an approach normalizing functional burden association for genetic constraint (**Methods**, normative constraint modeling), we demonstrate that the most functional gene sets (∼90%) exhibited trait associations within an expected range of 5th to 95th percentiles (**Fig. 4D, Table ST16**). However, several gene sets exhibited significantly higher levels of pleiotropy than expected based on the proportion of genes under constraint (Wilcox Ranksum test, FDR corrected p-value < 0.05, **Figs. 4D-4E**). Overall, even after accounting for genetic constraint, the brain compared to non-brain gene sets showed higher levels of pleiotropy (Wilcox Ranksum test, FDR corrected p-value=2.1e-11, for Tissues, **Fig. 4D**).

**Fig. 4:**
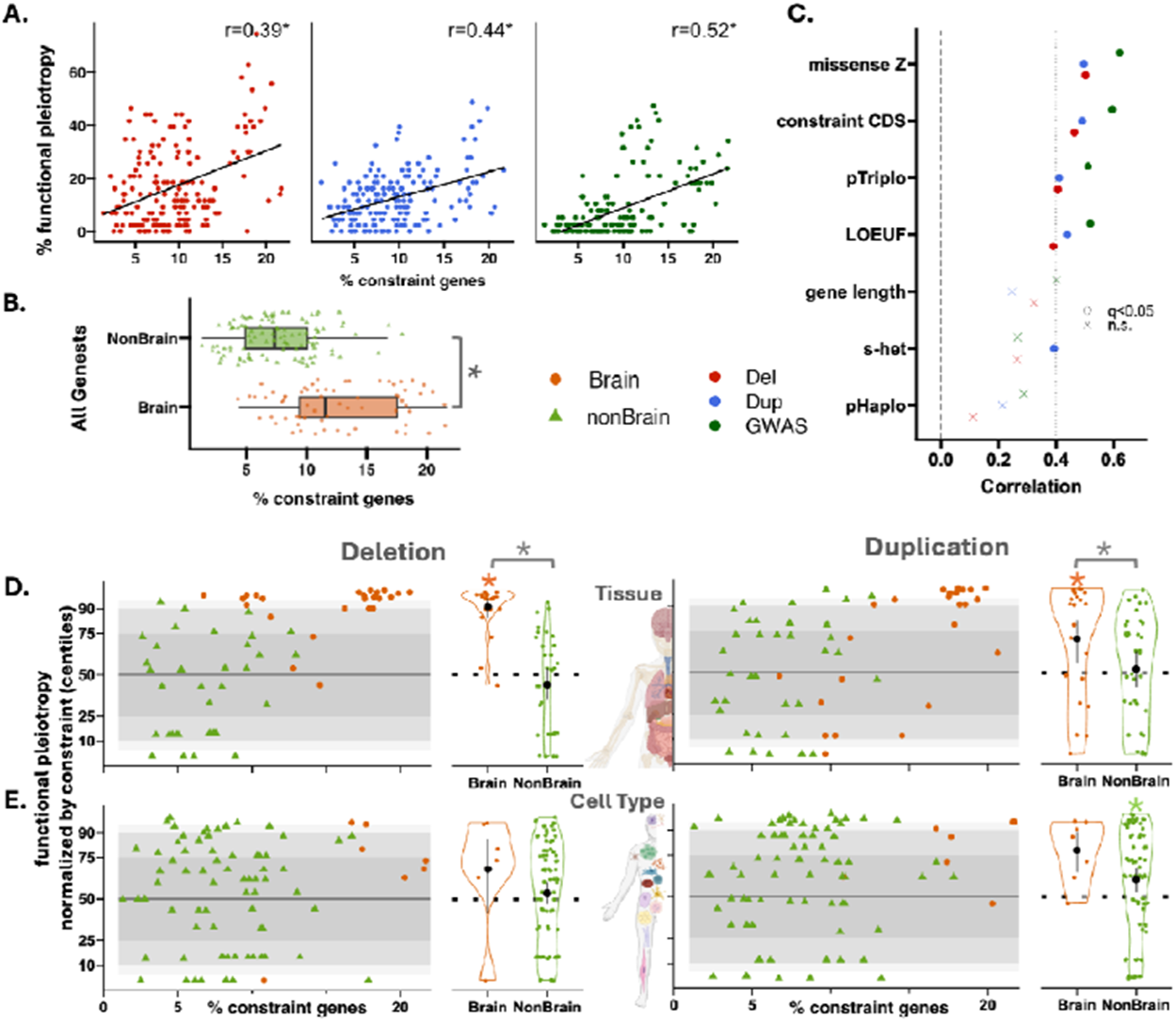
Dissecting functional burden pleiotropy, gene function, and genetic constraint. **A)** Correlation between the fraction of constraint genes (LOEUF top-decile) within a gene set and the number of traits showing significant associations with that gene set for deletions and duplications. Each data point is a gene set. Y-axis: the percentage of significantly associated traits for a variant (functional pleiotropy); X-axis: the percentage of top decile LOEUF genes within the gene set. * indicate FDR-adjusted P-value Jaccard (1000 nulls). **B)** Box plots show the distribution of constraint gene percentages for all Brain and Non-Brain gene sets (172). Each point represents a gene set, and asterisks (*) indicate statistically significant differences between groups. **C)** Shows the same information on correlations in panel (A) for different constraint metrics. q<0.05: FDR-adjusted P-value Jaccard (1000) are shown with circles; n.s.: not significant. **D)** Functional pleiotropy, normalized by the fraction of genetic constraint at the tissue level, is shown for 27 brain and 33 non-brain gene sets – deletions are presented on the left and duplications on the right. X-axis: represents the proportion of intolerant genes for the different gene sets. Y-axis: functional pleiotropy normalized by genetic constraint (centile). I.e., the 50th centile shows median functional pleiotropy computed across 1000 randomly sampled gene sets. Circles and triangles represent brain and non-brain gene sets, respectively. Gray shaded ribbons indicating 25th–75th (ribbon 1), 10th–90th (ribbon 2), and 5th–95th (ribbon 3) centiles. This is followed by violin plots showing the distribution of normalized functional pleiotropy across brain and non-brain traits. Orange and green asterisks demonstrate significantly (FDR-corrected, q < 0.05) increased functional pleiotropy compared to what is expected for a gene set with a comparable fraction of genetic constraint. The grey star indicates a significant difference between the brain and non-brain gene sets, normalized functional pleiotropy. **E)** The same analyses at the whole-body cell type level, including 7 brain and 74 non-brain gene sets. **Abbreviations:** CDS: coding sequence; Del: deletion; Dup: duplication; GWAS: genome-wide significant association; LOEUF: the loss-of-function observed/expected upper bound fraction; n.s.: non-significant; pHaplo: the probability of haploinsufficiency; pTriplo: the probability of triplosensitivity; SC: single cell; s-het: the fitness reduction for heterozygous carriers of a loss-of-function in any given gene; SNP: single nucleotide polymorphism.

### Between-trait CNV burden correlations and comparison to other variant classes

Studies often quantify pleiotropy and shared biological mechanisms^27,49^ through genetic correlation between pairs of traits. This approach has been widely used for common variants and recently extended to Loss of Function single-nucleotide variants (LoF SNVs)^27^, but has not yet been investigated for CNVs. We estimated the CNV-burden correlation between pairs of traits, separately for deletions and duplications, and tested their concordance with each other, as well as against previously published single-nucleotide polymorphism (SNP)-based and rare LoF SNV-based genetic correlations^27^ (**Fig. 5A, Table ST17**).

**Fig. 5:**
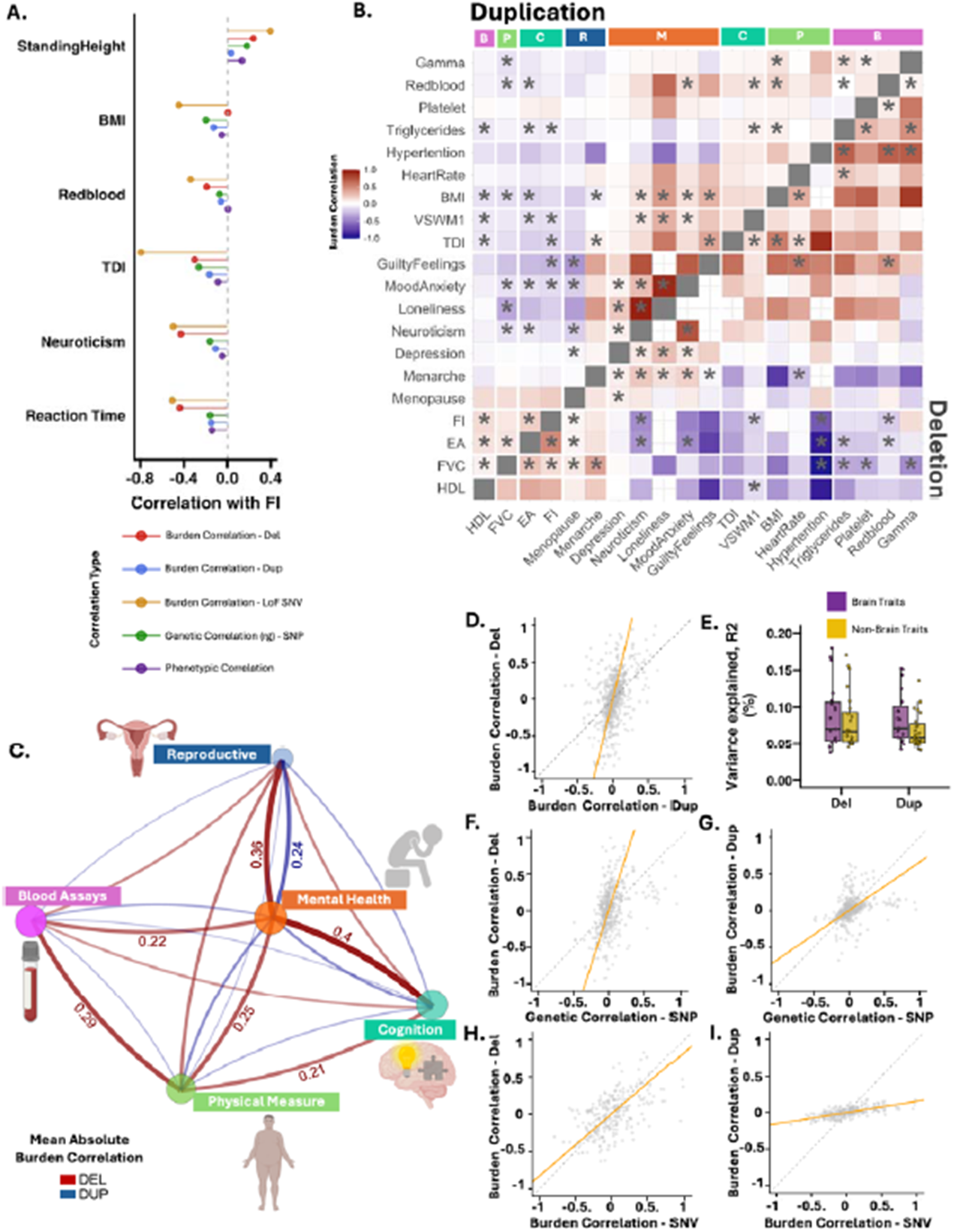
Comparison of rare and common variant architectures across complex traits. **A)** Dot plot illustrating the genetic and phenotypic correlations between fluid intelligence (FI) and six other traits., with each point color-coded by correlation type: SNP-based genetic correlations, deletion, duplication burden correlations, loss-of-function SNV burden correlation, and phenotypic correlations. **B)** The heatmap displays CNV burden correlations for trait pairs, where the lower triangle matrix shows correlations based on deletions and the upper triangle matrix shows correlations based on duplications. Significant correlations are marked by asterisks (*), based on FDR corrected p-values (p-Jaccard). The color bar on the top (x-axis) represents the five different trait categories: B: Blood Assays; P: Physical Measures; C: Cognitive Metrics; M: Mental Health; R: Reproductive Factors. **C)** A network plot visualizing the Mean Absolute CNV Burden Correlation between the five trait categories (Blood Assays, Physical Measures, Cognitive Metrics, Mental Health, and Reproductive Factors. Red and blue edges indicate deletion and duplication burden correlations, respectively. Edge thickness represents the strength of the mean absolute CNV burden correlation, with thicker lines showing a higher mean absolute burden correlation. Edge labels indicate the magnitudes of the strongest correlations (only values > 0.2 are shown). **D-I)** Comparison of between-trait correlations across variant types. **D)** A scatter plot comparing the CNV burden correlations between deletions and duplications. The orange line represents the Total Least Squares regression, and the dashed line indicates perfect concordance (y = x). **E)** Boxplots summarizing the variance explained by deletions and duplications aggregated across functional gene sets for brain and non-brain traits, determined via regression analysis (**Methods**). Concordance between CNV burden correlations and **(F-G)** SNP-based between trait genetic correlations and **(H-I)** loss-of-function (LoF) SNV burden correlations (phenotypic correlations are shown in **Fig. S42**). **Abbreviations:** BMI: body mass index; EA: educational attainment; FI: fluid intelligence; TDI: Townsend deprivation index; VSWM: visuospatial working memory; FVC: Forced vital capacity; Del: deletion; Dup: duplication; SNP: single nucleotide polymorphism; SNV: single nucleotide variants; LoF: loss of function;

Our CNV-burden correlation approach showed widespread between-trait correlations, with distinct patterns of correlations for deletions and duplications (**Fig. 5B**). Deletions exhibited stronger between-trait correlations across the five trait categories compared to duplications. Specifically, for deletions, the mean absolute burden correlations (between trait categories) exceeded 0.2 for the majority of trait category pairs. In contrast, this strong correlation (mean absolute burden correlation > 0.2) was observed for duplications only between mental health and reproductive traits (**Fig. 5C**). Notably, the between-trait correlations for mental health traits showed the highest mean absolute correlations with other trait categories for deletions, suggesting a pleiotropic impact of deletions that extends beyond the brain to encompass whole-body traits (**Figs. 5B-5C**).

Overall, the between-trait burden correlations were concordant across all classes of variants (**Fig. 5, Table ST18**). The sign concordance between types of variants ranged from 65% to 74% across all traits, which shows the consistent directionality of the between trait genetic correlations across types of variants (**Table ST18**). Deletion burden correlations were higher than those observed for duplications (4-fold, **Figs. 5D, S42-S43**), and SNPs (2.9-fold, **Fig. 5F**), and comparable to those observed for loss-of-function SNVs (0.82-fold, **Fig. 5H**). In contrast, duplication burden correlations were the lowest among all classes of variants (SNPs: 0.66-fold, **Fig. 5G**, LoF SNVs: 0.15-fold, **Fig. 5I, Table ST18**). These results suggest that deletions show high levels of pleiotropy, consistent with previous observations for loss-of-function SNVs^27^. Of note, variance explained estimates for CNVs were small (**Fig. 5E, Table ST19**) and could impact the accuracy of CNV burden correlation estimates. Between-trait CNV burden correlations were concordant between UKBB and All of Us (**Fig. S18E-S18F**), confirming that the shared genetic architecture across traits replicates independently.

We used formal mediation analysis to distinguish biological from mediated pleiotropy across six trait pairs with CNV burden correlations exceeding 0.3, using BMI as the mediator. Across trait pairs and gene sets, approximately 84% of the shared genetic architecture was attributable to the direct genetic effect, with a mean mediated proportion of 16% (**Fig. S44, Table ST20**). The exception was deletions for the BMI–HDL pair, which showed a higher mediated proportion (37.7%), consistent with specific metabolic dependencies for deletion-driven associations.

### Monotonic versus non-monotonic gene dosage responses across traits

Deletions and duplications represent a unique paradigm for comparing two classes of variants with opposing effects on gene expression^9^. While monotonic gene dosage responses (i.e., deletions and duplications have opposing effects on a trait, **Fig. 6A**) have been observed for some traits at specific genomic loci^50^, it has been challenging to characterize those at the phenome-wide and genome-wide level ^51,52^. We defined true gene dosage responses as gene-set–trait pairs in which both deletion and duplication burden associations were FDR-significant. We restricted our analyses to traits associated with at least 5 gene sets and gene sets associated with at least 2 traits (for robust statistics). We identified an excess (binomial test p = 4.8e-15) of non-monotonic (n=183 with same-direction effects) compared to monotonic responses (n= 62 with opposing direction effects; **Fig. 6C, Fig. S45A, Tables ST16, ST19**).

**Fig. 6:**
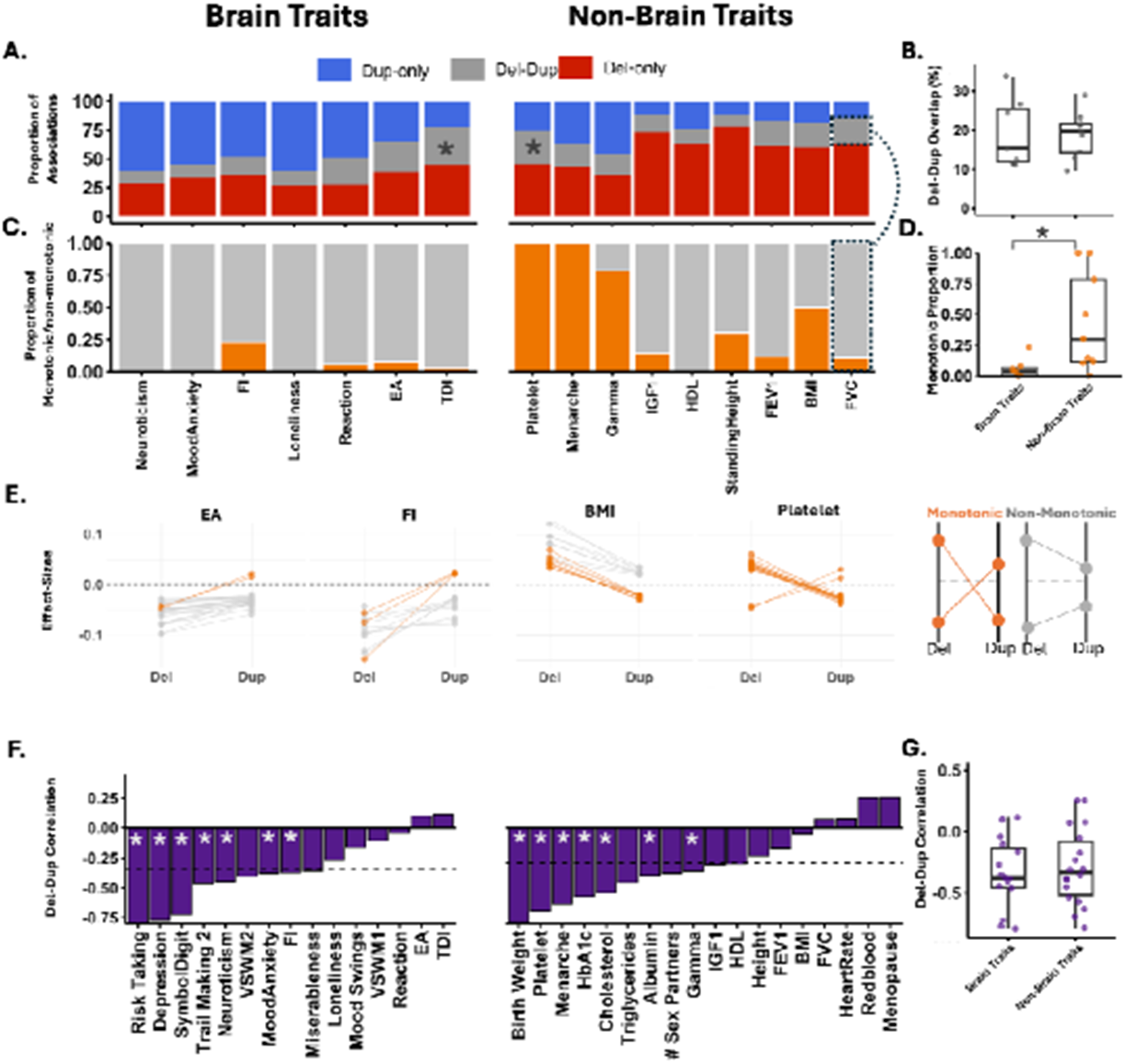
Gene dosage responses across traits and functional gene sets. **A)** Proportion of gene dosage responses significant for deletions-only, duplication-only, and both deletion-duplication, across brain and non-brain traits. Asterisks denote significant overlap (permutation test p-value; **Fig. S48). B)** Brain and non-brain traits showed similar levels of deletion-duplication association overlap. **C)** The proportions of monotonic and non-monotonic responses are shown across brain and non-brain traits. The total number of responses includes only FDR-significant gene sets’ effects for both deletions and duplications, and the proportions of monotonic (orange) versus non-monotonic (grey) responses are calculated within this subset. **D)** Box plots depict the distribution of monotonic responses for brain and non-brain traits; * indicates statistically significant (Wilcoxon rank-sum) differences between groups. **E)** Illustration of gene dosage responses across representative traits. Each line connects the effect sizes of deletion (left) and duplication (right) associations for a given functional gene set and a given trait. Orange lines represent monotonic responses (defined as significant effect sizes for both dosages with opposing directionality). Grey lines represent a non-monotonic response (same effect size directionality for both associations). **F-G)** Summary of deletion-duplication effect size correlations across traits. **F)** Barplot showing the deletion-duplication effect size correlations across traits. The * indicates significant correlation (FDR corrected p-Jaccard). **G)** Brain and non-brain traits show similar levels of deletion-duplication effect size correlations.

Brain traits associated with brain gene sets showed responses that were almost exclusively non-monotonic (96.1%), whereas non-brain traits showed substantially higher (41% versus 6%, Fisher’s exact test p=1e-9) monotonic proportions compared to brain traits (**Fig. 6C, Fig. S45**). At the gene-set level, genetic constraint was negatively correlated with the proportion of monotonic responses (Pearson r = −0.425, p = 0.008; **Fig. S45C**). Ability to detect monotonicity is not driven by different genes within a gene set affected by deletions and duplications (r = −0.08, p = 0.54; **Fig. S46**).

### Associations between functional gene sets and traits are variant-specific

The predominance of non-monotonic gene dosage responses raised the question of whether associations between traits and functional gene sets are specific to classes of variants. To address this, we examined variant-specific sensitivity across gene-set–trait associations. Of the 1,916 FDR-significant associations (out of 172 gene sets x 43 trait pairs tested), 85% were observed for either deletions (60%) or duplications (40%), and only 15% of them were significant for both (**Fig. 6A, Tables ST16, ST19**). Carrier-count differences (i.e., power imbalance) between variant types did not explain these deletion and /duplication-specific patterns (**Fig. S15**). Further characterizing this variant-specific pattern, we demonstrate that most traits (27 out of 33 traits, 82%) showed negative deletion-duplication effect size correlations (**Figs. 6F-6G, Table ST19**). Sensitivity analysis showed that this was consistent between UKBB and All of Us across all four replication traits (**Fig. S18D**). Overall, this suggests that associations between functional gene sets and traits are, in most cases, either deletion or duplication specific.

We reasoned that these variant-specific associations may extend to other classes of variants. We therefore quantified the overlap between gene sets showing CNV associations and common variant enrichments across the same 43 traits. Six brain traits showed significant functional gene sets overlap between common variants (S-LDSC enrichment analysis^5,28^) and duplications (permutation-based, **Figs. S47-S48**). We did not observe any significant overlap between deletions and common variants (**Figs. S47-S48**), despite the fact that deletions show a higher number of significant associations with traits compared to duplications (**Figs. 2B-3D**, FDR significant proportion test p-value<4.8e-3).

## DISCUSSION

This study represents the first comprehensive investigation into the biological determinants of pleiotropic effects of CNVs and gene dosage sensitivity. Genome-wide CNV functional burden tests revealed that pleiotropy was correlated to levels of genetic constraint. We also demonstrate higher levels of pleiotropy for the brain, compared to non-brain tissue and cell type gene sets, even after normalizing for genetic constraint. We observed distinct dosage-dependent effects across brain regions and cell types, with duplications preferentially linked to association cortical regions, and glial and neuronal functional categories contributing differently to cognitive and mental health traits. Shared genetic architecture between pairs of traits, indexed by between-trait burden correlations, was highest for deletions and loss-of-function SNVs compared to duplications and common variants. Surprisingly, most gene dosage responses were non-monotonic (deletions and duplications showed same-direction effects), and the proportion of monotonic responses decreased with genetic constraint. Consistent with paucity of monotonicity, associations between functional gene sets and traits were observed for either deletions or duplications, but rarely both. In addition, deletion and duplication effect sizes were negatively correlated, demonstrating that associations between traits and functional gene sets are variant-specific. Overall, brain-related gene sets were i) under higher genetic constraint, ii) showed higher levels of functional burden pleiotropy, and iii) had fewer monotonic gene dosage responses.

The robustness of our functional burden associations is supported by replication in an independent cohort and orthogonal validation. CNV burden associations discovered in UK Biobank were replicated in the All of Us cohort across four traits, despite substantial differences in sample size, ancestry, and continental origin between the two biobanks^53,54^. This is the first of a kind replication, providing strong evidence for the generalizability of the functional burden framework.

The observation of distinct dosage-dependent effects across brain regions and cell types provides key insights into the functional architecture of CNV pleiotropy. The preferential impact of deletions on sub-cortical gene sets and neuromodulation regions (e.g., locus coeruleus, substantia nigra) suggests that loss of function is particularly detrimental to fundamental motor, sensory, and regulatory systems. Conversely, duplications were primarily linked to association cortical regions, which are central to complex cognitive and executive functions. At the cellular level, these differences persist, as both glial and neuronal functional categories showed distinct contributions to cognitive and mental health traits depending on the variant type. These fine-grained functional distinctions demonstrate that CNV effects are not merely a broad dosage perturbation but are highly dependent on the affected functional module, potentially explaining the diverse clinical outcomes observed in patients.

FunBurd aggregating CNVs across biological functions demonstrates that rare CNVs genome-wide are associated with a broad spectrum of mental health, cognitive, physical, reproductive, and medical traits (such as blood assays, physical measures, and reproductive factors). Pleiotropic effects were higher in the brain compared to non-brain-related functional gene sets, even with respect to non-brain traits. This provides molecular-level support for the whole body multimorbid presentations in individuals with psychiatric and neurodevelopmental disorders, underscoring that psychiatric conditions share a substantial imprint of poor body health affecting unrelated bodily functions^55–57^. This has important implications for clinical interpretation and management. CNVs are commonly screened in individuals, often with a primary focus on neurodevelopmental disorders^58^, particularly within pediatric populations. Our results suggest that clinical management should be potentially reassessed when individuals transition from pediatric to adult care. The finding of a higher proportion of deletion associations for mental health traits in females suggests that sex-specific models of CNV risk propagation may be necessary for precise clinical stratification.

Disentangling genetic constraint from biological function has been challenging, and many studies have highlighted the predominant role of genetic constraint in shaping the genetic architecture of complex traits^22,27,28,47^. Using a novel approach to normalize for genetic constraint, we demonstrate that brain-related gene sets showed higher levels of pleiotropy compared to non-brain gene sets beyond what would be expected for gene sets with similar levels of genetic constraint^27,59^.

For deletions and duplications with opposing effects on transcription ^9^, it has been difficult to observe monotonic effects on complex traits^51^. Our FunBurd approach allowed us to quantify these opposing effects across 245 true gene dosage responses (gene-set–trait pairs significant for both deletions and duplications). Among these, non-brain traits have more monotonic gene dosage responses compared to brain traits. This may reflect the fact that the average population value is often the optimal one for non-brain traits (e.g., any deviation from median blood count values is associated with all-cause mortality)^60^. In contrast, median values are rarely optimal for brain traits (e.g., adaptive functioning, cognition, educational attainment, mood and depression are traits that are monotonically associated with all-cause mortality)^61– 64^.

We also demonstrate that gene sets under higher genetic constraint show lower proportions of monotonic gene dosage responses, an observation that, to our knowledge, has not previously been reported at the level of complex traits. This is consistent with prior work establishing that constrained genes are enriched for dose sensitivity in both directions, where haploinsufficiency and triplosensitivity independently impair function^65,66^. This intolerance has been shown at the molecular level for genes encoding subunits of multi-protein complexes, where too little or too much of a subunit disrupts complex assembly^65,66^. It is also evident at the population level, where constrained genes are depleted from both deletions and duplications in healthy individuals^67,68^. Our findings extend these molecular and population-level observations to the level of organismal phenotypes, suggesting that the narrow tolerable expression ranges characteristic of constrained genes propagate to trait-level non-monotonicity, whereby deviations from normal dosage in either direction produce phenotypic effects in the same direction.

Finally, we showed limited functional overlap between deletions and duplications, with the majority of functional gene sets being sensitive to either deletions or duplications, but rarely both, for a given trait. This suggests that different classes of genetic variation may exert their effects on traits through different biological processes^1,6,8,27^ This led to a negative deletion-duplication burden correlation for the majority of traits, which is more extreme than findings from previous studies showing discordant burden correlations between different classes of rare variant (e.g., missense and LoF variants) for a given trait, also implying that different classes of variants in the same genes often have divergent phenotypic effects^27^.

While the functional burden analysis adjusts for multigenic CNVs by statistically controlling for the count of disrupted genes outside the gene set of interest, we acknowledge that the full complexity of CNV architecture—including total CNV length, CNV count per individual, and underlying genomic context (e.g., segmental duplications)—presents a continuous challenge for rare variant association studies. To mitigate the risks of residual confounding and inflated associations due to gene set overlap (similar to LD structure), we incorporated several robust methodological safeguards. Specifically, we utilized a new permutation preserving Jaccard distance (P-Jaccard) method^69^ and LASSO/ridge regression to account for overlapping gene sets. The UK Biobank, as a relatively healthy population cohort^70^, poses limitations for disease analysis due to the under-representation of individuals with severe conditions and deleterious CNVs. This bias reduces our ability to detect associations between traits, conditions, and CNVs. However, using the UK Biobank’s highly harmonized genetic dataset, does offer some benefits including standardized genotyping and a consistent CNV calling pipeline, thereby minimizing technical artifacts that often plague heterogeneous CNV cohorts. The granularity of the gene sets is also a consideration. Tissue-level gene sets aggregate genes with potentially diverse functions, which is why we also analyzed 81 cell-type-specific gene sets that provide substantially finer resolution while maintaining comparable gene-set sizes. We did not use pathway-based gene sets such as GO terms because their highly variable sizes preclude direct comparisons of association strength across sets. Even so, our functional burden framework estimates the mean effect of gene dosage within a gene set. As a result, CNVs with heterogeneous effects across a given gene set may cancel or attenuate gene set-trait associations. This approach trades single-gene resolution for statistical power, which remains limited for coding CNVs even in samples exceeding 470,000 individuals^71^. Despite this trade-off, our gene-set-level findings are consistent with the broad pleiotropy reported at individual recurrent CNV loci^20,72^, indicating that the functional burden framework recapitulates and extends locus-level observations to a genome-wide view of CNV pleiotropy. Our functional burden association test provides critical insight into the interplay of genetic constraint, gene function, pleiotropy, and gene dosage response across human traits. We showed that the associations between gene functions and phenotypes are strongly modulated by classes of variants. Overall, these findings underscore the necessity of moving beyond locus-specific analyses to embrace functional and variant-specific models, thereby accelerating the deciphering of disease etiology and guiding interventions targeting the mechanistic basis of whole-body multimorbidity.

## Data Availability

This study utilizes data from the UK Biobank under application 40980. Access to UK Biobank data is subject to application and approval through their official portal (www.ukbiobank.ac.uk). Additional data and supplement material generated in this study are available upon reasonable request to the authors.
This study also utilizes data from the All of Us Research Program for replication purposes. Access to All of Us data is subject to registration and approval through the Researcher Workbench (www.researchallofus.org). Additional data and supplement material generated in this study are available upon reasonable request to the authors.

## SUPPLEMENTAL INFORMATION

Supplementary Figs. S1–S54, Supplementary Tables ST1-ST21.

## RESOURCE AVAILABILITY

### DATA AVAILABILITY

UK Biobank data was downloaded under the application 40980 and may be accessed via their standard data access procedure (see http://www.ukbiobank.ac.uk/register-apply). UK Biobank CNVs were called using the pipeline developed in the Jacquemont Lab, as described at https://github.com/labjacquemont/MIND-GENESPARALLELCNV. The final CNV calls are available for download from the UK Biobank returned datasets (Return ID: 3104, https://biobank.ndph.ox.ac.uk/ukb/dset.cgi?id=3104). References to the processing pipeline and R package versions used for analysis are listed in the methods. GWAS summary stats, heritability estimates, and genetic correlations for all the UK Biobank traits are publicly available and were downloaded from the NealeLab: https://www.nealelab.is/uk-biobank. SNV loss of function summary stats and burden correlations are publicly available (Genebass: https://app.genebass.org) and were downloaded from Weiner et al.^27^. Gene-level constraint data are available at https://gnomad.broadinstitute.org and other publications listed in methods.

### CODE AVAILABILITY

The code for generating all the Figures reported in the main analysis and supplement material can be found at the following GitHub link : https://github.com/SayehKazem/FunBurd

## FUNDING

This research was supported by Calcul Quebec (http://www.calculquebec.ca) and Compute Canada (http://www.computecanada.ca), the Brain Canada Multi-Investigator initiative, NIH U01 grant for CAMP (1U01MH119690-01), the Canadian Institutes of Health Research, CIHR_400528, The Institute of Data Valorization (IVADO) through the Canada First Research Excellence Fund, Healthy Brains for Healthy Lives through the Canada First Research Excellence Fund. Dr Dumas was supported by the Fonds de recherche du Québec - Santé (FRQ-S; 2024-2025 - CB - 350516) and IVADO. Dr Jacquemont is a recipient of a Canada Research Chair in neurodevelopmental disorders and a chair from the Jeanne et Jean Louis Levesque Foundation.

## COMPETING INTERESTS

The authors declare no competing interests.

## AUTHOR CONTRIBUTIONS

S.K., K.K., G.D., and S.J. designed the study, analyzed data, and drafted the manuscript. M.J.L., Z.S., and G.H. performed CNV calling and quality control. All authors contributed to the interpretation of the results and the editing of the manuscript.

## METHODS

### Participants

The UK Biobank recruited 502,534 individuals (54% female) ages 37 to 73 years, living in the United Kingdom, between 2006 and 2010^73^. Phenotypic measures were collected at the UK Biobank assessment centers (using touchscreen devices), or in an online follow-up. UK Biobank procedures contributing to this work comply with the ethical standards of the relevant national and institutional committees on human experimentation and with the Helsinki Declaration of 1975, as revised in 2008. Data was released under application number 40980. The ethics board of CHU Sainte Justine Research Center, Montreal, Canada, approved this study.

### Genotyping and CNV calling

UK Biobank DNA from blood samples was genotyped using the UK BiLEVE Axiom (n=49,950, 820k probes) and UK Biobank Axiom arrays (n=438,427, 807k probes)^53^. Quality control and processing followed our previously published pipeline^24,26,74^. 733,256 shared, QC-passed, hg19-mapped biallelic probes were used. Samples with high missingness (mind>0.05, |waviness factor|<0.05, log R ratio SD<0.35, B allele frequency SD<0.08) were excluded (n=28,522), leaving 459,855 individuals. CNVs were called using PennCNV^75^ and QuantiSNP^76^, combined with CNVision^31^, and concatenated with CNVs INHERITANCE ANALYSIS (CIAv.2.0), following our published pipeline (https://martineaujeanlouis.github.io/MIND-GENESPARALLELCNV/#/)^24,26,74,77^. CNVs were called using both algorithms with the following parameters: number of probe coverage per CNV ≥3, CNV size ≥1Kb, and confidence scores ≥ 15. CNVs detected by both algorithms were combined according to their types using CNVision to minimize the number of potential false discoveries. Following the data merging steps, CNVs were concatenated using the CNVs INHERITANCE ANALYSIS (CIAv.2.0) algorithm using the following criteria: a) CNV gapping ≤150 kb; b) CNV size ≥ 1000 bps; and c) number of probes ≥ 3. CNVs were filtered according to previously published studies^74^. We kept CNVs passing inclusion criteria such as confidence score ≥ 30 (with at least one of the detection algorithms), size ≥ 50 kb, and unambiguous type (deletions or duplications). All recurrent CNVs were verified visually.

CNVs were annotated using Gencode version 35 lifted to hg19 coordinates (https://www.gencodegenes.org/human/release_35lift37.html). We used bedtools intersect (https://bedtools.readthedocs.io/en/latest/) to identify breakpoints overlapping coding DNA elements such as UTRs, start and stop codons, exons, and introns^24,26,74^.

CNVs included in this study were ≥ 50 kb and fully encompassed at least one protein-coding gene. The size distribution and gene content of unique CNVs are summarized in **Fig. S49** and **Table ST21**. Among unique CNVs analyzed, 43.0% of deletions and 28.9% of duplications encompassed a single protein-coding gene; deletions and duplications encompassed a median of 2 and 3 genes respectively (mean: 3.55 and 4.64). Proportions of genes in tissue gene sets hit by observed CNVs are shown in **Fig. S50**, for BMI.

### Gene-sets

Gene sets were defined based on expression patterns across tissues and cell types (**Table ST2**). We used three resources:

a. Whole-body tissue expression data from FANTOM (Functional Annotation of the Mammalian Genome)^78^ – FANTOM provides whole-body normalized gene expression data (normalized Transcripts Per Million) across 46 tissues. For sensitivity analysis, we used two independent whole-body tissue expression data: GTEx ^32^ and the Human Protein Atlas ^45^ (https://www.proteinatlas.org/about/download; version 23).
b. Whole-body single-cell data from the Human Protein Atlas (HPA)^33^, which provides expression profiles across 81 cell types from 31 human tissues, based on single-cell RNA-seq (scRNAseq) data. In our analysis, we used the average expression data for 81 cell-type super-clusters across protein-coding genes (https://www.proteinatlas.org/about/download; version 23).
c. Whole-brain single-cell data from the Human Brain Cell Atlas v1.0 (snRNAseq)^46^, which consists of 3.369 million nuclei successfully sequenced using snRNAseq across 106 anatomical locations in adult brains. We used cell types from 31 superclusters and 461 clusters, aggregated data from (https://github.com/linnarsson-lab/adult-human-brain).

### Gene-set creation

We use the Top Decile Expression Proportion (TDEP) approach ^28,30^ to create tissue and cell-type gene sets (**Table ST2**). To do so, we divided each gene’s expression in a given tissue or cell type by its total expression across all tissues or cell types, producing values between 0 and 1. This metric identifies the proportion of a gene’s total expression attributed to a specific tissue or cell type. We then selected the top decile of these expression proportion measures to create each gene set.

### Traits preprocessing and normalization

We extracted 43 binary and continuous complex traits from the UK Biobank (UKBB, **Table ST1**). For traits with multiple measures over time, we only considered data from the initial visit and assessment. Individuals with missing trait measures were removed from the dataset. We excluded outliers by considering values beyond ±6 standard deviations (SD). To be consistent with previous GWAS for the same UKBB continuous traits, we used PHESANT ^79^, an inverse rank normalization transformation (IRNT). This method involves ranking the continuous data and then transforming these ranks into quantiles of the standard normal distribution^80^.

### Functional Burden Association Test (FunBurd)

FunBurd is designed to test the association between variants aggregated across a gene set and a given trait. The traits of interest were considered as a function of the number of genes within the gene set disrupted by CNVs. To avoid effect size inflation, due to multigenic CNVs, we adjusted for the number of genes (not members of the gene set) disrupted by the same CNV. We also adjusted for age, sex, and ancestry.

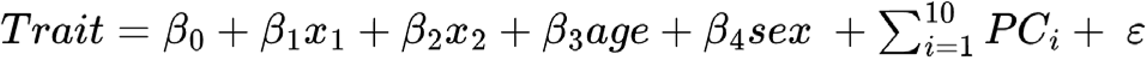

In this equation, x_1_ is the count of genes disrupted by CNVs within the target gene set for a given individual, and x_2_ is the count of disrupted genes outside the target gene set (adjusting for multigenic CNVs). The inside-set effect size (β_1_) and outside-set effect size (β_2_) are estimated jointly; full β_1_ and β_2_ statistics for all analyses are provided in **Table ST4**, and their distributions are compared in **Fig. S51**. The burden variable x_1_ follows a sparse, Poisson-like distribution across gene sets, with most carriers disrupting one or two genes per set (**Fig. S52**), limiting power to detect non-linear dose–response relationships; we therefore retained the linear model as the most parsimonious specification. The coefficient B_1_ represents the effect size of a given gene set on the scaled trait (IRNT) of interest. In this analysis, we examined 14,792 associations, derived from 172 gene sets, 43 traits, and 2 types of variations (deletions and duplications). Our group has recently explored similar burden analysis models for the analysis of a single trait^26,36,81,82^.

A significant functional burden association indicates that genes with preferential (relative) expression in a given tissue or cell type are, on average, sensitive to gene dosage change for that trait. This reflects the functional properties of the genes within the set and does not necessarily imply that a tissue or cell type is proximally involved in mechanisms modulating a given trait.

### Multiple comparison correction

All analyses were corrected for multiple testing using the Benjamini–Hochberg False Discovery Rate (FDR)^83^. FunBurd considered the entire matrix of 172 gene sets × 43 traits × 2 CNV types = 14,792 tests; associations with FDR-corrected q < 0.05 were considered significant.

### Functional Burden Pleiotropy

We define Functional Burden Pleiotropy as the number of traits significantly associated with a given functional gene set through CNV burden analysis. This extends the classical concept of pleiotropy, a single gene or variant influencing multiple traits, to the gene-set level, consistent with recent frameworks that aggregate variant-level effects within genes^27^ or across multigenic CNV loci^72^.

### Replication in the All of Us cohort

We replicated the FunBurd analysis in the All of Us (AoU) Research Program^84^ (N = 508,764 participants with genotyping data) for four traits with sufficient sample size and comparable phenotypic characterization across biobanks: BMI, Standing Height, Heart Rate, and Platelet Count. We applied the same CNV calling pipeline (PennCNV^75^ + QuantiSNP^76^), quality control, and FunBurd model used for the primary UKBB analysis. Effect size estimates and between-trait burden correlations were compared across cohorts (**Figs. 2D-2E, 3D-3E, and S17–S18**). To our knowledge, this represents the first genome-wide CNV study conducted in All of Us.

### Permutation preserving Jaccard distance (P-Jaccard)

Because gene sets partially overlap, correlations between gene sets may be inflated. To adjust the p-values of such correlations, we developed a method based on the Jaccard distance matrix. This approach performs permutations conditioned on the Jaccard distance of the gene sets and thus avoids inflated p-values by generating a plausible null distribution ^69,85,86^ (the corresponding Jupyter notebook is available on GitHub: https://github.com/SayehKazem/FunBurd).

### GWAS summary statistics, heritability estimates, and genetic correlations

We included the GWAS summary statistics for the same traits derived from nearly the same set of participants from the UK Biobank (**Table ST1**). GWAS summary statistics, heritability estimates, and genetic correlations for all the UK Biobank traits are available at https://www.nealelab.is/uk-biobank.

### Comparing functional overlap between CNVs and common variants

To systematically compare functional enrichments between CNVs and common variants, we used the same gene sets and UKBB traits to run functional burden association tests (FunBurd) and Stratified LD Score Regression (S-LDSC)^5^. S-LDSC assesses whether certain biological functions are enriched for common variant GWAS signals ^28^.

Functional overlap between CNVs and SNPs was defined as the proportion of gene sets enriched jointly in CNVs (FunBurd) and common variant associations (S-LDSC). The Functional overlap between deletions and duplications was the proportion of gene sets associated with both types of CNVs. Significance was tested by fixing the gene sets enriched in common variants and permuting CNV-associated gene sets 1000 times. We then tested whether the observed functional overlap was greater than expected by chance (null distribution).

### CNV-burden correlations

To compute CNV-burden correlations, we used the geneset-trait association profiles of deletions and duplications. Prior to this, we addressed two key issues: the overlap between gene sets and the heritability (variance explained) attributable to functional gene sets. To address gene set redundancy, we integrated two complementary analytical approaches: pairwise Jaccard distances to quantify overlap among gene sets, and LASSO (Least Absolute Shrinkage and Selection Operator) regression to perform variable selection and regularization (**Fig. S53**). The LASSO model ran on scaled (IRNT) residuals after regressing out the covariates (Sex, Age, and ancestry (PCs) from the scaled trait:

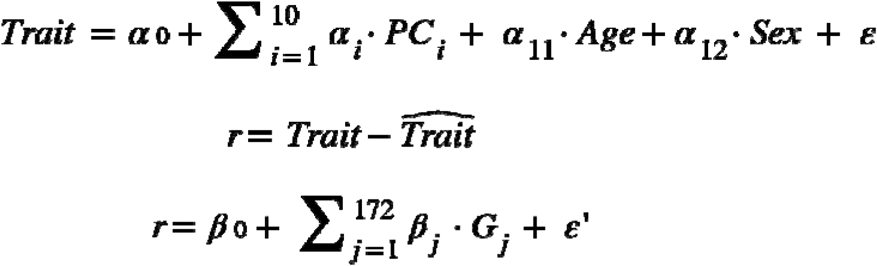

By considering these two measures, we excluded the gene sets that had a Jaccard similarity percentile greater than 0.2, that are also removed by LASSO in at least 80% of traits, from further analysis (n = 13; see **Fig. S53**). We use selected gene sets (159 out of 172) to calculate the variance explained by these gene sets for each trait (the model is applied on residuals after regressing out the covariance):

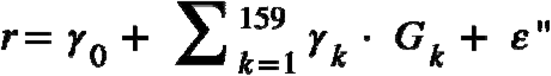

We then extracted the coefficient of determination (R^2^) from each regression model as a proxy for trait heritability (h^2^ or variance explained).

Finally, we computed deletion and duplication burden correlations between trait pairs using a genetic correlation formula:

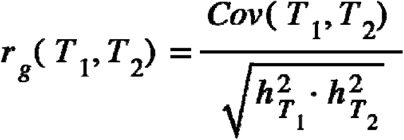

The numerator was defined as the covariance between shrinkage adjusted effect size profiles (excluding the 13 filtered gene sets), while the denominator represented trait specific heritability, estimated as the variance explained by effective gene sets using a linear regression model.

For binary traits, we first applied logistic regression to regress out covariates. The resulting residuals were then treated as continuous outcomes in downstream analyses to compute variance explained (the denominator). Moreover, to enable meaningful comparison of effect sizes and their covariances across binary–binary, binary–continuous, and continuous– continuous trait pairs (the numerator), we transformed the binary trait effect sizes to the liability scale. This transformation ensures that all effect sizes are on a comparable scale:

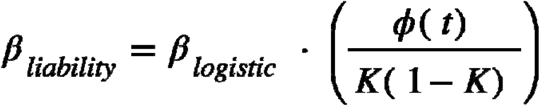

⍰_logistic_ is the logistic regression coefficient, K is the population prevalence of the binary trait, t is the threshold on the standard normal distribution corresponding to the prevalence, and ⍰ (t) is the standard normal density evaluated at t.

### LoF SNV summary statistics of burden correlations

SNV loss of function (LoF) summary stats and burden correlations were downloaded from https://pmc.ncbi.nlm.nih.gov/articles/instance/10614218/bin/NIHMS1933335-supplement-Supplementary_tables.xlsx^27^. Notably, these results were derived from the same set of UK Biobank participants ^27^.

### Genetic constraint metrics

We used several measures of constraint, including: i) the loss-of-function observed/expected upper bound fraction (LOEUF) score^22^; ii) probability of being loss-of-function intolerant (pLI)^22^; iii) missense Z-score^22^; iv) CDS: percentage of coding sequence (CDS) under evolutionary constraint^47^; v) two measures of dosage sensitivity ^87^: probability of haploinsufficiency (pHaplo), and probability of triplosensitivity (pTriplo); vi) s-het ^88^, a constraint metric predicted from multiple transcriptomic and gene features using machine learning; and vii) gene-length ^22,87^ in base pairs. We calculated a mean statistic of these measures of constraint for each of our 172 gene sets. To do this, we used genes that overlapped with the top decile (10%) of genes for each constraint measure^22^.

### Normative constraint modeling

We developed a normative model to evaluate whether the association between functional gene sets and traits is explained by genetic constraint. We generated a null distribution by randomly sampling gene sets 1000 times (n = 1,298 genes; without replacement) and using different proportions within constraint (top decile of LOEUF scores) and remaining (other LOEUF score deciles), then calculated the corresponding number of trait associations (1,720,000 individual Funburd associations, **Fig. S54**). This approach allowed us to define the expected relationship between the number of trait associations and the proportion of genes under constraint. Our normative model revealed a sharp increase in trait associations for gene sets with a constraint fraction exceeding 0.25, highlighting a nonlinear relationship between genetic constraint and pleiotropy. By providing a constraint-normalized effect size for each gene set, this approach offers a more refined interpretation of functional pleiotropy.

### Mediation analysis

To assess whether between-trait CNV burden correlations are partly attributable to mediated pleiotropy, where a genetic variant affects one trait that in turn causally influences another, we performed mediation analysis using BMI as a mediator for trait pairs with CNV burden correlations exceeding 0.3. Total effects were decomposed into direct (c⍰; biological pleiotropy) and indirect (a × b; mediated pleiotropy) components separately for deletions and duplications, across all FDR-significant gene sets. Analyses were performed in both forward (trait → BMI) and reverse (BMI ← trait) directions. Full path coefficients, standard errors, mediated proportions, and FDR-adjusted p-values are provided in **Table ST20**.

### Comparison with pLoF burden

To provide orthogonal validation, we compared deletion burden effect sizes with aggregated predicted loss-of-function (pLoF) burden from exome sequencing (GeneBass^34^) across 24 of the 43 traits. For each trait, we aggregated gene-level pLoF burden β estimates within each functional gene set (**Table ST10**) and computed sign concordance and Pearson correlations with the corresponding deletion CNV burden effect sizes. Comparisons were restricted to gene sets with nominal CNV burden significance (p < 0.05), as FDR-corrected thresholds yielded too few data points for meaningful concordance statistics. Sign concordance was tested against the 50% random expectation using a binomial test with FDR correction across the 24 traits.

## Notes

### Competing Interest Statement

The authors have declared no competing interest.

### Author Declarations

The Ethics Committee of UK Biobank approved the use of these data under application 40980. The Ethics Committee of All of Us approved the use of these data

### Summary of Updates

Independent Replication: We performed a comprehensive replication analysis using the All of Us (AoU) Research Program (N = 508,764). Results for BMI, height, heart rate, and platelet count showed strong effect-size concordance and significant sign concordance with the discovery cohort. Orthogonal Validation: Deletion burden findings were validated using rare loss-of-function (pLoF) variants from UKBB exome sequencing, demonstrating 88.6 percent mean sign concordance across 24 traits. Robustness Testing: We confirmed associations are not driven by large multigenic events by excluding CNVs encompassing 5 or more genes. We also confirmed that results are not driven by recurrent CNVs by removing variants with a population frequency greater than 1 in 22,000. Mediation Analysis: We implemented mediation models using BMI as a mediator to distinguish biological from mediated pleiotropy, finding that direct genetic effects explain the majority of shared architecture. Conceptual Refinements: The central concept was updated to "Functional Burden Pleiotropy" to clarify its definition at the gene-set level. Additionally, gene-dosage analyses were restructured to distinguish between true dosage responses and variant-specific sensitivity. Normative Model: The genetic constraint normative model was updated using 1,000 iterations to provide improved statistical power. Clarifications: We added comprehensive descriptive statistics for unique CNVs and clarified the classification of the Townsend Deprivation Index based on established socio-genetic correlations. Updated Figures and Tables: Main Figures 2, 3, 4 and 6 have been updated. We have also added further Supplemental Figures and updated the Supplemental Tables to reflect these comprehensive revisions.

