## Supplementary Figures for "Determinants of functional burden pleiotropy and gene dosage responses across human traits"

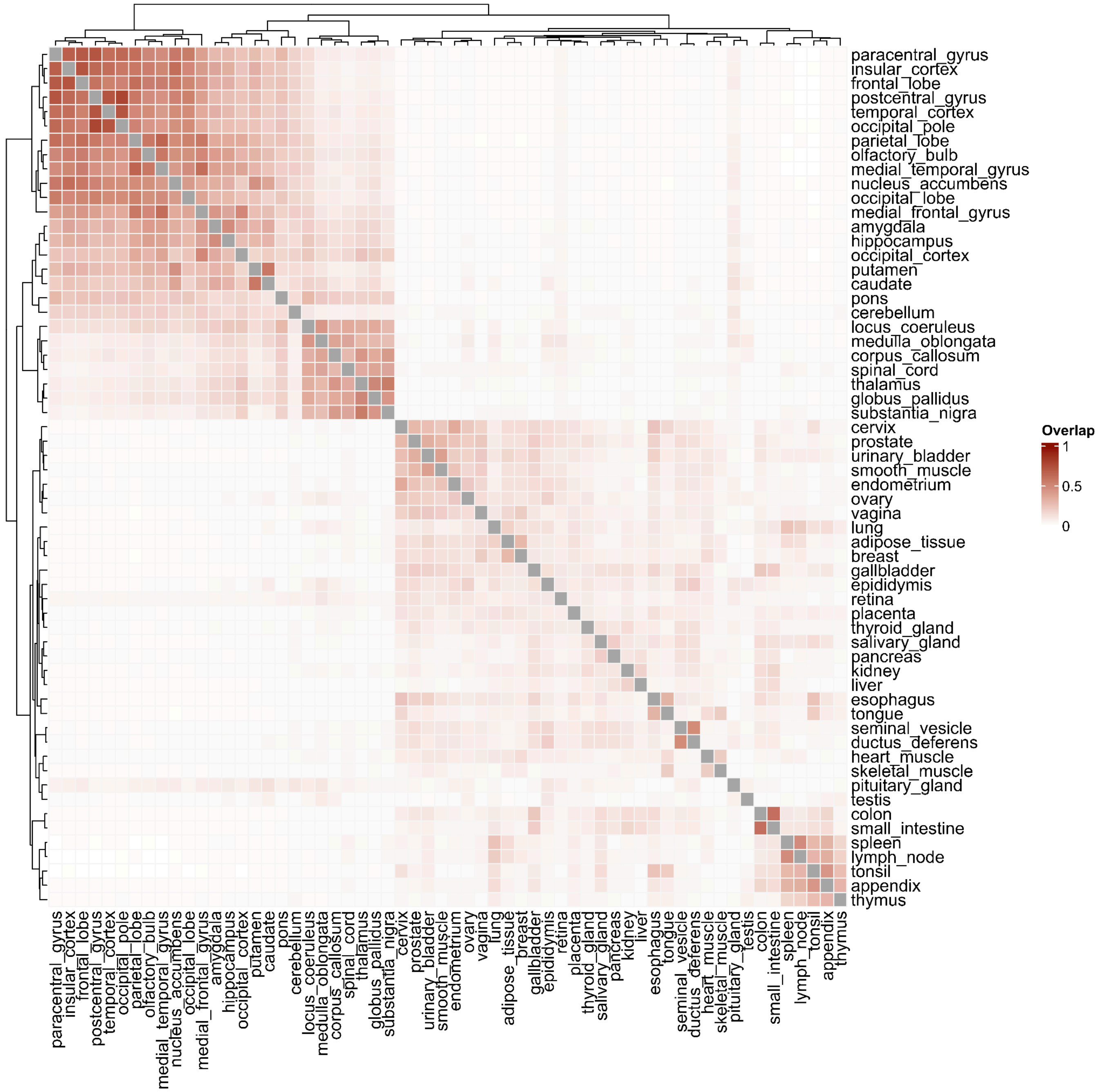


**Supplementary Figure 1: Pairwise overlap matrix (Jaccard overlap) of tissue gene sets**

The heatmap displays the pairwise overlap (1-Jaccard Distance) between tissue gene sets. Dark red indicates high overlap between gene sets, pink represents moderate overlap, and white reflects low overlap. The maximum, minimum, mean, and median values of the overlap are: 77.2%, 0.07%, 7.9% and 2.7%.

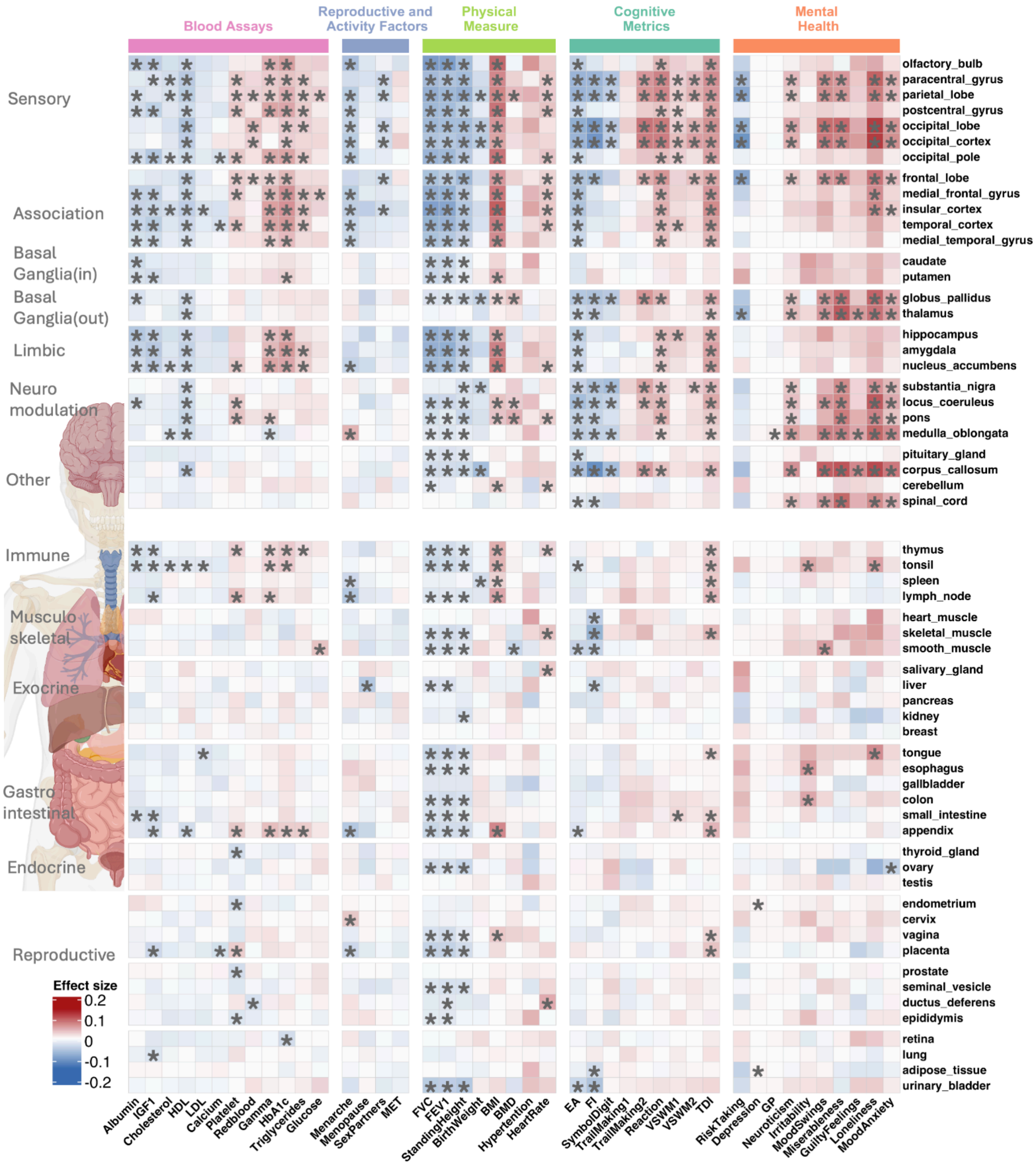


**Supplementary Figure 2: Heatmap of deletion effect sizes for whole-body tissue across traits**

The heatmap displays the effect sizes of deletion burden associations between five categories of traits (x-axis) and tissue-specific gene sets (y-axis). The five trait categories are shown along the x-axis (top), and gene sets are listed on the y-axis, with their categories/annotations indicated on the right side. The color intensity reflects the direction and magnitude of the association (blue = negative effect size; red = positive effect size). Black asterisks (*) indicate statistically significant associations between traits and genes (FDR correction across all gene sets, traits, and CNV type, 172 ✕ 43 ✕ 2 =14,792 tests). We tested all trait–gene set combinations to provide a complete picture of associations, including biologically unexpected combinations (Female-specific phenotypes (e.g., menopause) with male gene sets (e.g., testis)) that serve as negative controls.

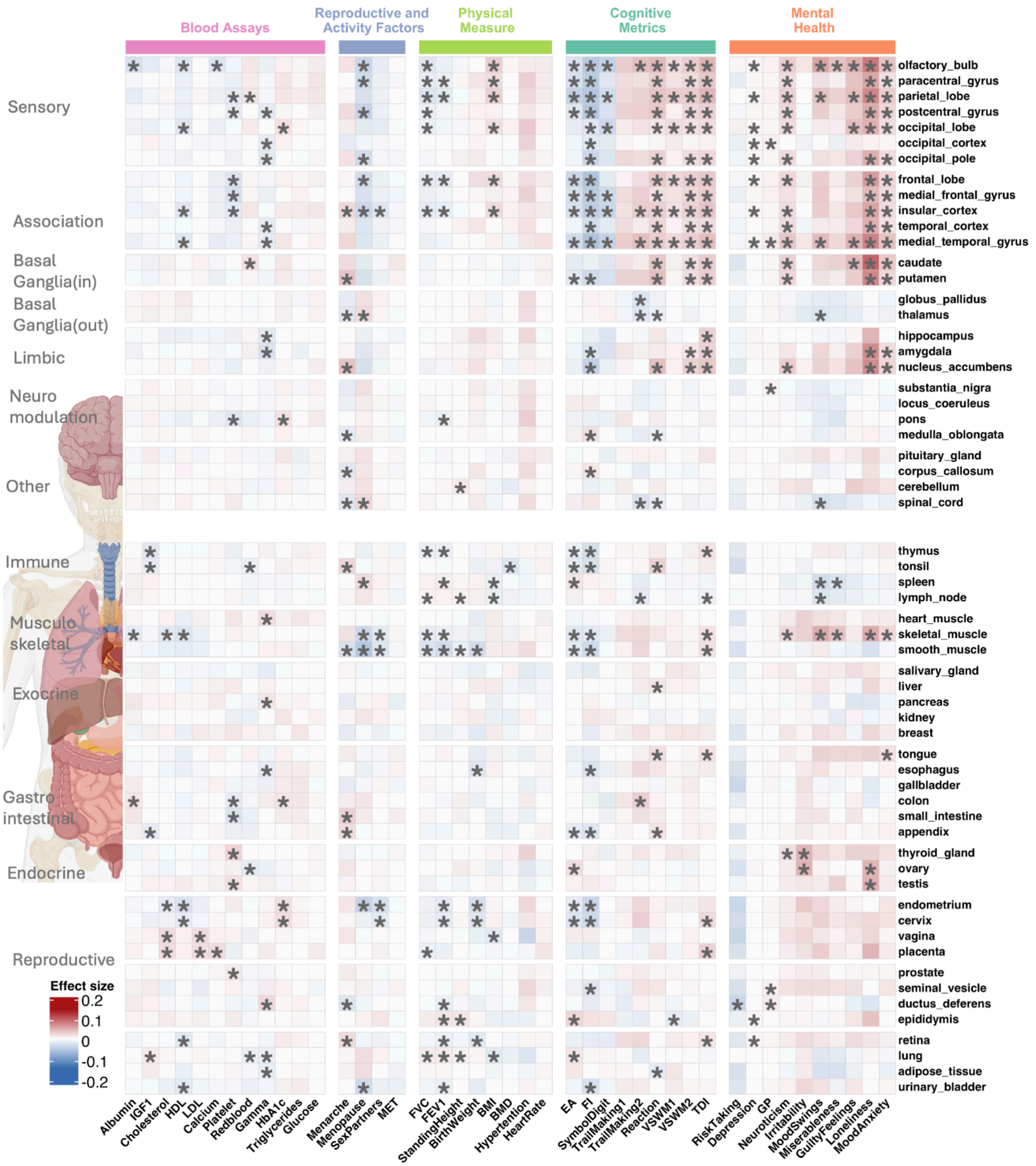


**Supplementary Figure 3: Heatmap of duplication effect sizes for whole-body tissue across traits**

The heatmap displays the effect sizes of duplication burden association between five categories of traits (x-axis) and tissue-specific gene sets (y-axis). The five trait categories are shown along the x-axis (top), and gene sets are listed on the y-axis, with their categories/annotations indicated on the right side. The color intensity reflects the direction and magnitude of the association (blue = negative effect size; red = positive effect size). Black asterisks (*) indicate statistically significant associations between traits and genes (FDR correction across all gene sets, traits, and CNV type, 172 × 43 × 2 =14,792 tests). We tested all trait–gene set combinations to provide a complete picture of associations, including biologically unexpected combinations (Female-specific phenotypes (e.g., menopause) with male gene sets (e.g., testis)) that serve as negative controls.

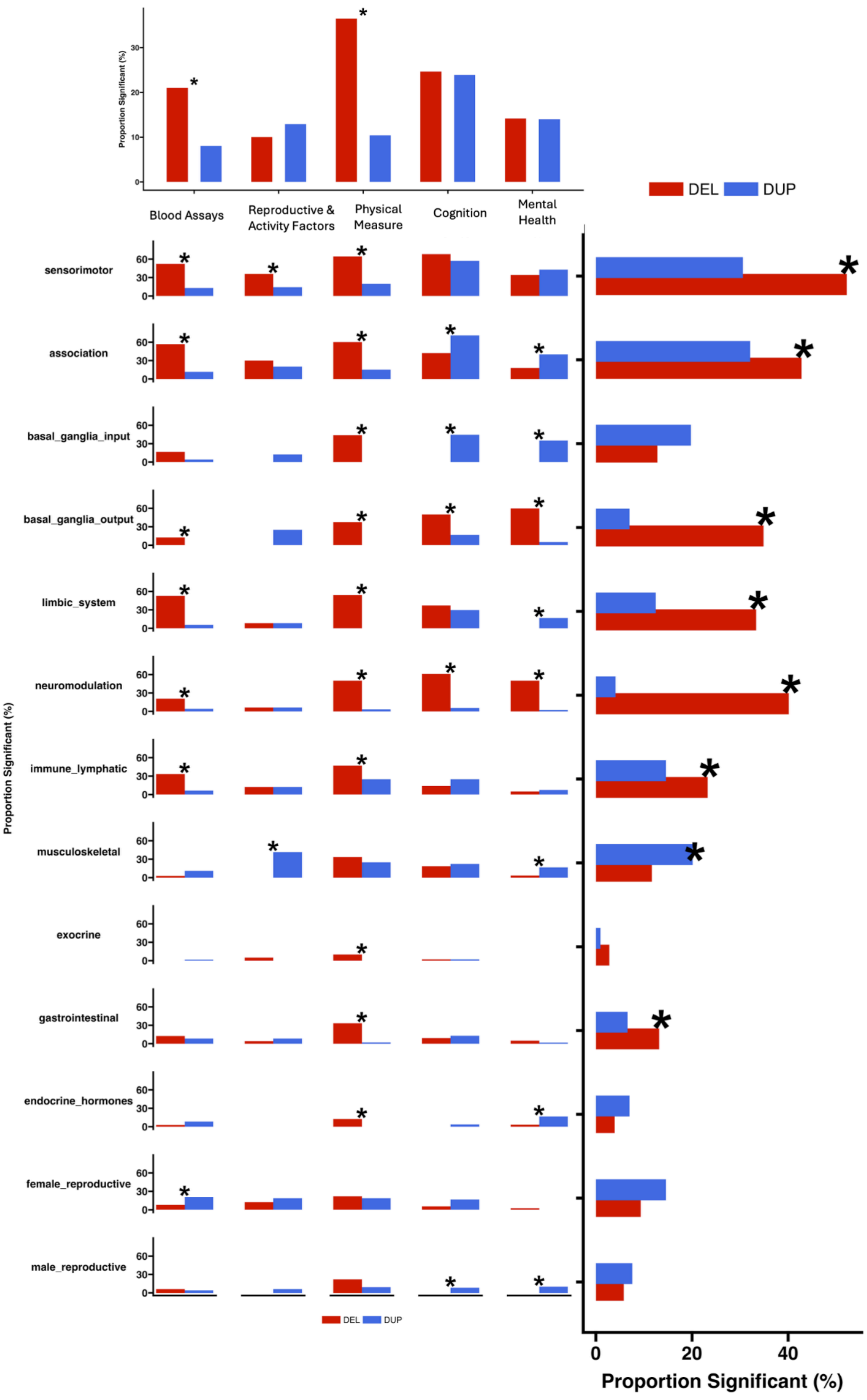


**Supplementary Figure 4: Differences between del-dup associations across trait and gene-set categories**

Bar plots summarizing the differences in the level of association between deletions and duplications (proportion of significant association) for five trait categories and functional gene set categories/annotations. Marginal plots show the differences per trait category (top) or gene-set category (right). Black asterisks (*) indicate statistically significant proportion differences (FDR-adjusted).

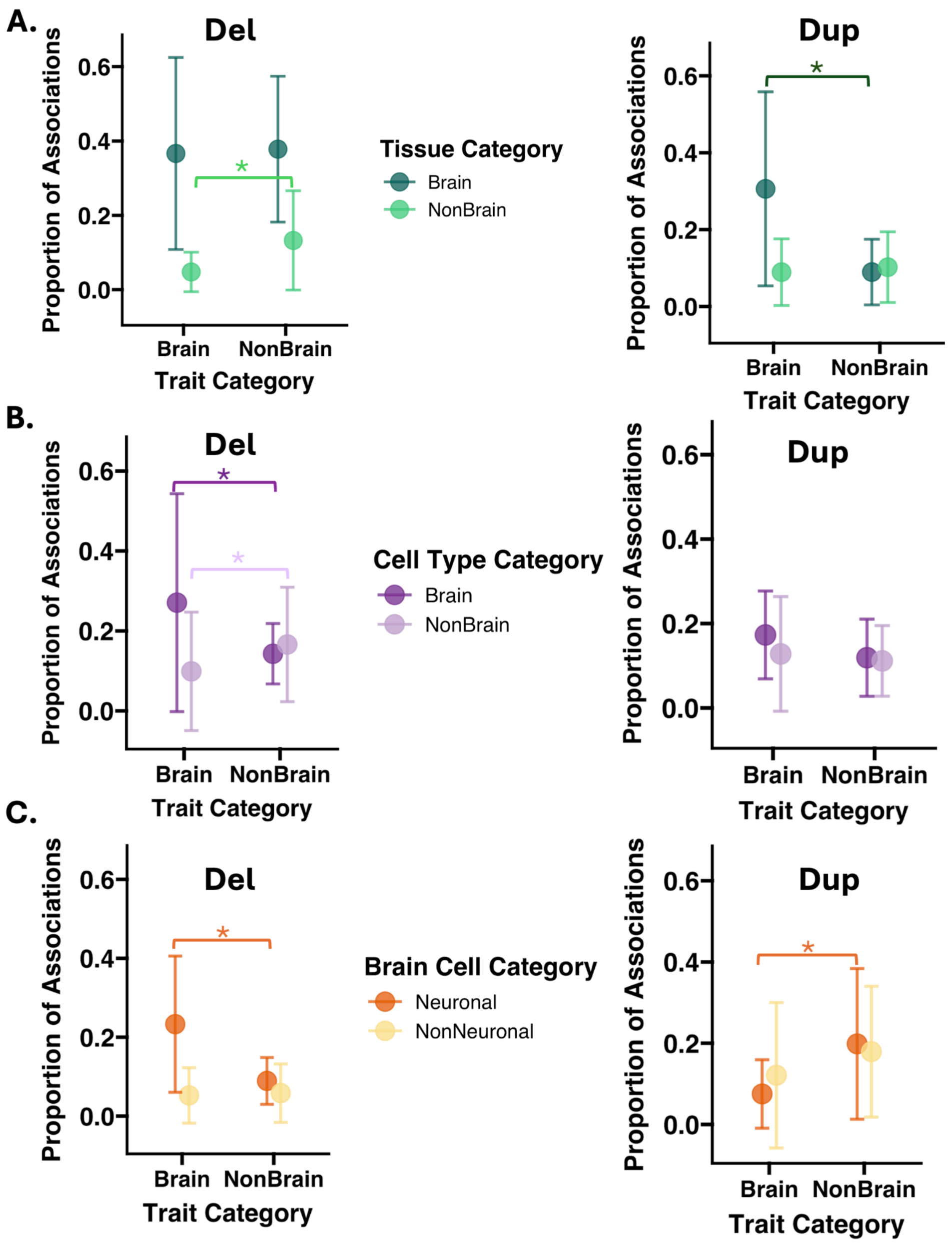


**Supplementary Figure 5: Summary of significant association proportions across tissue- and cell-type gene sets.**

The point range plots in A–C illustrate the proportion of statistically significant associations identified between distinct gene sets (colors) and trait categories (x-axis), and error bars represent the 95% confidence interval. Gene sets are grouped by Tissue (A), Whole-Body Cell Types (B), and Whole-Brain Cell Types (C). An asterisk (*) denotes a statistically significant difference in the association proportion when comparing the two trait categories for a specific gene set, indicating the specificity of the gene set's association to the trait category. Significance was determined using a proportion test after FDR correction (p-value < 0.05).

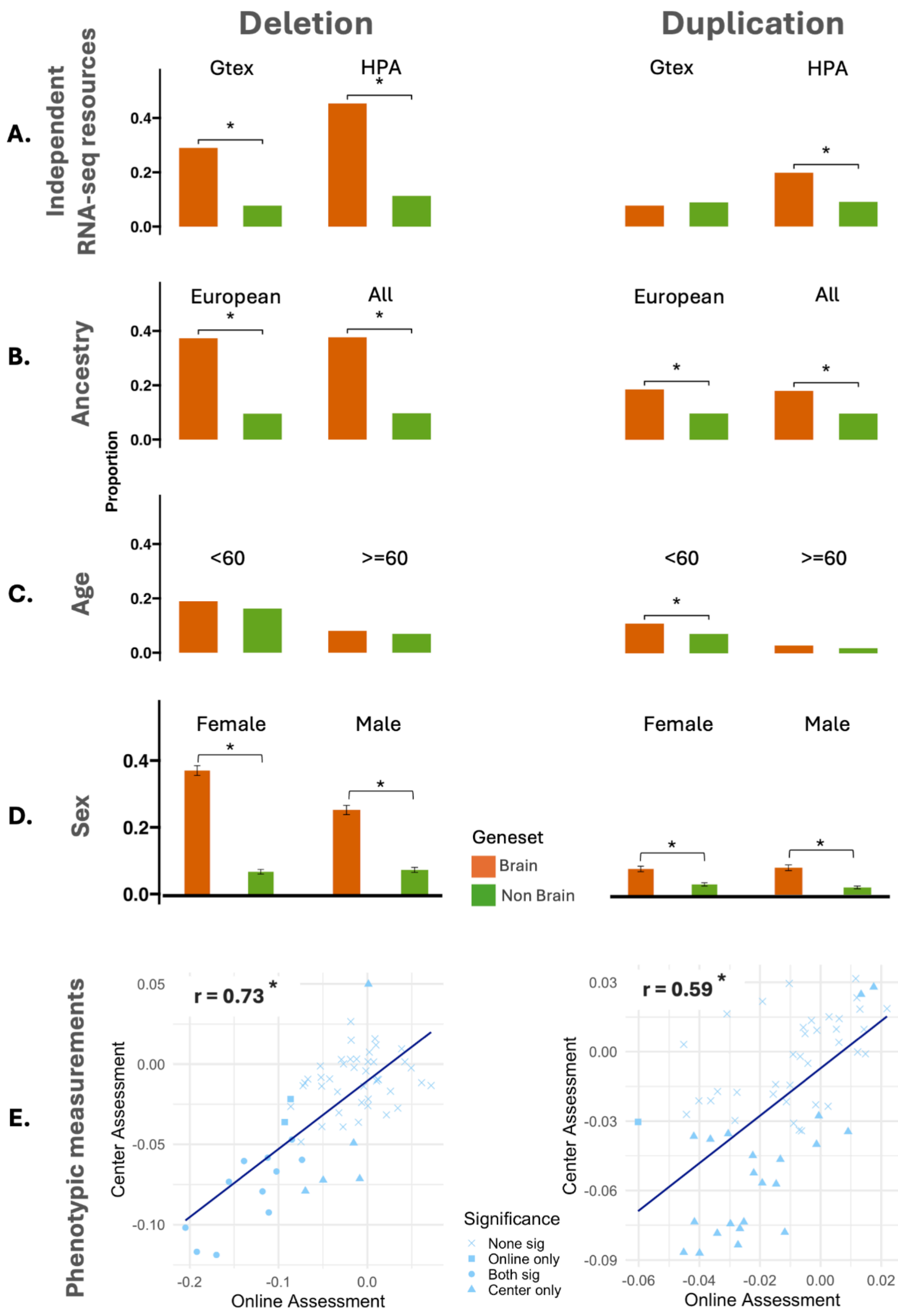


**Supplementary Figure 6: Sensitivity analysis of CNV associations across brain and non-brain gene sets.**

The proportion of significant CNV-trait associations (y-axis, pleiotropy) across brain and non-brain gene sets (x-axis). Each row corresponds to a different sensitivity analysis: (A) independent RNA-seq resources (GTEx vs HPA), (B) ancestry (White vs all ancestries in UK Biobank), (C) age (<60 vs ≥60 years), (D) sex, and (E) phenotype measurement methods (Fluid Intelligence (FI) measured at assessment center, N = 217,809, vs online assessments, N = 92,106).

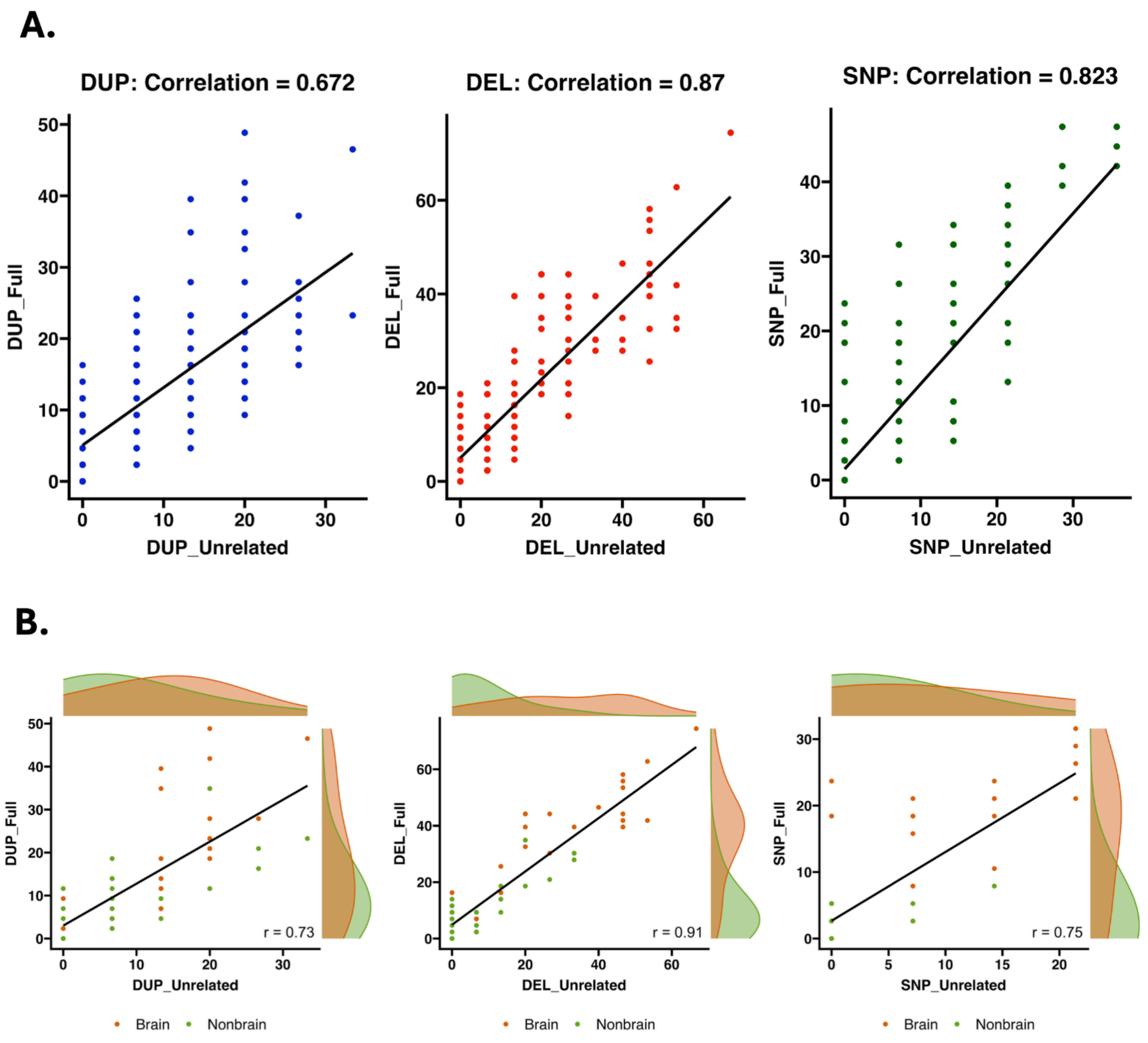


**Supplementary Figure 7: Sensitivity analysis on pleiotropy computed by unrelated and all traits**

X-axis & Y-axis represent the pleiotropy calculated on unrelated and all traits, respectively, using: A) All gene sets (172) and B) Tissue gene sets (60). For pleiotropy on unrelated traits, we recalculated pleiotropy using a subset of 15 unrelated traits that have less than 0.3 correlation with other traits. The pleiotropy estimates from this subset remained highly correlated with those from the full set, with correlations of 68% and 87% for duplications and deletions, respectively. These correlations are higher for tissue gene sets, 73% and 91%, with significant separation between brain and non-brain tissue gene sets.

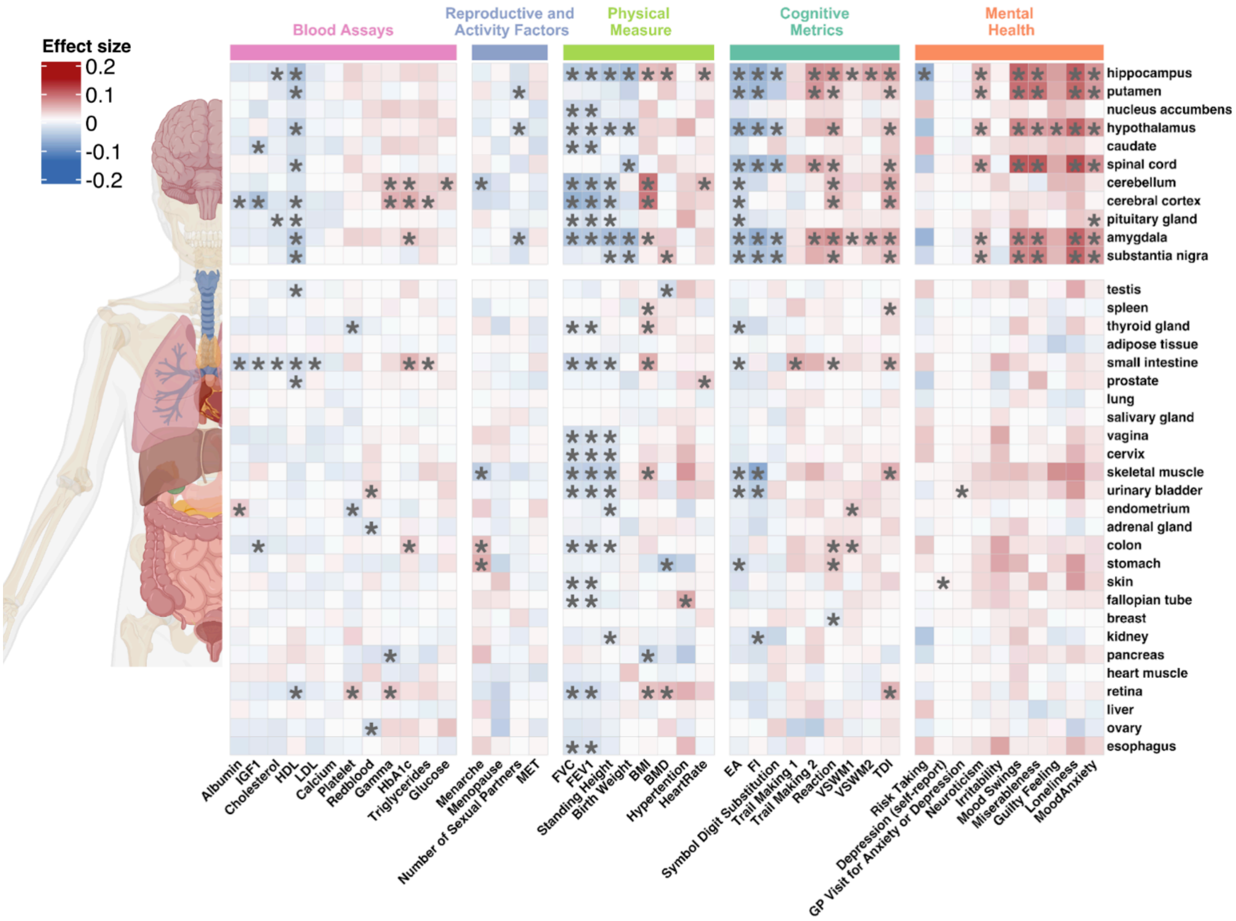


**Supplementary Figure 8: Heatmap of deletion effect sizes for whole-body tissue (GTEx) across traits**

The heatmap displays deletion burden association effect sizes between five categories of traits (x-axis) and GTEx-based tissue-specific gene sets (y-axis). The color intensity reflects the direction and magnitude of the association (blue = negative effect size; red = positive effect size). Black asterisks (*) indicate statistically significant associations between traits and genes (FDR correction across all GTEx gene sets, 43 traits, and 2 CNV types).

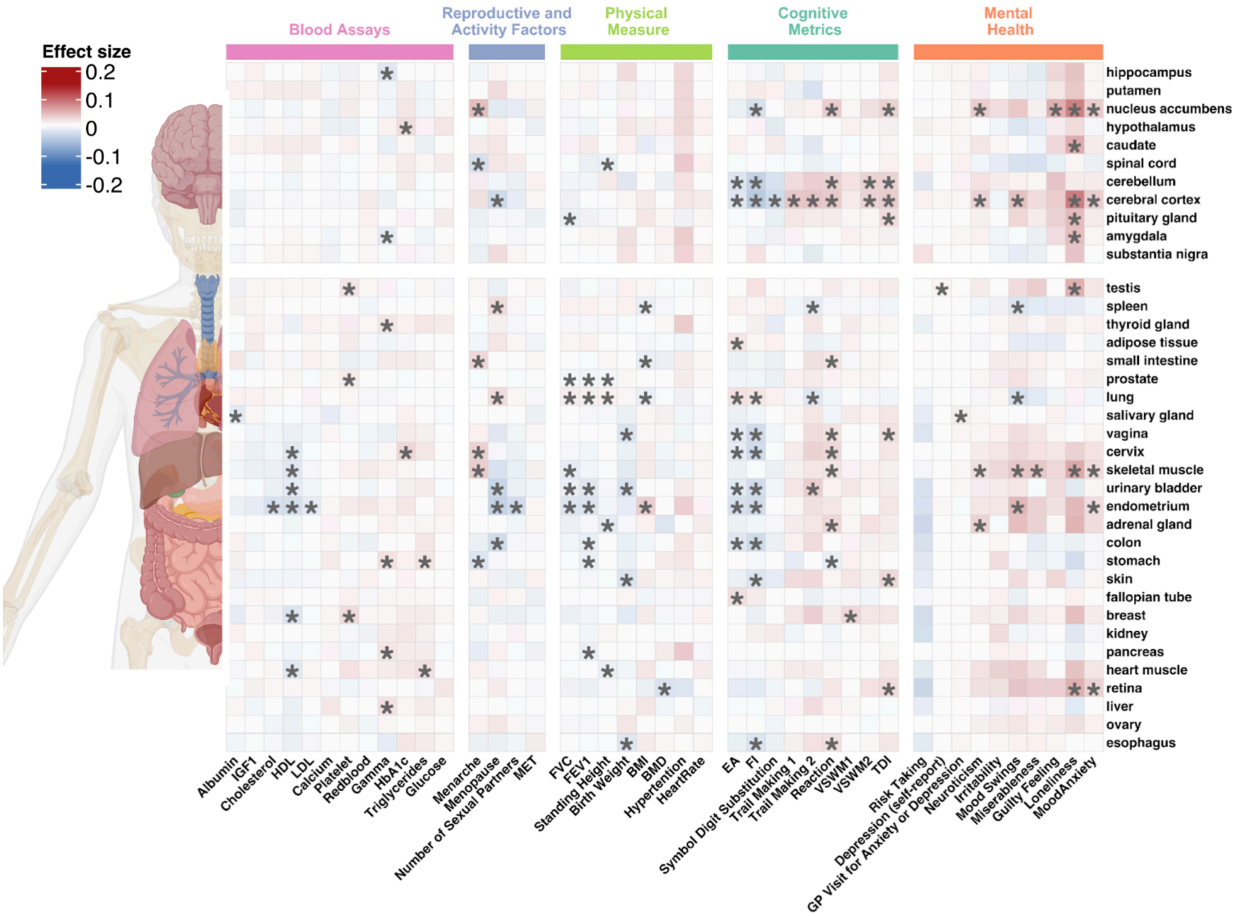


**Supplementary Figure 9: Heatmap of duplication effect sizes for whole-body tissue (GTEx) across traits**

The heatmap displays duplication burden association effect sizes between five categories of traits (x-axis) and GTEx-based tissue-specific gene sets (y-axis). The color intensity reflects the direction and magnitude of the association (blue = negative effect size; red = positive effect size). Black asterisks (*) indicate statistically significant associations between traits and genes (FDR correction across all GTEx gene sets, 43 traits, and 2 CNV types).

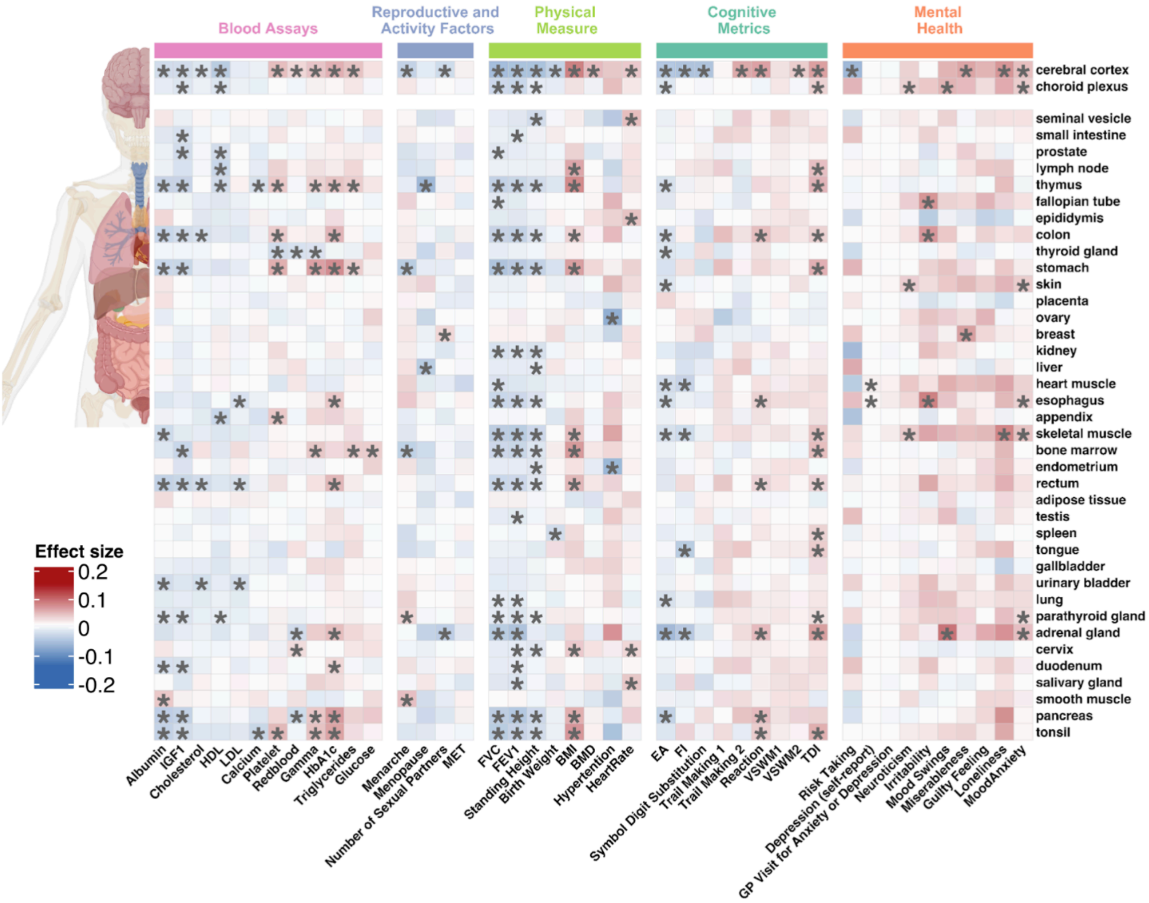


**Supplementary Figure 10: Heatmap of deletion effect sizes for whole-body tissue (HPA) across traits**

The heatmap displays deletion burden association effect sizes between five categories of traits (x-axis) and Human Protein Atlas (HPA) based tissue-specific gene sets (y-axis). The color intensity reflects the direction and magnitude of the association (blue = negative effect size; red = positive effect size). Black asterisks (*) indicate statistically significant associations between traits and genes (FDR correction across all HPA tissue gene sets, 43 traits, and 2 CNV types).

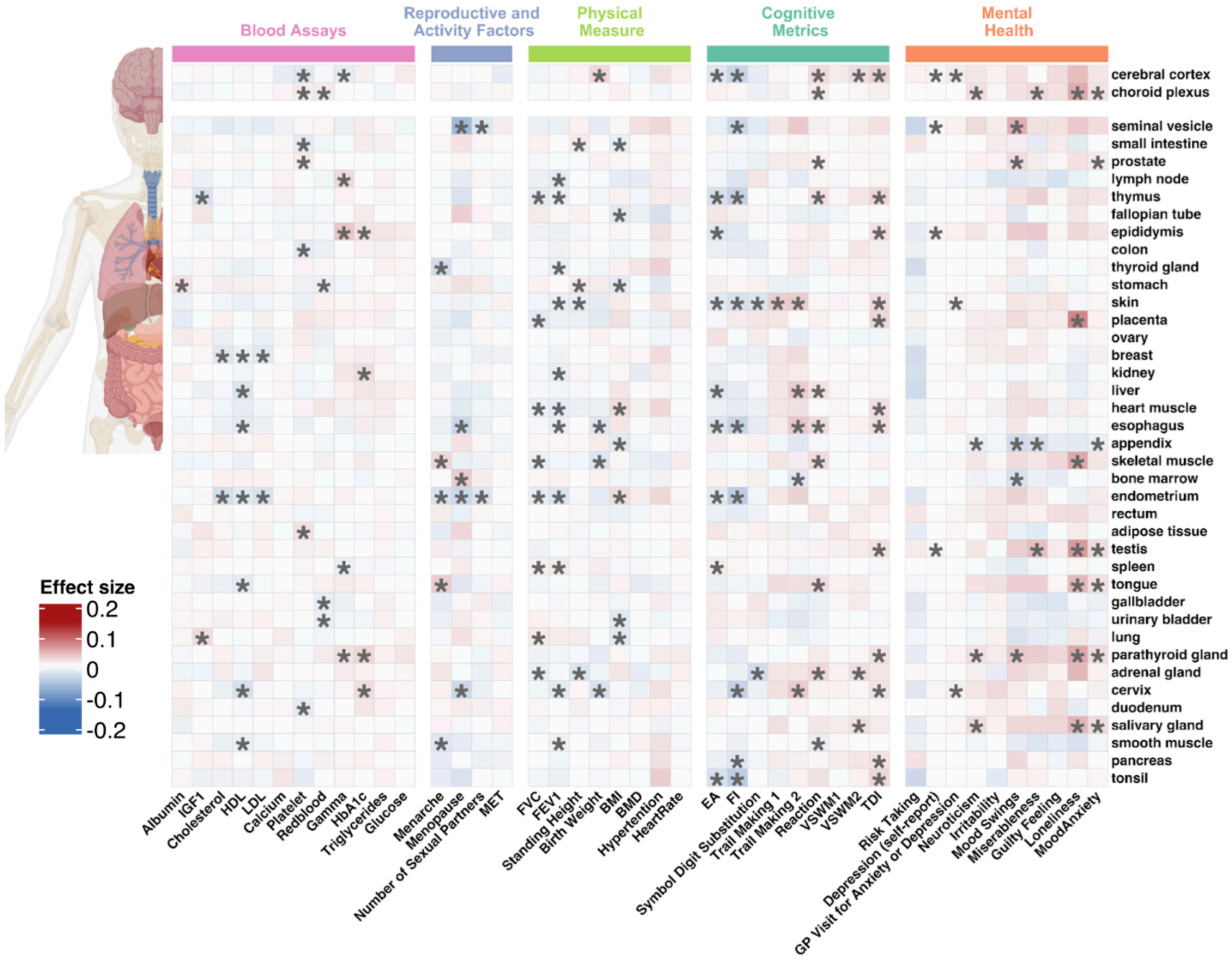


**Supplementary Figure 11: Heatmap of duplication effect sizes for whole-body tissue (HPA) across traits**

The heatmap displays duplication burden association effect sizes between five categories of traits (x-axis) and Human Protein Atlas (HPA) based tissue-specific gene sets (y-axis). The color intensity reflects the direction and magnitude of the association (blue = negative effect size; red = positive effect size). Black asterisks (*) indicate statistically significant associations between traits and genes (FDR correction across all HPA tissue gene sets, 43 traits, and 2 CNV types).

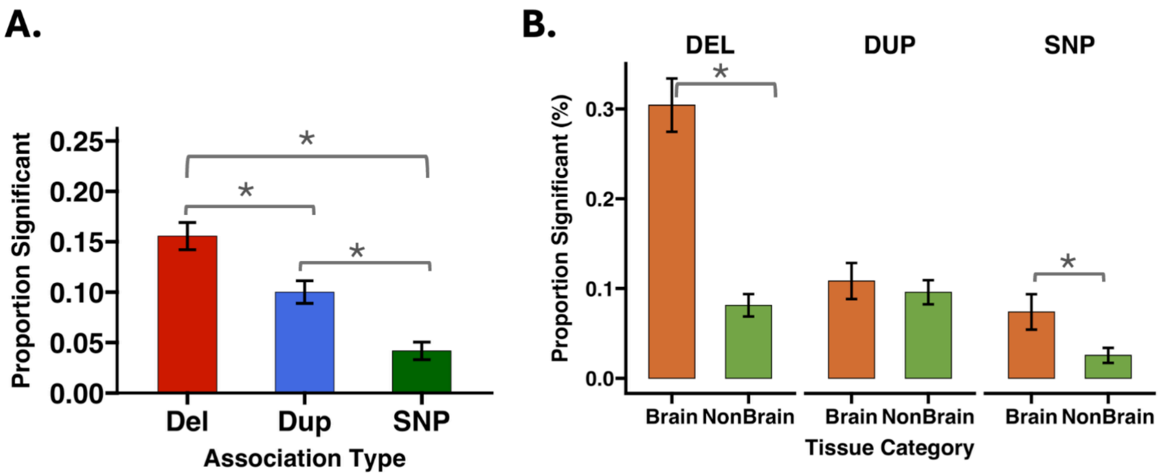


**Supplementary Figure 12: Proportion of significant associations across variant types and tissue categories on independent gene sets and traits**

(A) Proportion of Significant Associations by Variant Type. Bar chart displaying the proportion of significant traits–gene set associations for deletions (Del, red), duplications (Dup, blue), and single nucleotide polymorphisms (SNP, green). (B) Comparison of Significance Proportions Between Brain and Non-Brain Tissue Gene Sets. Proportions are stratified by tissue category (Brain, orange; Non-Brain, green) across the three variant types. To account for non-independence, calculations were performed using a filtered dataset consisting of: (i) gene sets with pairwise Jaccard overlap <= 20% selected via LASSO regularization, and (ii) a subset of 15 independent traits with average pairwise phenotypic correlations < 0.3. Error bars represent the standard error of the proportion; asterisks * indicate significant differences between categories based on two-proportion z-tests.

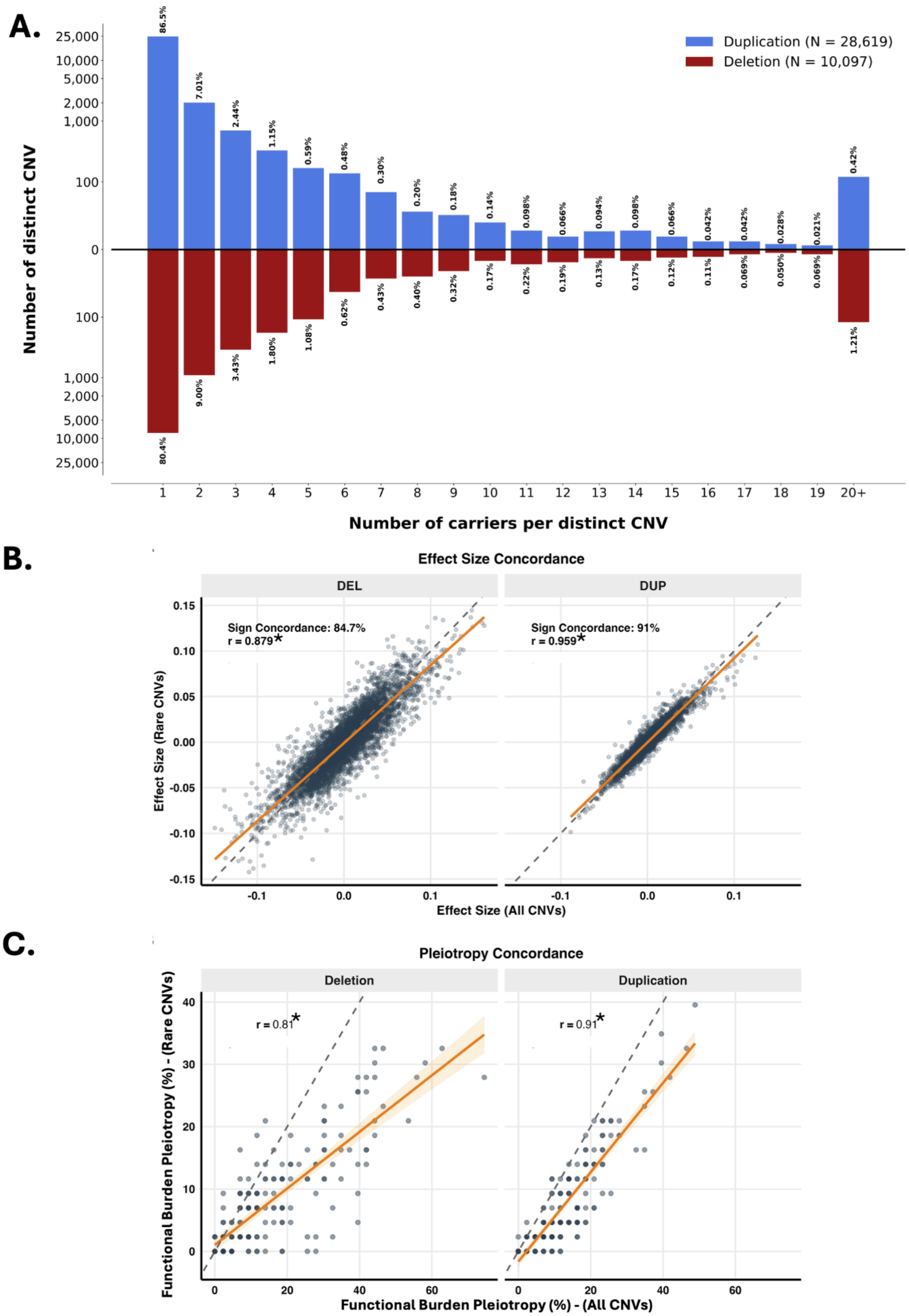


**Supplementary Figure 13: Results are not related to a small number of recurrent CNVs**

(A) Distribution of CNV Recurrence. Mirror bar chart showing the frequency of distinct duplications (N = 28,619, blue) and deletions (N = 10,097, red) based on carrier counts within the full biobank population analyzed for BMI. The majority of observed events are ultra-rare, with only 0.42% of duplications and 1.21% of deletions carried by 20 or more individuals. (B) Trait–Gene Set Effect Size Concordance. Scatter plots comparing the effect size estimates derived from all observed CNVs versus rare CNVs only (excluding recurrent events with >20 carriers). Concordance was evaluated across 43 traits and 172 defined gene sets. High sign concordance (84.7% for DEL; 91% for DUP) and strong Pearson correlations (r = 0.88) indicate that trait–gene set associations are robust to the removal of recurrent structural variants. (C) Preservation of Functional Burden Pleiotropy. Correlation of functional burden pleiotropy rankings (percentage of traits hit) for the 172 gene sets across the 43 traits. Strong correlation coefficients (r = 0.81 for DEL; r = 0.91 for DUP) and significant Jaccard-based overlaps ($p < 0.05$) confirm that functional burden pleiotropy is driven by a distributed genic CNV burden rather than the influence of a small number of recurrent events. Orange lines represent linear regression fits with 95% confidence intervals; dashed lines indicate the identity (y = x) slope.

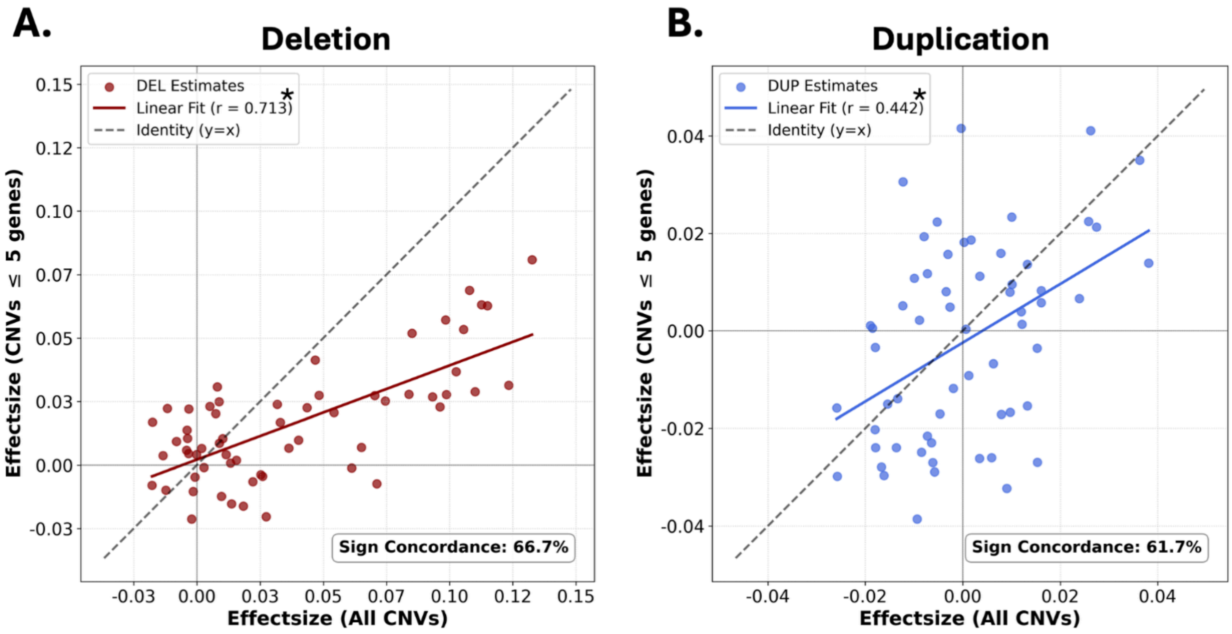


**Supplementary Figure 14: Excluding large multigenic CNVs (≥5 Genes) shows concordant effect size profiles**

Scatter plots comparing the effect size estimates (beta) derived from all CNVs versus CNVs encompassing fewer than 5 protein-coding genes, for (A) Deletions and (B) Duplications, across Fantom tissue gene sets, for BMI. The x-axis represents the effect sizes derived from the primary model, including all CNVs, while the y-axis represents the effect sizes after excluding large multigenic CNVs (those encompassing >5 genes). The dashed line represents the line of perfect concordance (y=x). The solid lines indicate the linear regression line of best fit, with Pearson correlation coefficients (r) provided in the legend (* indicates r p-value < 0.05). Text boxes in the lower right indicate the overall sign concordance, representing the percentage of gene sets that maintained the same direction of effect across both models. The positive correlations and high sign concordance demonstrate that the observed associations are robust and not solely driven by large multigenic CNVs.

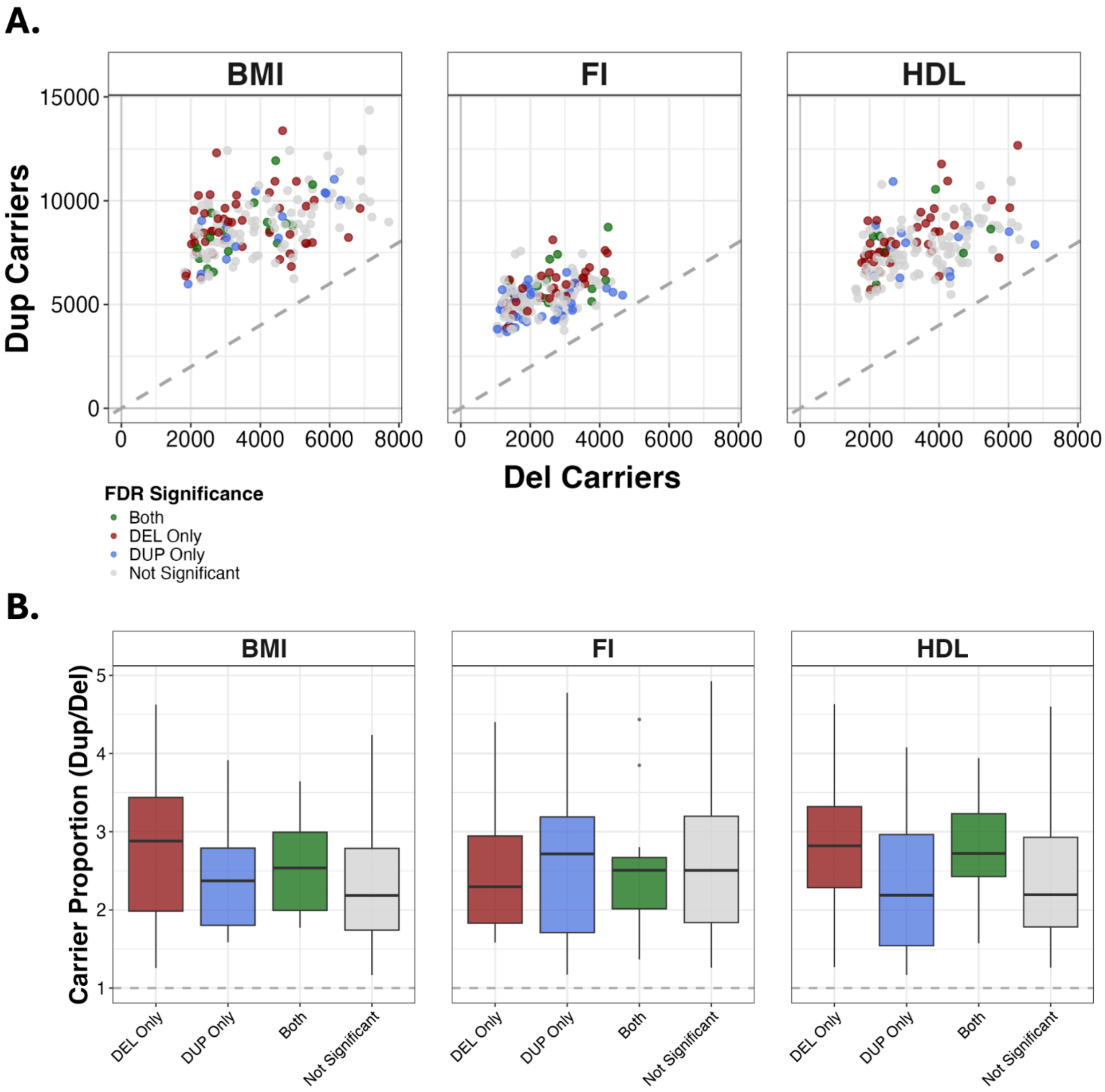

**Supplementary Figure 15: Specific effects of deletions and duplications are not explained by power imbalance**

(A) Absolute duplication carriers (y-axis) versus deletion carriers (x-axis) counts across 172 gene sets. For three traits spanning distinct biological domains (BMI, Fluid Intelligence, HDL cholesterol), we compared the ratio of duplication to deletion carriers across gene sets (172 gene sets), stratified by significance category: deletion-only, duplication-only, both significant, or neither. Across all categories, duplication carriers were 2- to 3-fold more frequent than deletion carriers. This means that a gene set reaching significance for deletions only versus duplications only is not predicted by the relative number of carriers. Points are colored by FDR significance category: both significant (green), deletion only (red), duplication only (blue), or neither (grey). Dashed line: identity (y = x). (B) To formally demonstrate the absence of power imbalance, we represent boxplots of the ratio of duplication to deletion carriers across significant trait-gene set associations stratified by significance category: deletion-only, duplication-only, both significant, or neither. The dup/del carrier ratio did not differ between deletion-only and duplication-only gene sets for any trait, nor between any pair of significance categories (Wilcoxon tests, all non-significant after correction). The median ratio consistently ranges between 2 and 3 across all categories.

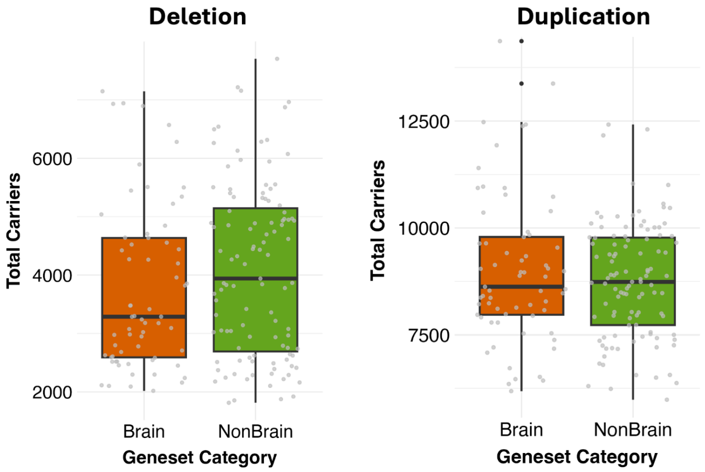


**Supplementary Figure 16: No differences in the number of carriers between the brain and non-brain gene sets**

Boxplots show the total number of CNV carriers for deletions (left) and duplications (right) across brain-specific and non-brain gene sets, using body mass index (BMI) as a representative trait. Each point represents a specific gene set. There was no significant difference in carrier frequency between brain and non-brain categories for either deletions (Wilcoxon $p = 0.15$) or duplications (Wilcoxon $p = 0.69$). Center lines represent the median; box limits indicate the 25th and 75th percentiles. These suggest that the observed higher functional pleiotropy in brain-specific gene sets is not driven by differences in CNV frequency or statistical power.

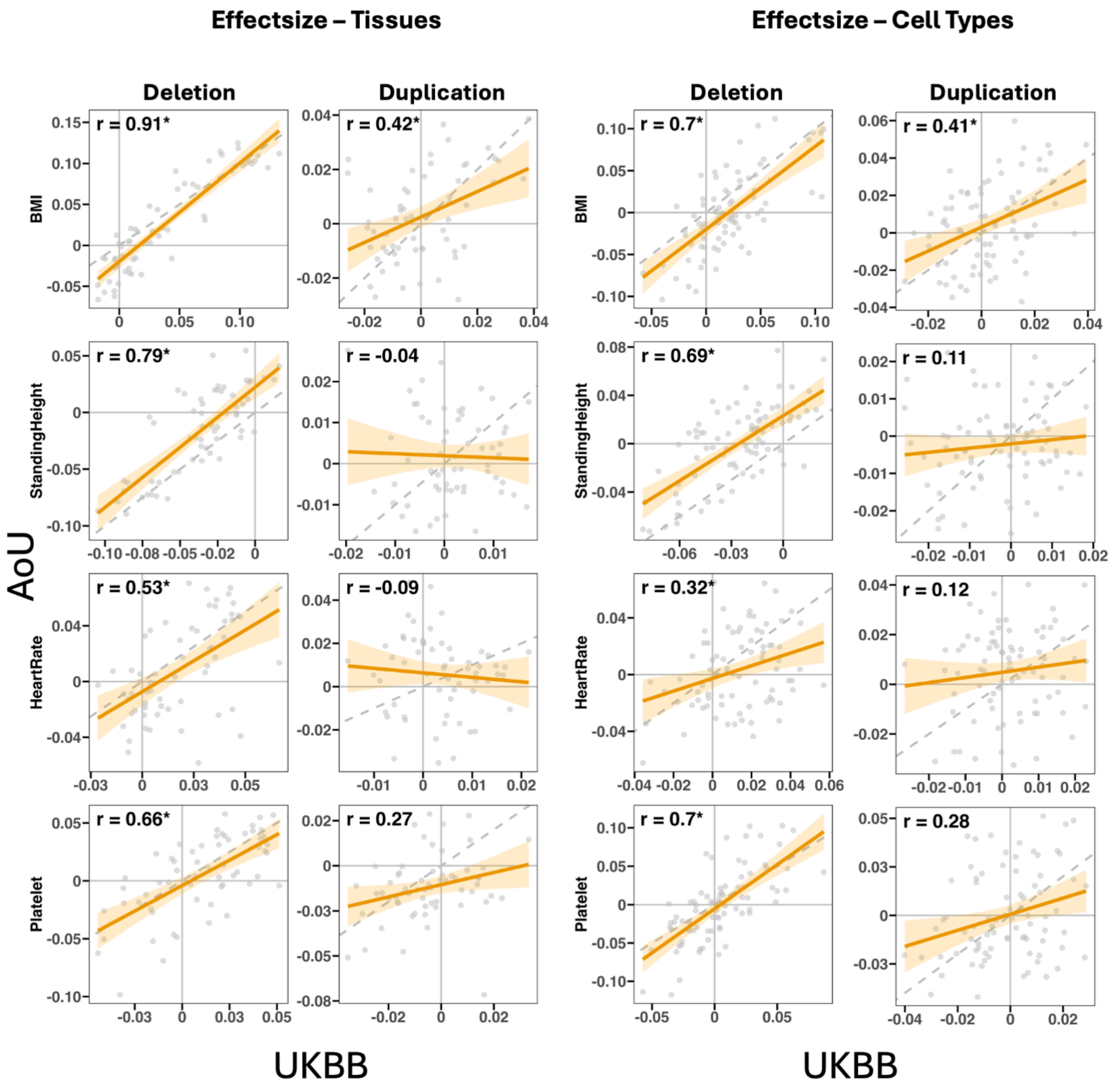


**Supplementary Figure 17: Replication of tissue-specific CNV burden associations in All of Us (AoU)**

Scatter plots compare effect size estimates (Beta) between UKBB and AoU across 60 whole body tissue (left panel) and 81 whole body cell type (right panel) gene sets for four replication traits (BMI, Heart Rate, Platelet Count, Standing Height). Pearson correlations (r) denote the correlation between biobanks for deletions (left) and duplications (right). Orange lines and shaded areas represent linear regression fits with 95% confidence intervals; dashed lines indicate the identity (y = x) slope.

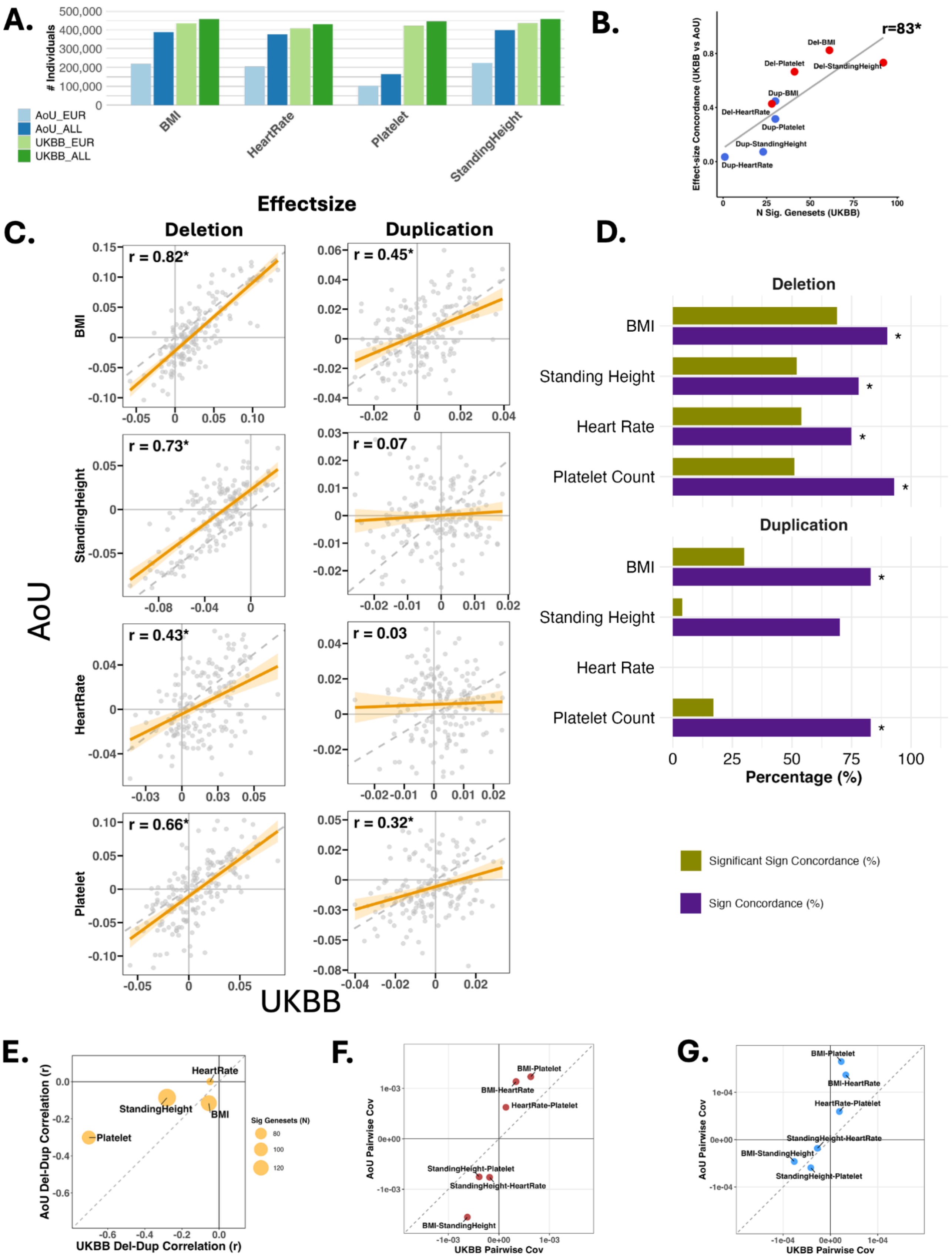
A.
### Individuals
500,000
400,000
300,000
200,000
100,000
0
AoU_EUR
AoU_ALL
UKBB_EUR
UKBB_ALL
BMI
HeartRate
Platelet
StandingHeight
B.
Effect-size Concordance (UKBB vs AoU)
N Sig. Genesets (UKBB)
r=83*
Del-BMI
Del-Platelet
Del-StandingHeight
Dup-BMI
Del-HeartRate
Dup-Platelet
Dup-StandingHeight
Dup-HeartRate
Effectsize
C.
Deletion
Duplication
BMI
StandingHeight
HeartRate
Platelet
r = 0.82*
r = 0.45*
r = 0.73*
r = 0.07
r = 0.43*
r = 0.03
r = 0.66*
r = 0.32*
AoU
UKBB
D.
Deletion
Duplication
BMI
Standing Height
Heart Rate
Platelet Count
Percentage (%)
Significant Sign Concordance (%)
Sign Concordance (%)
E.
AoU Del-Dup Correlation (r)
UKBB Del-Dup Correlation (r)
HeartRate
StandingHeight
BMI
Platelet
Sig Genesets (N)
80
100
120
F.
AoU Pairwise Cov
UKBB Pairwise Cov
BMI-Platelet
BMI-HeartRate
HeartRate-Platelet
StandingHeight-Platelet
StandingHeight-HeartRate
BMI-StandingHeight
G.
AoU Pairwise Cov
UKBB Pairwise Cov
BMI-Platelet
BMI-HeartRate
HeartRate-Platelet
StandingHeight-HeartRate
BMI-StandingHeight
StandingHeight-Platelet

**Supplementary Figure 18: Replication of CNV burden associations in the All of Us (AoU) across 172 gene sets**

(A) Sample sizes. Number of individuals with genotyping and phenotypic data in AoU and UK Biobank (UKBB) for four replication traits (BMI, Heart Rate, Platelet Count, Standing Height), stratified by European-specific (EUR) and all-ancestry (ALL) groupings. Replication analyses used European-ancestry participants only to match the main UKBB analyses. (B) Replication strength vs. discovery power. Per-trait cross-cohort concordance of gene-set effect sizes (Pearson r) plotted against the number of FDR-significant gene sets in UKBB. Bubble size reflects the number of FDR-significant gene sets in UKBB. (C) Gene-set effect size concordance. Comparison of 172 gene-set effect sizes (Beta) between UKBB (x-axis) and AoU (y-axis), for deletions and duplications across the four traits. Pearson r is indicated per comparison; asterisks (*) denote FDR significance. (D) Directional replication of FDR-significant associations. For each gene-set association that reached FDR significance in UKBB, we assessed whether the effect was in the same direction in AoU (sign concordance), and among those, whether the AoU association also reached nominal significance ($p < 0.05$; significant sign concordance) — representing increasingly stringent levels of replication. Under a null model with no true signal, 50% sign concordance would be expected by chance. Sign concordance (purple bars) and significant sign concordance (olive bars) are shown separately for deletions (top) and duplications (bottom) across the four traits. Asterisks denote FDR-corrected binomial test significance for sign concordance exceeding the 50% null. Across all 172 gene sets, deletion associations showed 84% sign concordance (186/222; binomial p = 7e-26) with 57% reaching nominal significance in the same direction; duplication associations showed 79% sign concordance (66/84; p = 7e-8) with 18% reaching nominal significance. Full per-trait and per-gene-set-category replication statistics are provided in **Table ST9**. (E) Deletion–duplication correlation concordance. Per-trait correlation between deletion and duplication effects across the 172 gene sets in AoU (y-axis) versus UKBB (x-axis). Bubble size reflects the number of FDR-significant gene sets in UKBB. (F–G) Between-trait CNV burden correlation concordance. Pairwise between-trait covariance from (F) deletion (red) and (G) duplication (blue) effect sizes, UKBB (x-axis) versus AoU (y-axis), across all 172 gene sets. Orange lines: linear regression fits with 95% CI. Dashed lines: identity (y = x).

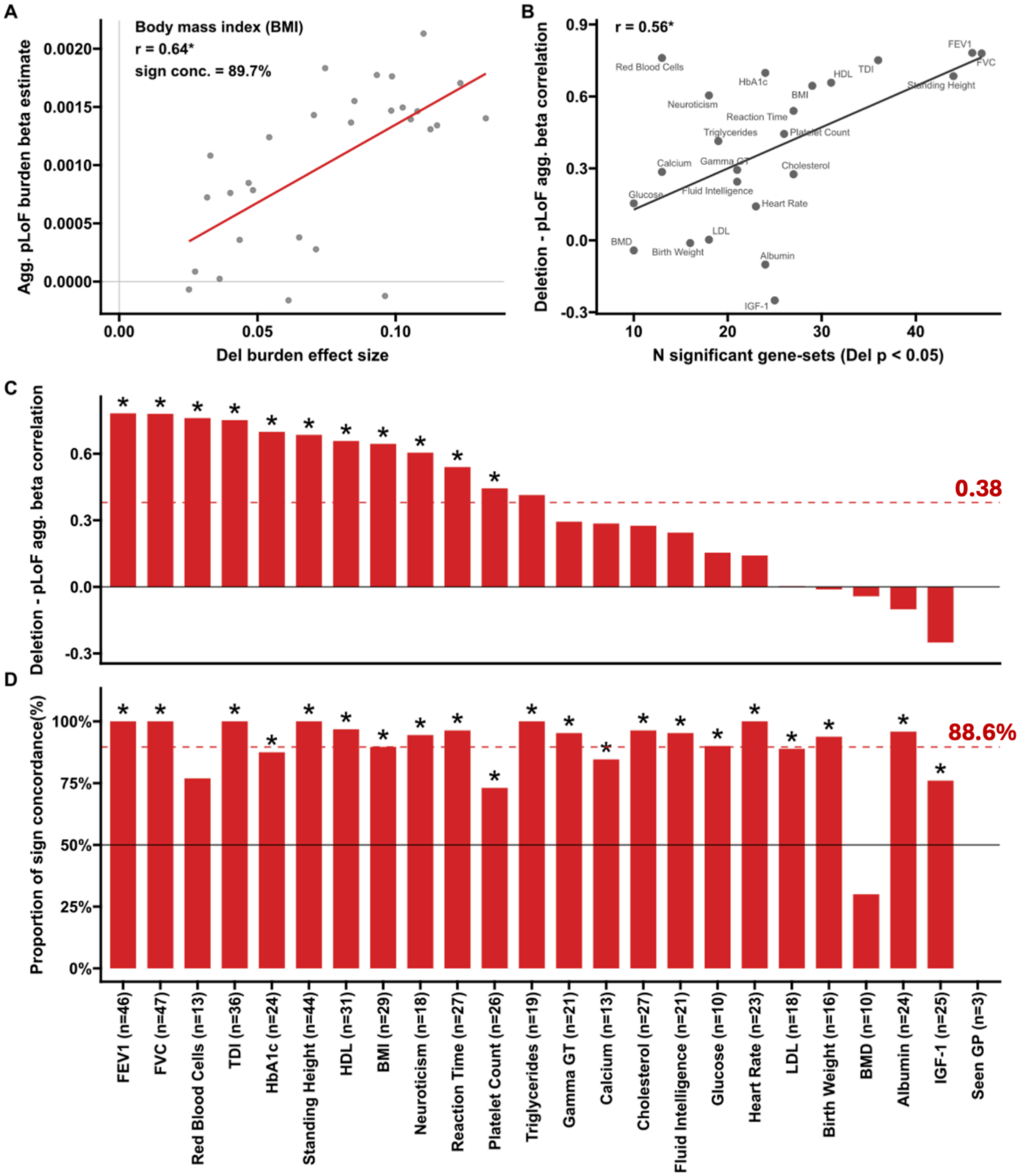


**Supplementary Figure 19: Concordance between CNV deletion burden and aggregated pLoF burden effect sizes (tissue gene-sets)**

(A) CNV burden (deletion) vs aggregated pLoF burden beta estimates for an example trait (Body mass index, BMI) across nominally significant gene-sets (Del p < 0.05). Red line: linear fit; * p-Jaccard < 0.05. Pearson correlation and sign concordance (sign conc.) percentage are reported in the top left corner. (B) Per-trait Del-pLoF correlation as a function of the number of nominally significant gene-sets per trait (24 available traits with Del Burden and pLoF Beta burden). (C) Del-pLoF Pearson correlations across 24 traits, ordered by decreasing r. Parentheses indicate the number of gene-sets used per trait (minimum 6 required, 10% of tissue gene sets). Dashed red line: mean across traits; * FDR corrected p-Jaccard < 0.05. (D) Sign concordance (proportion of gene-sets with same-sign Del and pLoF effects) for the same traits. Black line: 50% chance level; dashed red line: mean across traits; * FDR < 0.05 (binomial test, 24 tests).

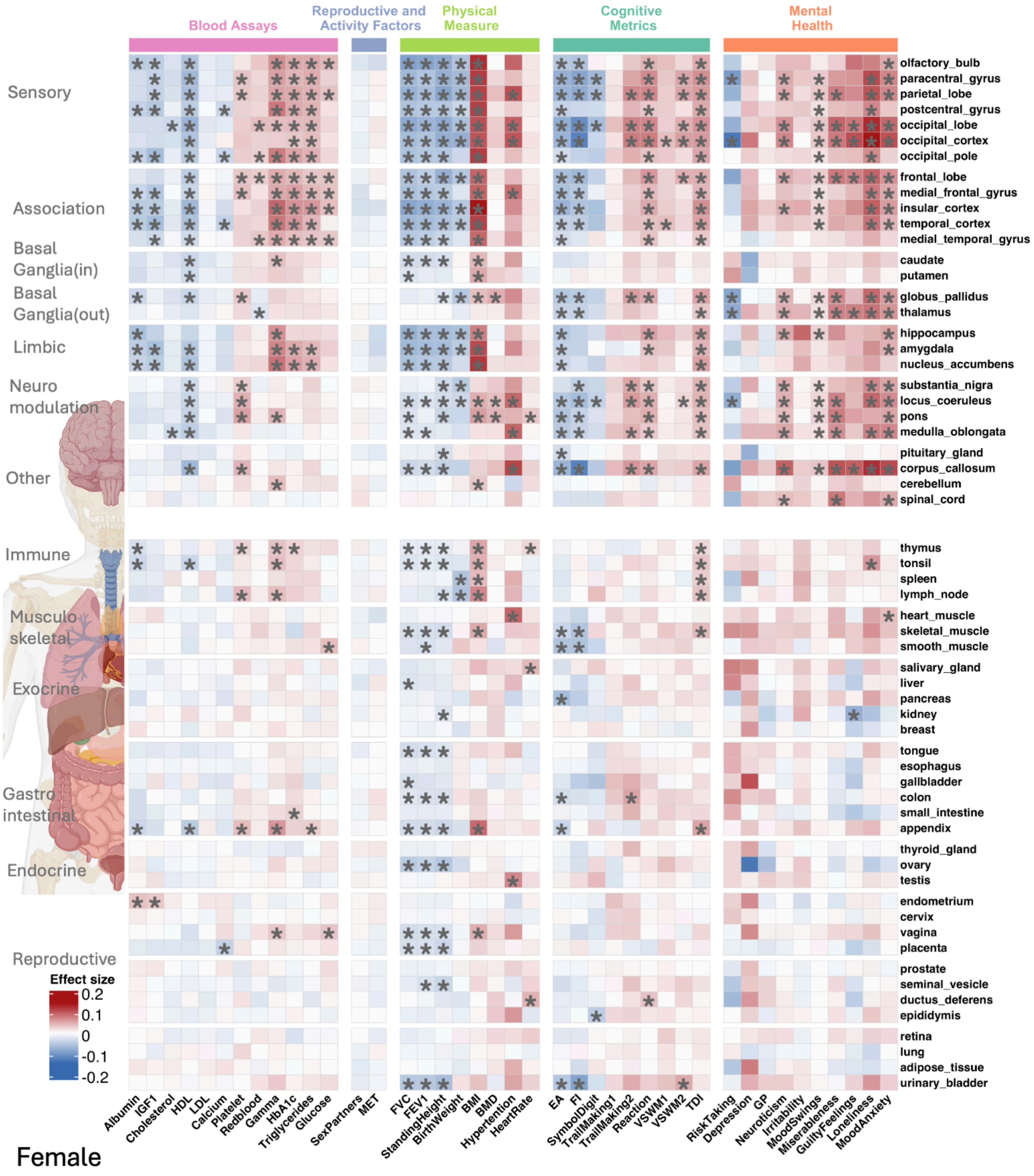


**Supplementary Figure 20: Heatmap of deletion effect sizes for whole-body tissue across traits for Female**

The heatmap displays deletion burden association effect sizes between five categories of traits (x-axis) and tissue-specific gene sets (y-axis) for sex stratified analysis (Female). The color intensity reflects the direction and magnitude of the association (blue = negative effect size; red

= positive effect size). Black asterisks (*) indicate statistically significant associations between traits and genes (FDR correction across all 172 gene sets, 43 traits, and 2 CNV types for Female).

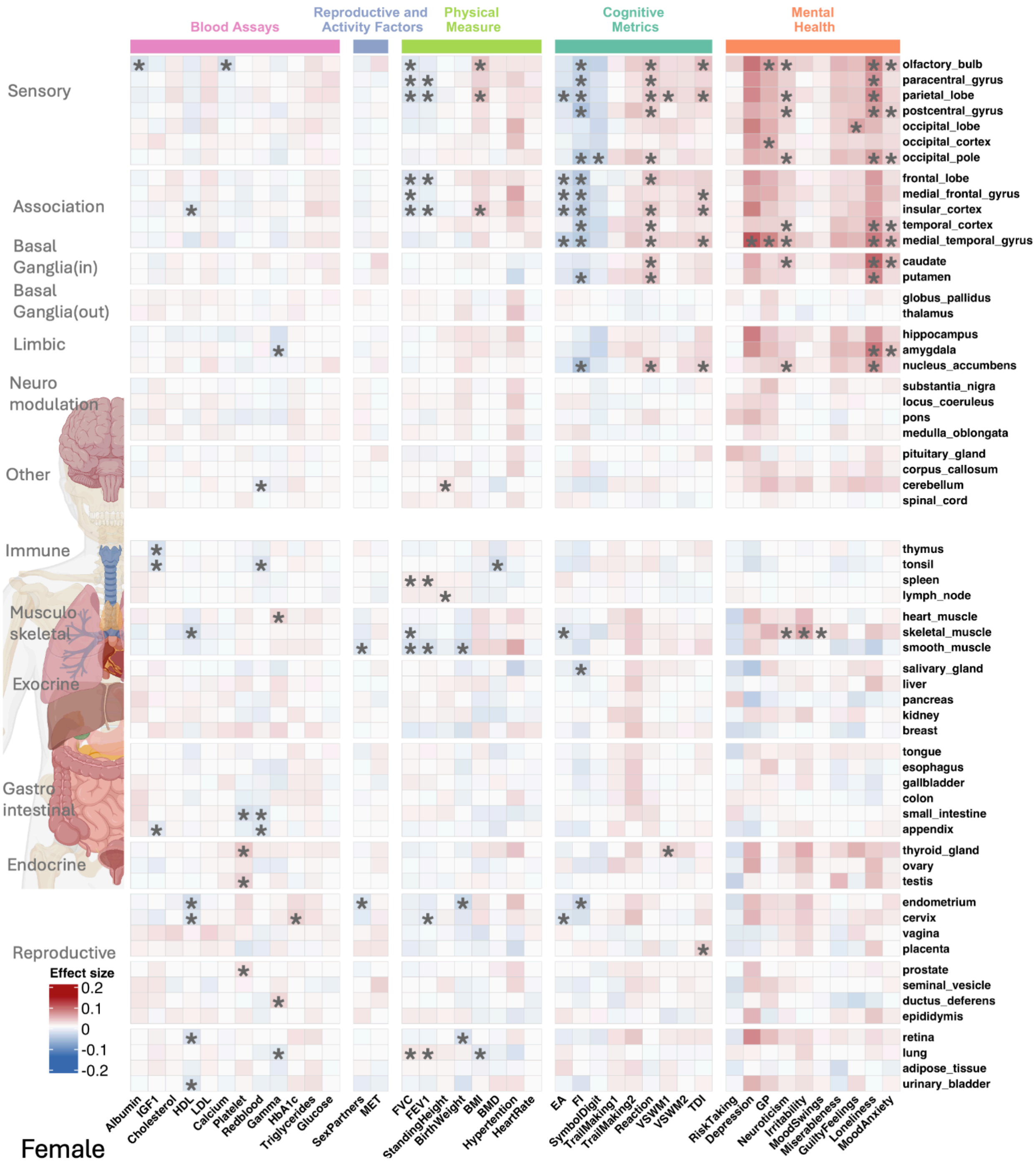


**Supplementary Figure 21: Heatmap of duplication effect sizes for whole-body tissue across traits for Female**

The heatmap displays duplication burden association effect sizes between five categories of traits (x-axis) and tissue-specific gene sets (y-axis) for sex stratified analysis (Female). The color intensity reflects the direction and magnitude of the association (blue = negative effect size; red = positive effect size). Black asterisks (*) indicate statistically significant associations between traits and genes (FDR correction across all 172 gene sets, 43 traits, and 2 CNV types for Female).

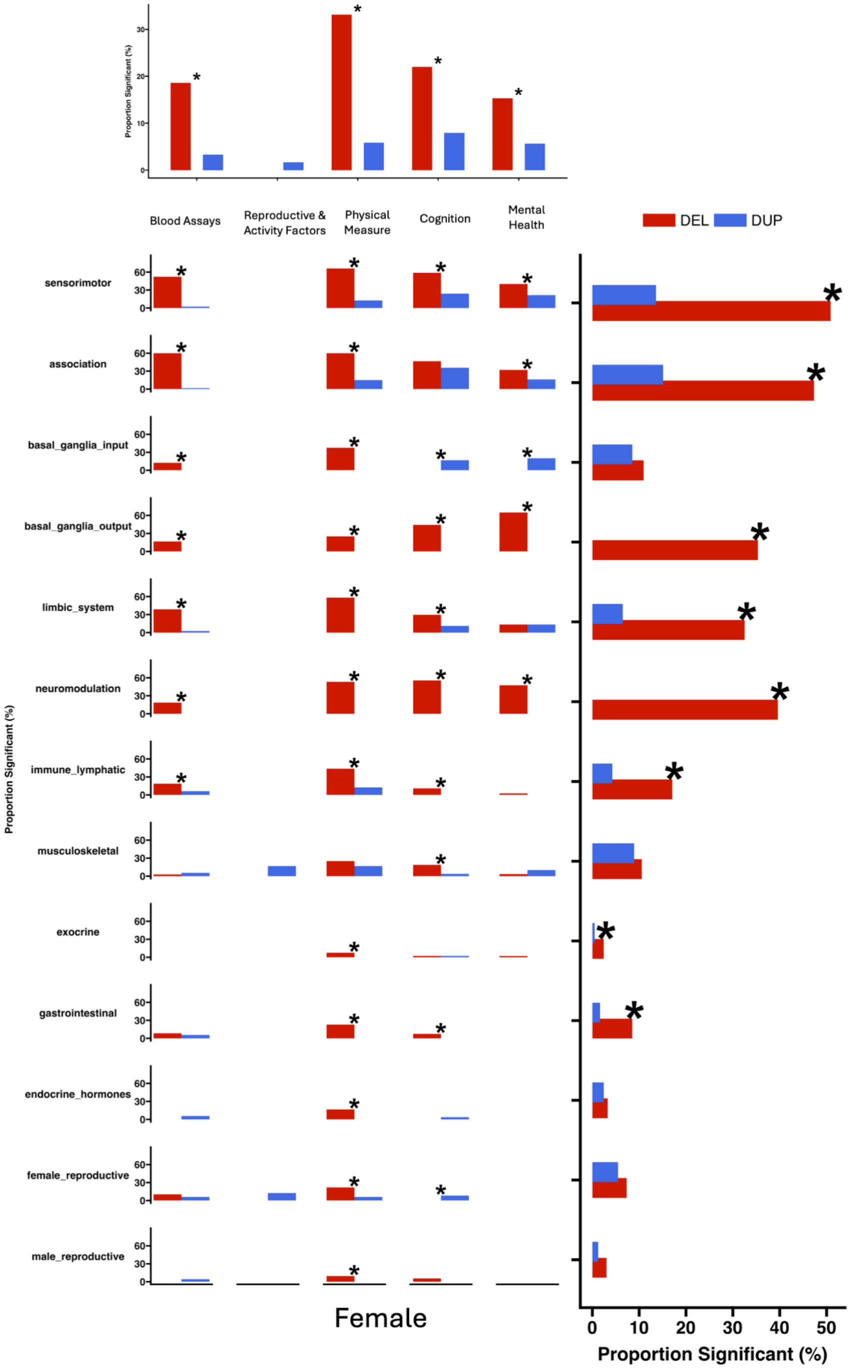


**Supplementary Figure 22: Differences between del-dup associations across trait and gene-set categories for Female**

Bar plots summarizing the differences in the level of association between deletions and duplications (proportion of significant association) for five trait categories and functional gene set categories/annotations for sex stratified analysis (Female). Marginal plots show the differences per trait category (top) or gene-set category (right). Black asterisks (*) indicate statistically significant proportion differences (FDR-adjusted).

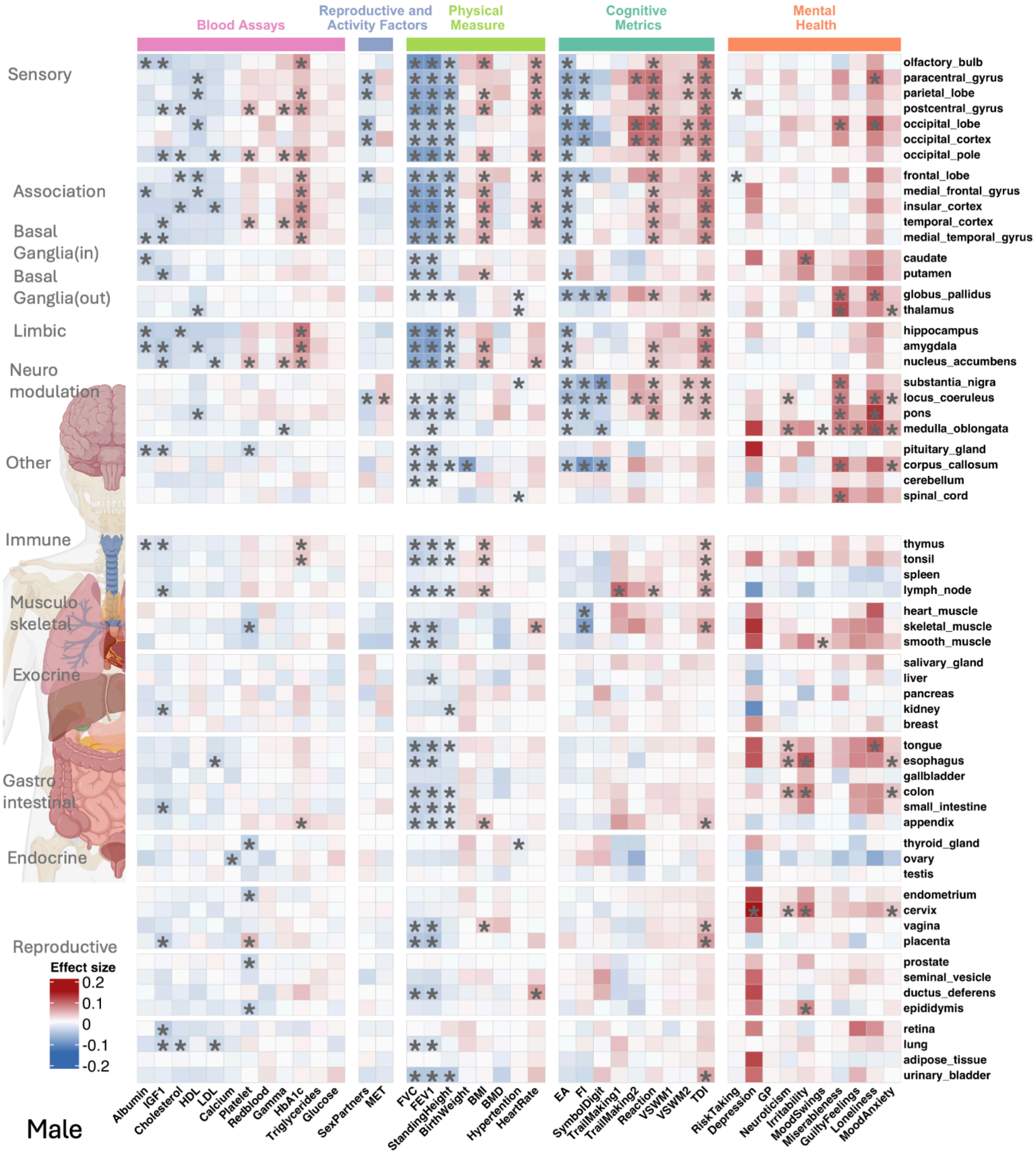


**Supplementary Figure 23: Heatmap of deletion effect sizes for whole-body tissue across traits for Male**

The heatmap displays deletion burden association effect sizes between five categories of traits (x-axis) and tissue-specific gene sets (y-axis) for sex stratified analysis (Male). The color intensity reflects the direction and magnitude of the association (blue = negative effect size; red = positive effect size). Black asterisks (*) indicate statistically significant associations between traits and genes (FDR correction across all 172 gene sets, 43 traits, and CNV type for Male).

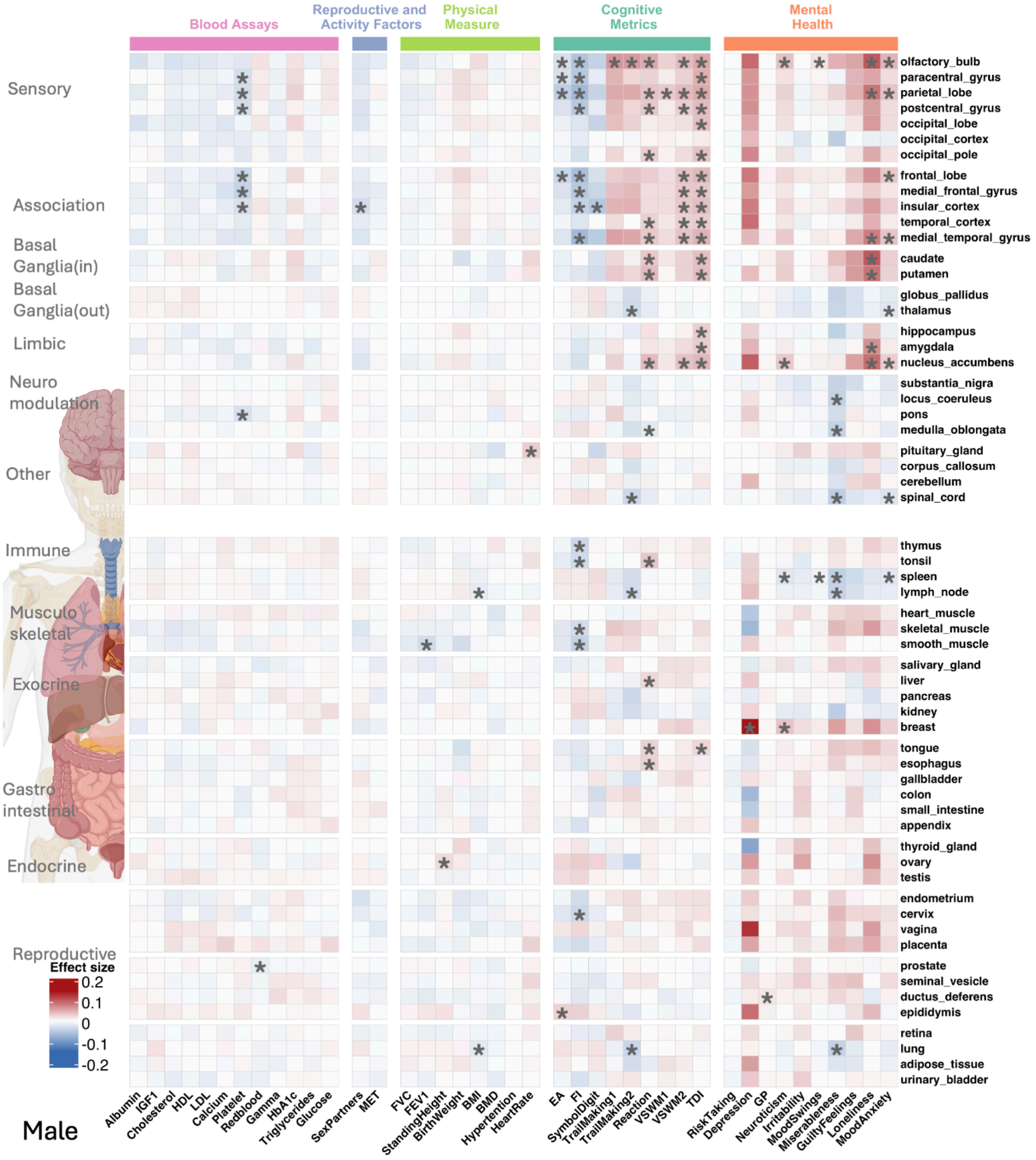


**Supplementary Figure 24: Heatmap of duplication effect sizes for whole-body tissue across traits for Male**

The heatmap displays duplication burden association effect sizes between five categories of traits (x-axis) and tissue-specific gene sets (y-axis) for sex stratified analysis (Male). The color intensity reflects the direction and magnitude of the association (blue = negative effect size; red = positive effect size). Black asterisks (*) indicate statistically significant associations between traits and genes (FDR correction across all 172 gene sets, 43 traits, and 2 CNV types for Male).

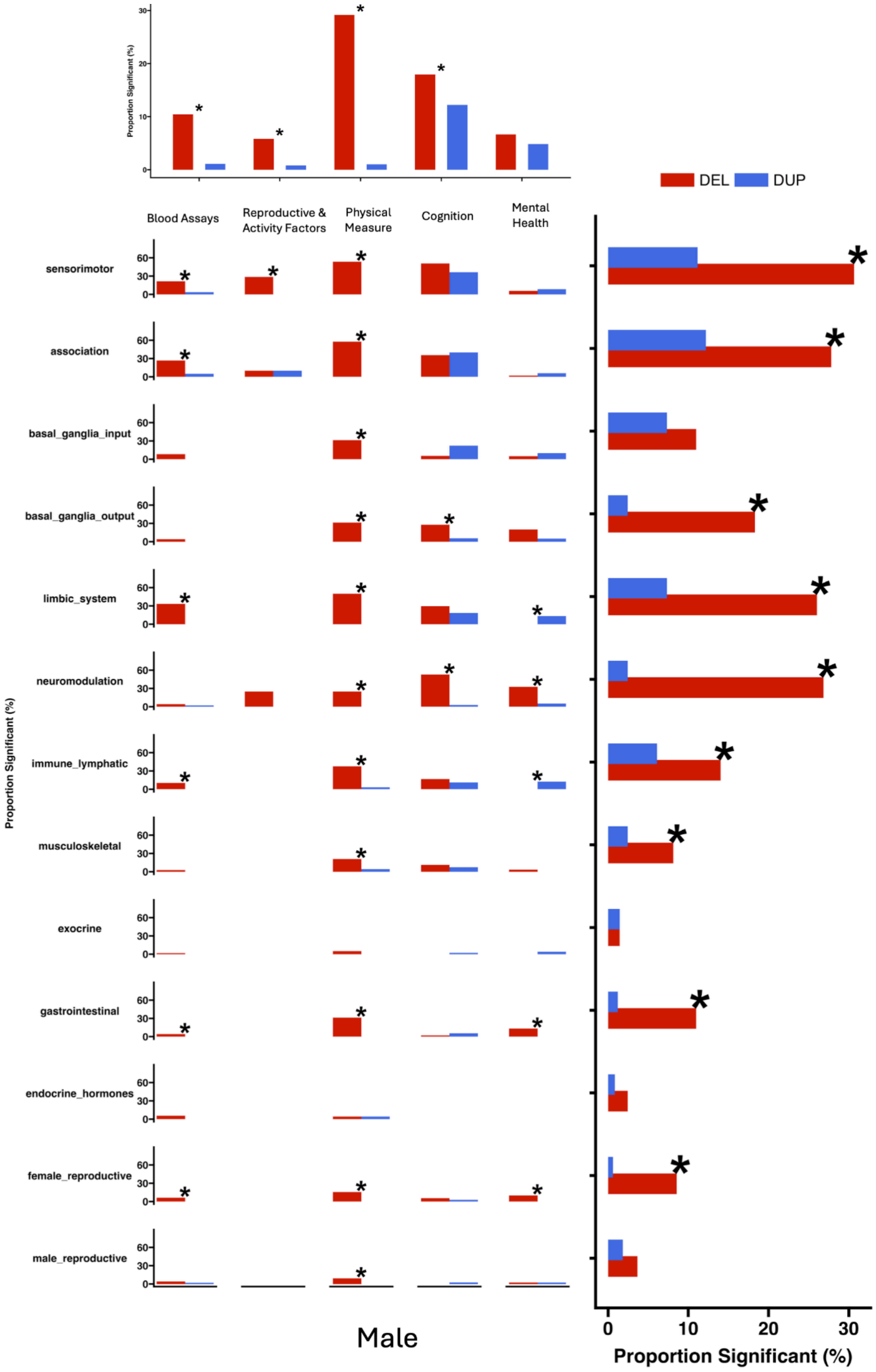


**Supplementary Figure 25: Differences between del-dup associations across trait and gene-set categories for Male**

Bar plots summarizing the differences in the level of association between deletions and duplications (proportion of significant association) for five trait categories and functional gene set categories/annotations for sex stratified analysis (Male). Marginal plots show the differences per trait category (top) or gene-set category (right). Black asterisks (*) indicate statistically significant proportion differences (FDR-adjusted).

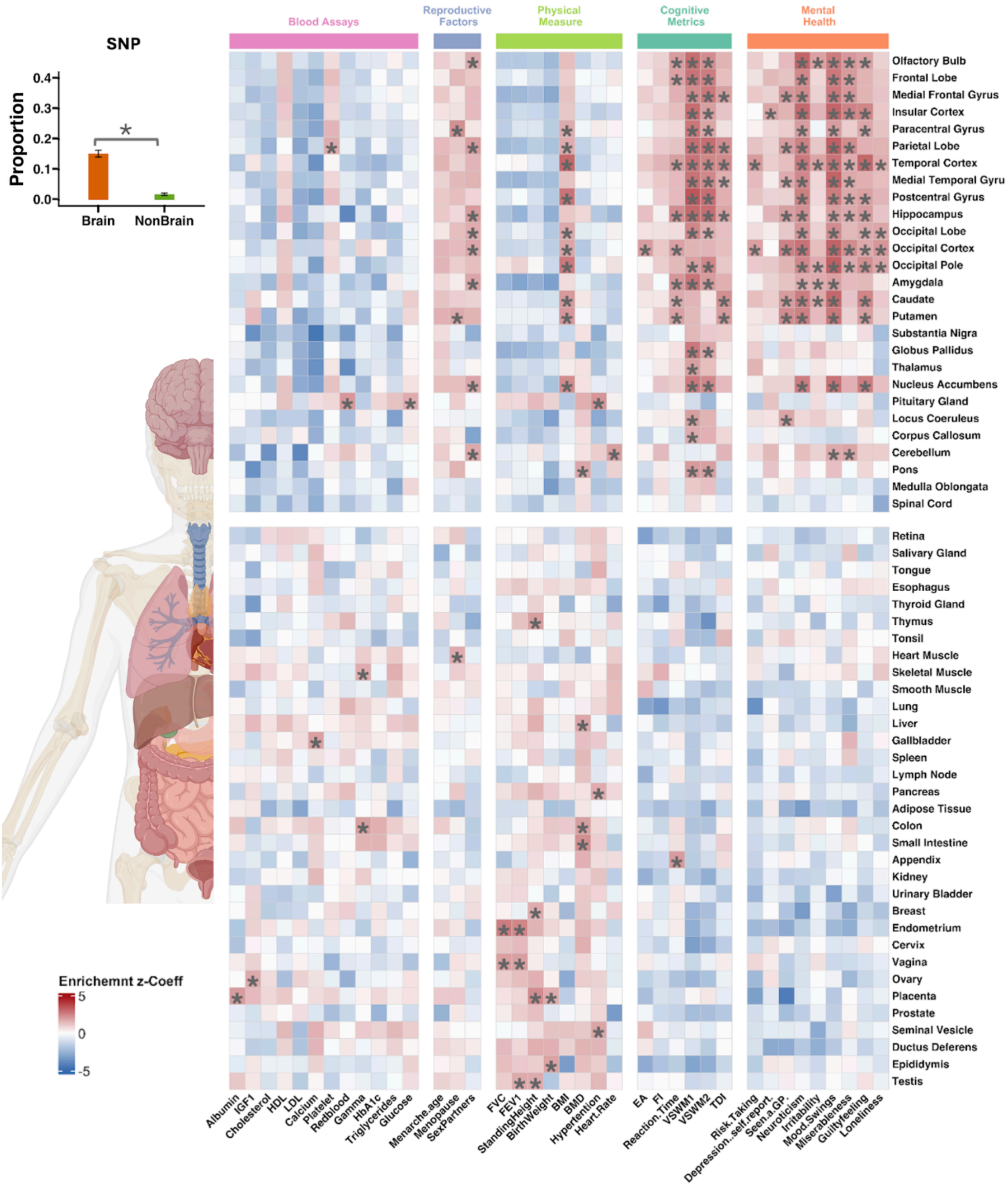


**Supplementary Figure 26: Heatmap of GWAS enrichments for whole body tissues across traits using S-LDSC.**

The heatmap displays S-LDSC-derived GWAS enrichments for five categories of traits (x-axis) across tissue-specific gene sets (y-axis). The color intensity reflects the enrichment Z coefficient. Black asterisks (*) indicate statistically significant associations between traits and genes (nominal).

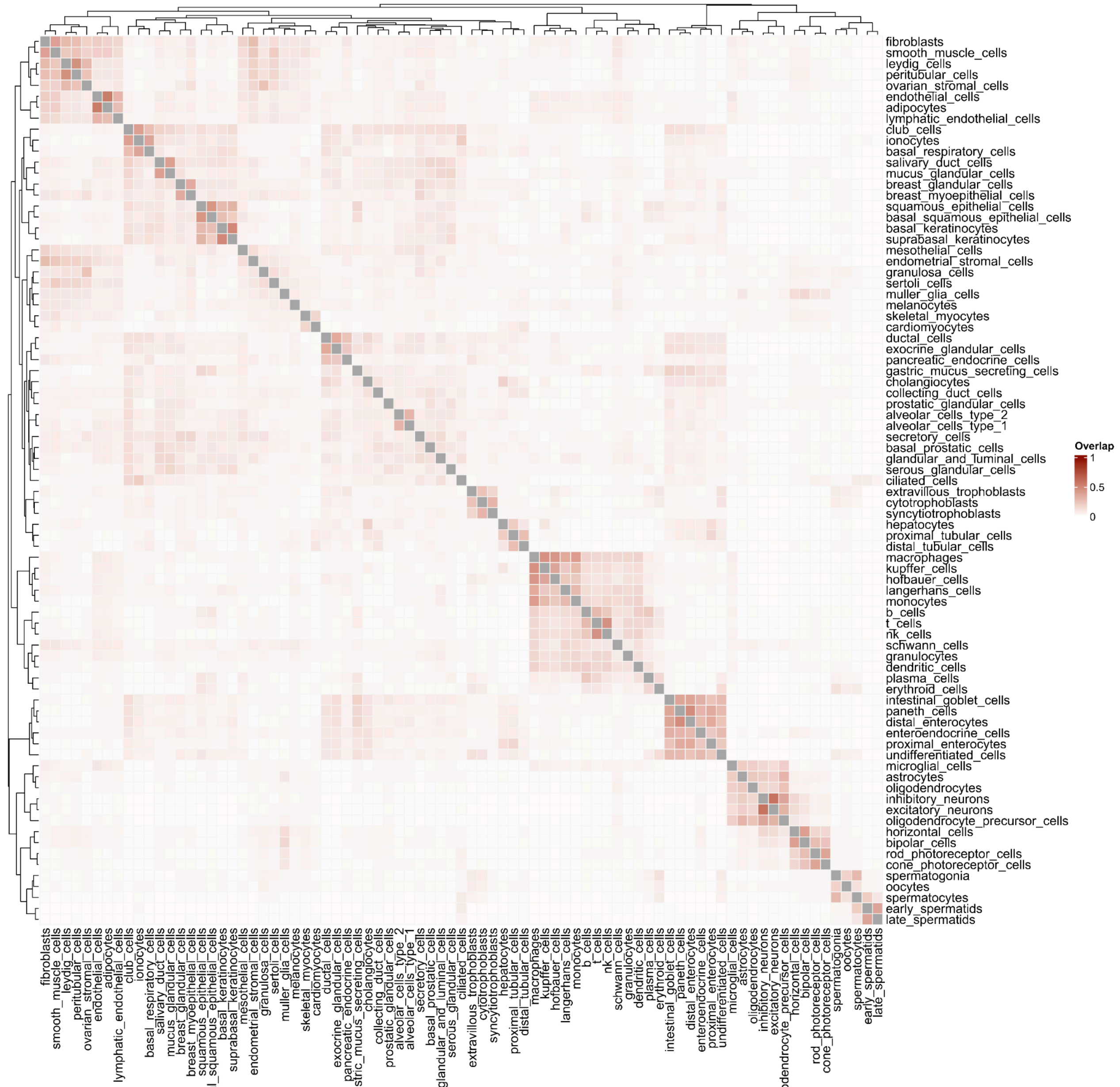


**Supplementary Figure 27: Pairwise overlap matrix of whole body gene sets**

The heatmap displays the pairwise overlap between whole-body cell-type gene sets. The maximum, minimum, mean, and median values of the overlap are: 58.8%, 22.9%, 4.6%, and 3.0%.

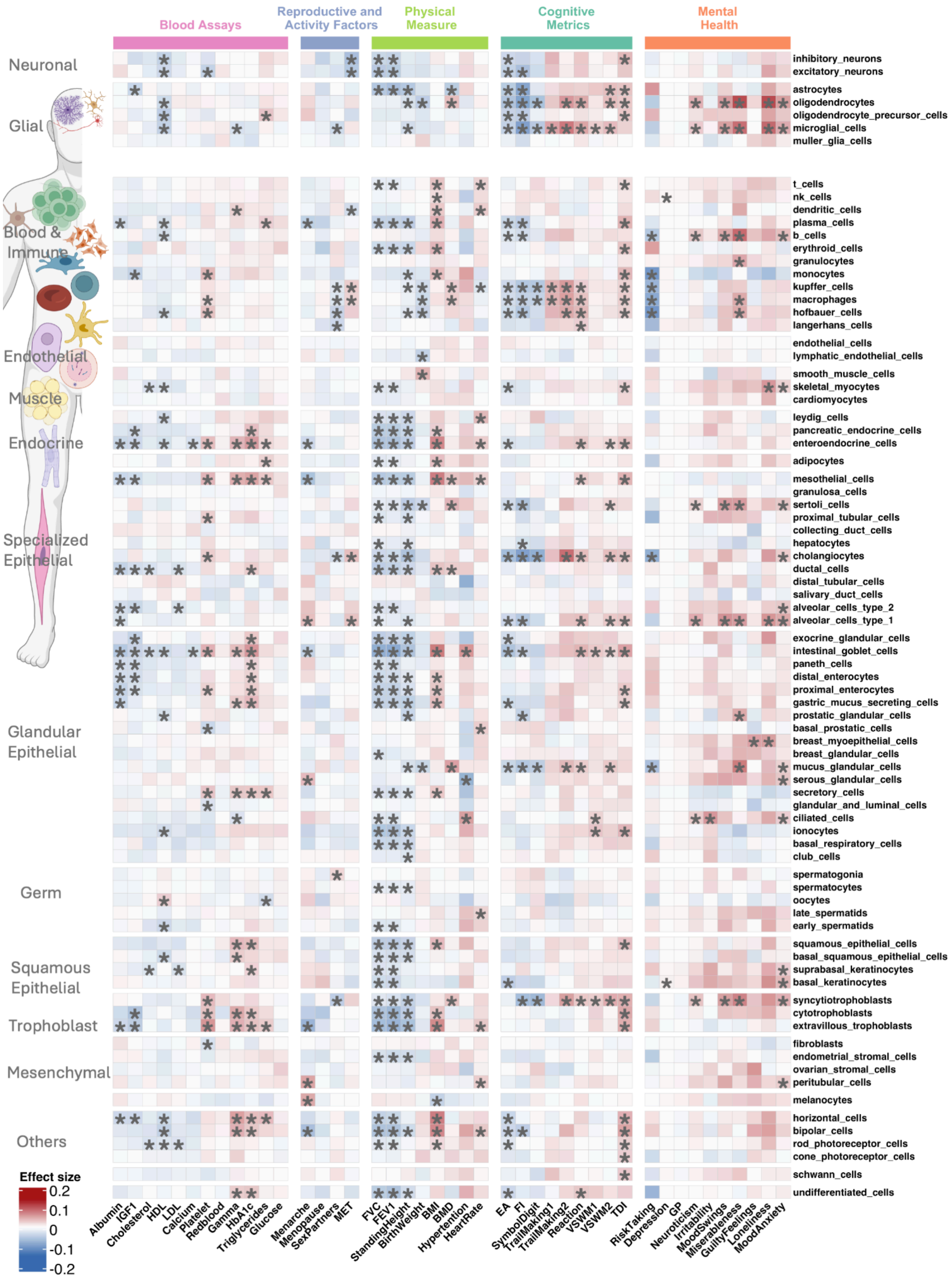


**Supplementary Figure 28: Heatmap of deletion effect sizes for whole-body cells across traits**
The heatmap displays deletion burden association effect sizes between five categories of traits (x-axis) and cell-type-specific gene sets (y-axis). y-axis: gene sets with their categories/annotations indicated on the right side. *: FDR significant associations.

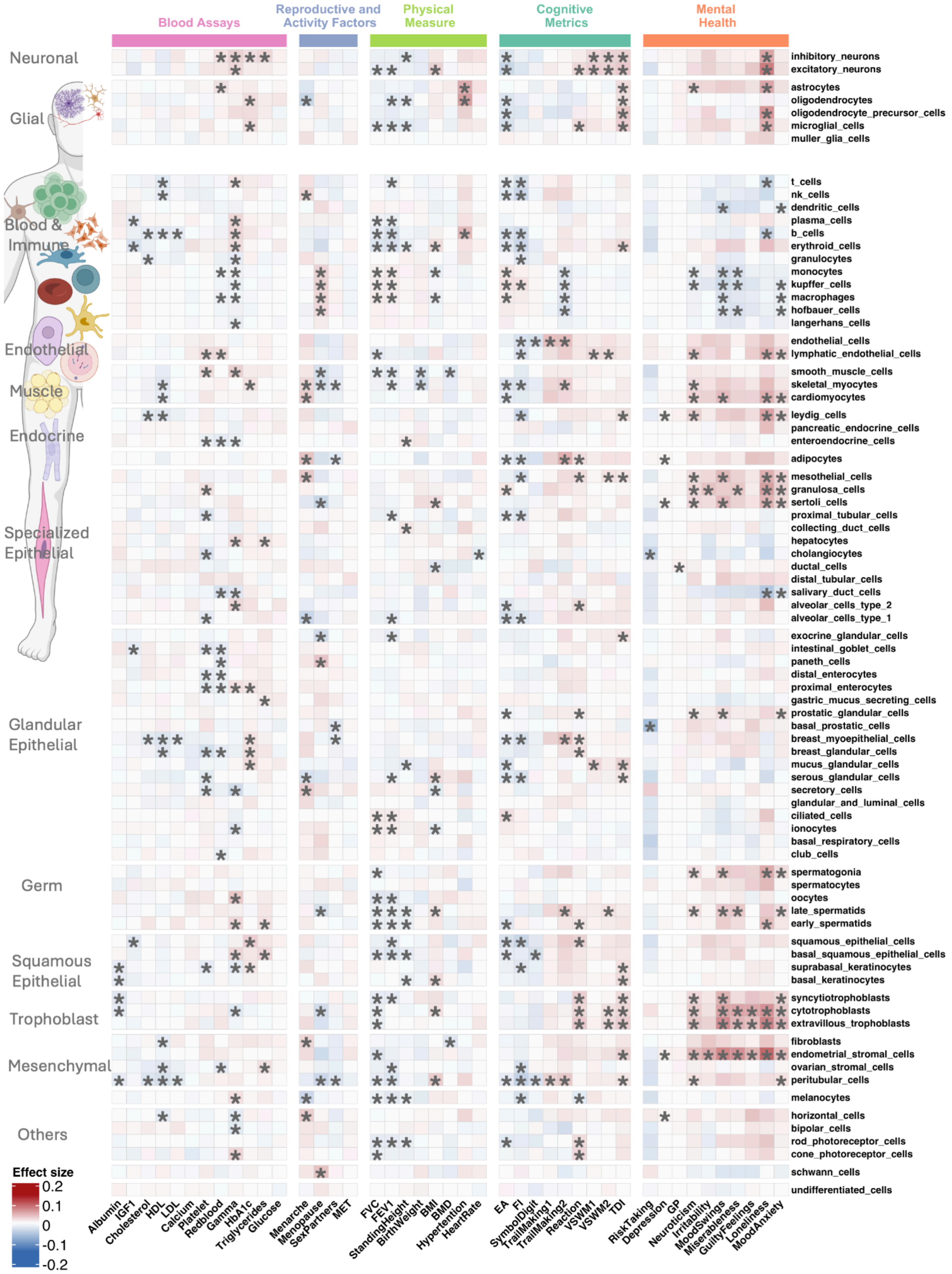


**Supplementary Figure 29: Heatmap of duplication effect sizes for whole-body cells across traits**

The heatmap displays duplication burden association effect sizes between five categories of traits (x-axis) and cell-type-specific gene sets (y-axis, with their categories/annotations indicated on the right side). *: FDR significant associations.

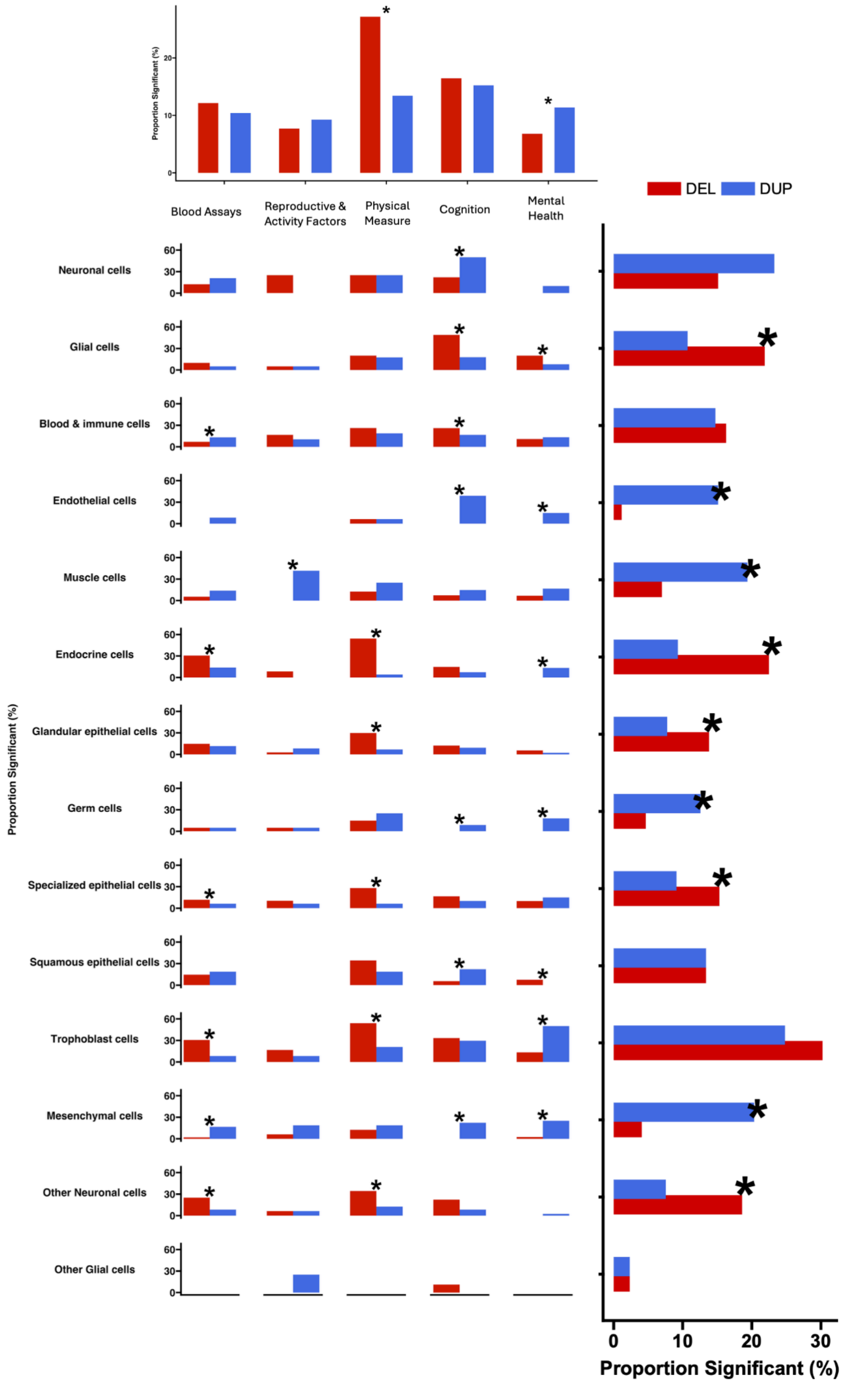


**Supplementary Figure 30: Differences between del-dup associations across trait and cell-type gene-set categories**

Bar plots summarizing the differences in the level of association between deletions and duplications (proportion of significant association) for five trait categories and functional gene set categories/annotations. Marginal plots show the differences per trait category (top) or gene-set category (right). *: statistically significant proportion differences (FDR-adjusted).

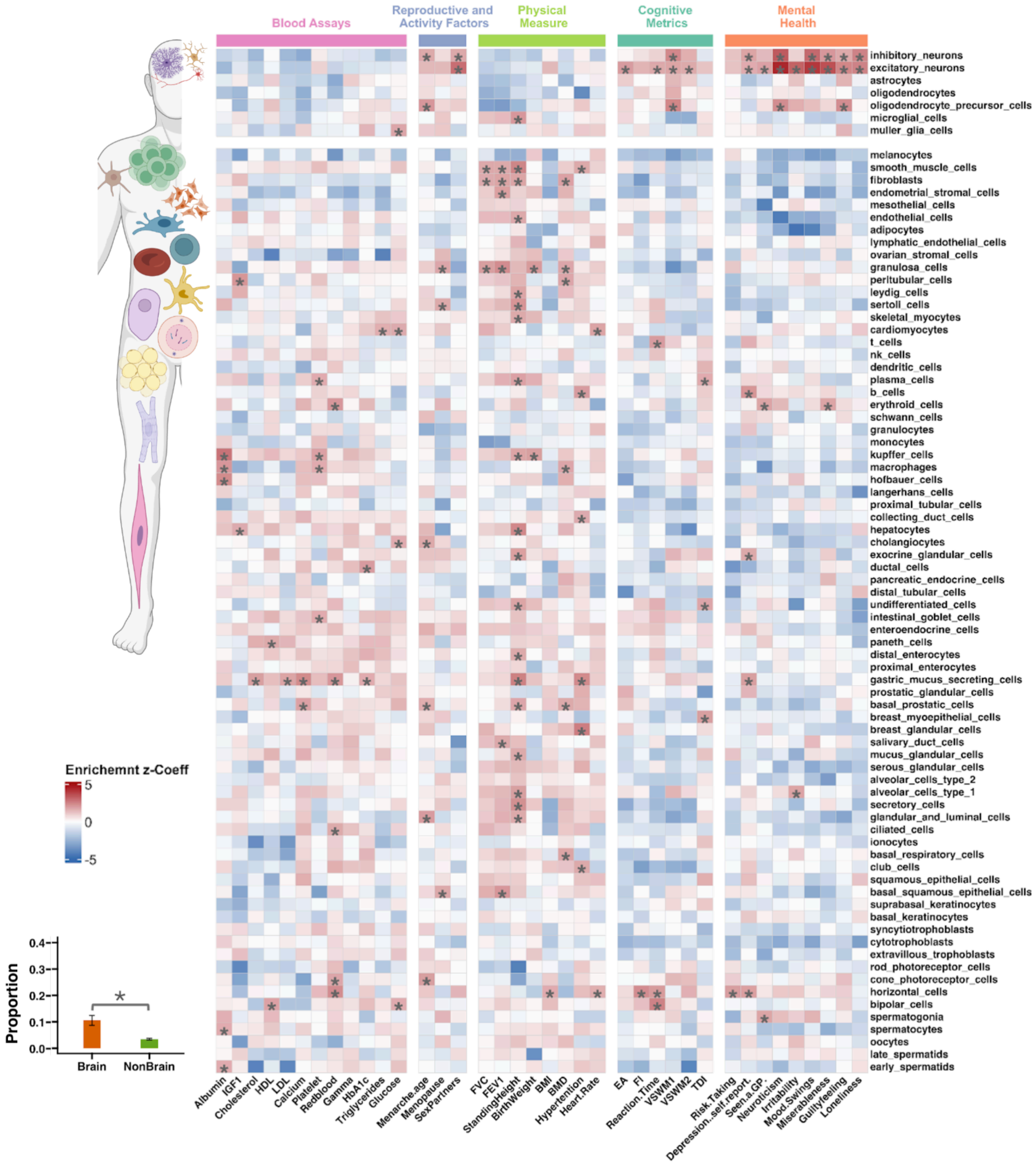


**Supplementary Figure 31: Heatmap of GWAS enrichments for whole body cell types across traits using S-LDSC.**

The heatmap displays S-LDSC-derived GWAS enrichments for five categories of traits (x-axis) across tissue-specific gene sets (y-axis). The color intensity reflects the enrichment Z coefficient. Black asterisks (*) indicate statistically significant associations between traits and genes (nominal).

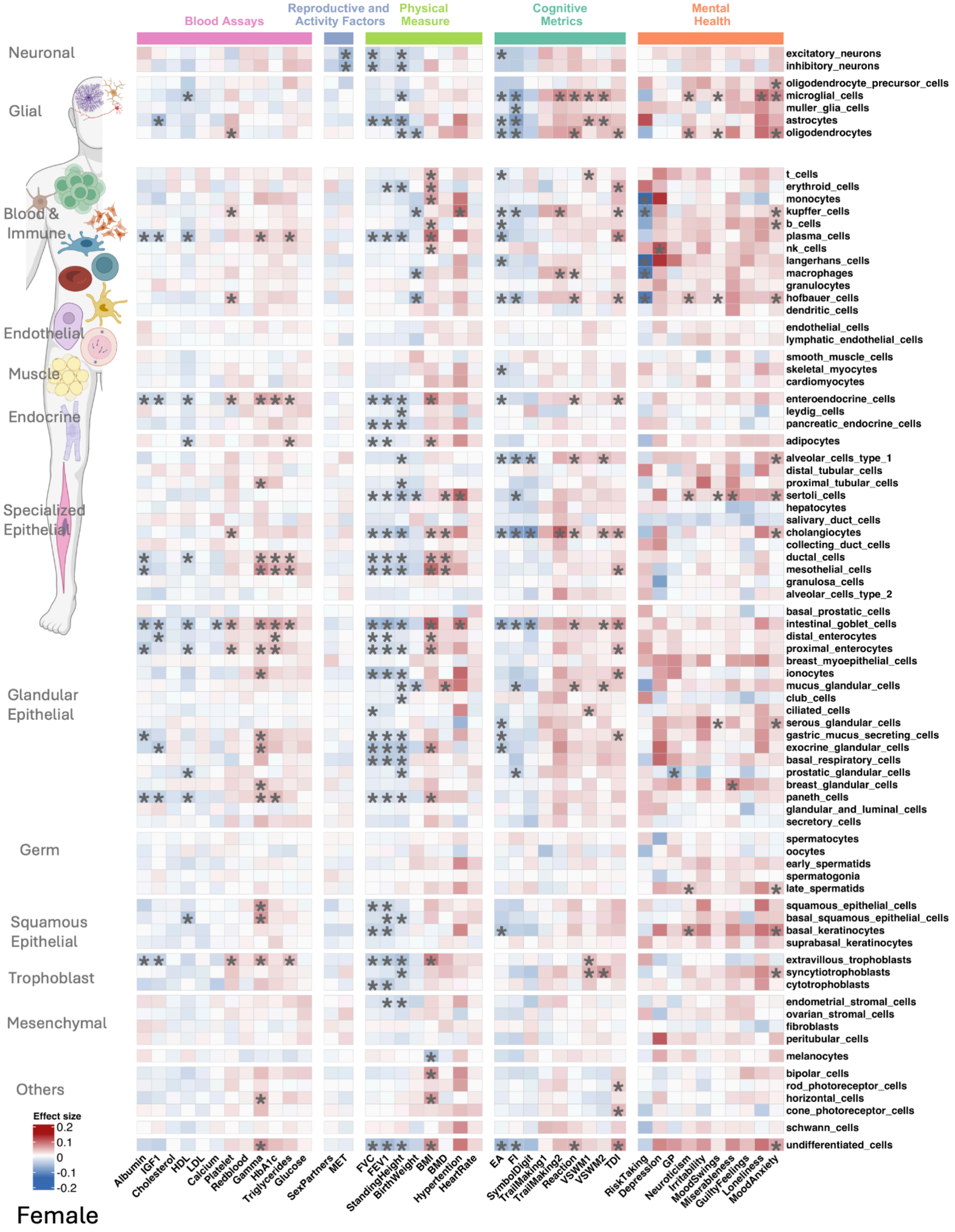


**Supplementary Figure 32: Heatmap of deletion effect sizes for whole-body cells across traits for Female**

Sex stratified (Female) deletion burden association effect sizes between five categories of traits (x-axis) and cell-type-specific gene sets (y-axis, with their categories/annotations indicated on the right side). blue = negative effect size; red = positive effect size. * indicate statistically significant associations between traits and genes.

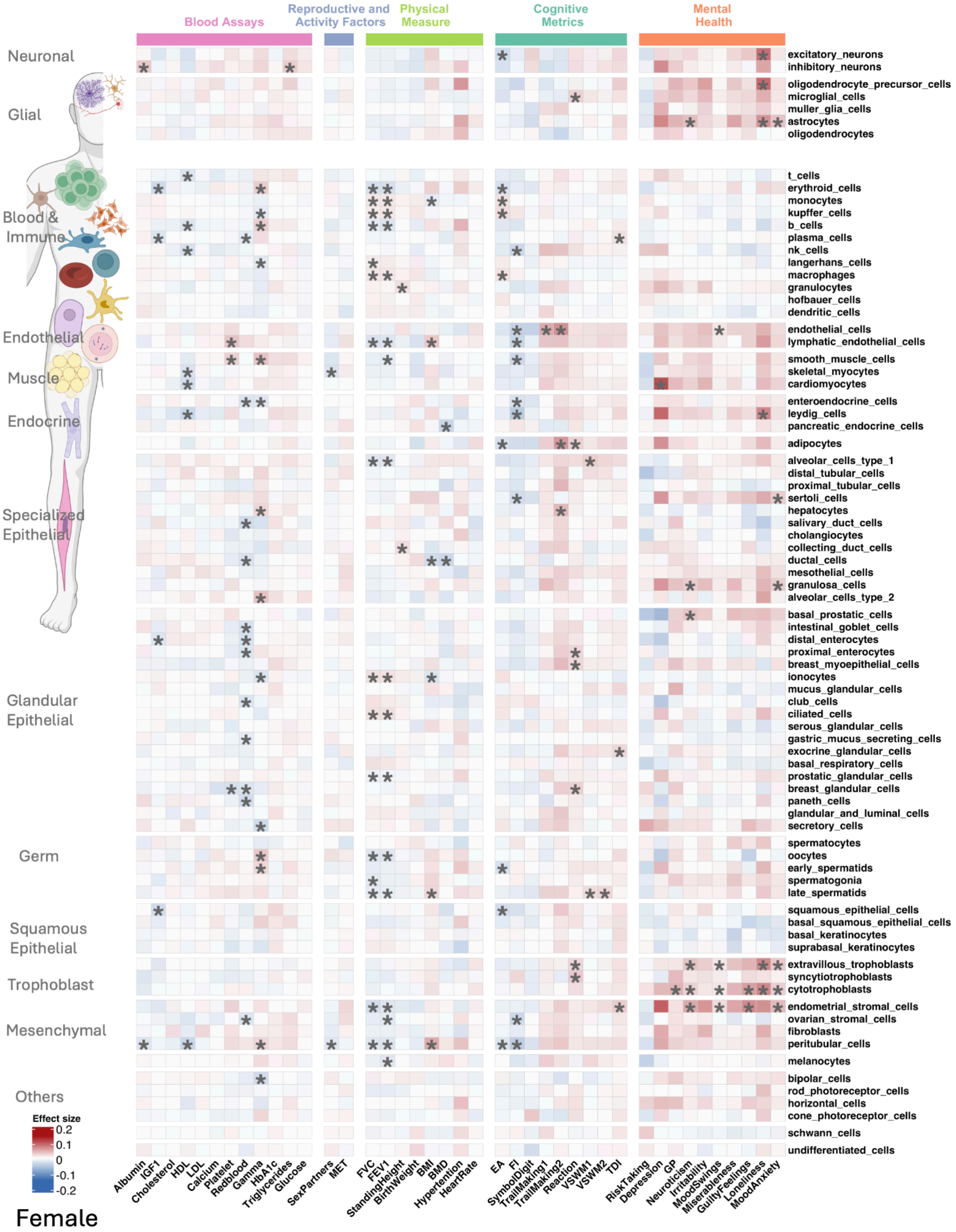


**Supplementary Figure 33: Heatmap of duplication effect sizes for whole-body cells across traits for Female**

Sex stratified (Female) duplication burden association effect sizes between five categories of traits (x-axis) and cell-type-specific gene sets (y-axis, with their categories/annotations indicated on the right side). * indicate statistically significant associations between traits and genes.

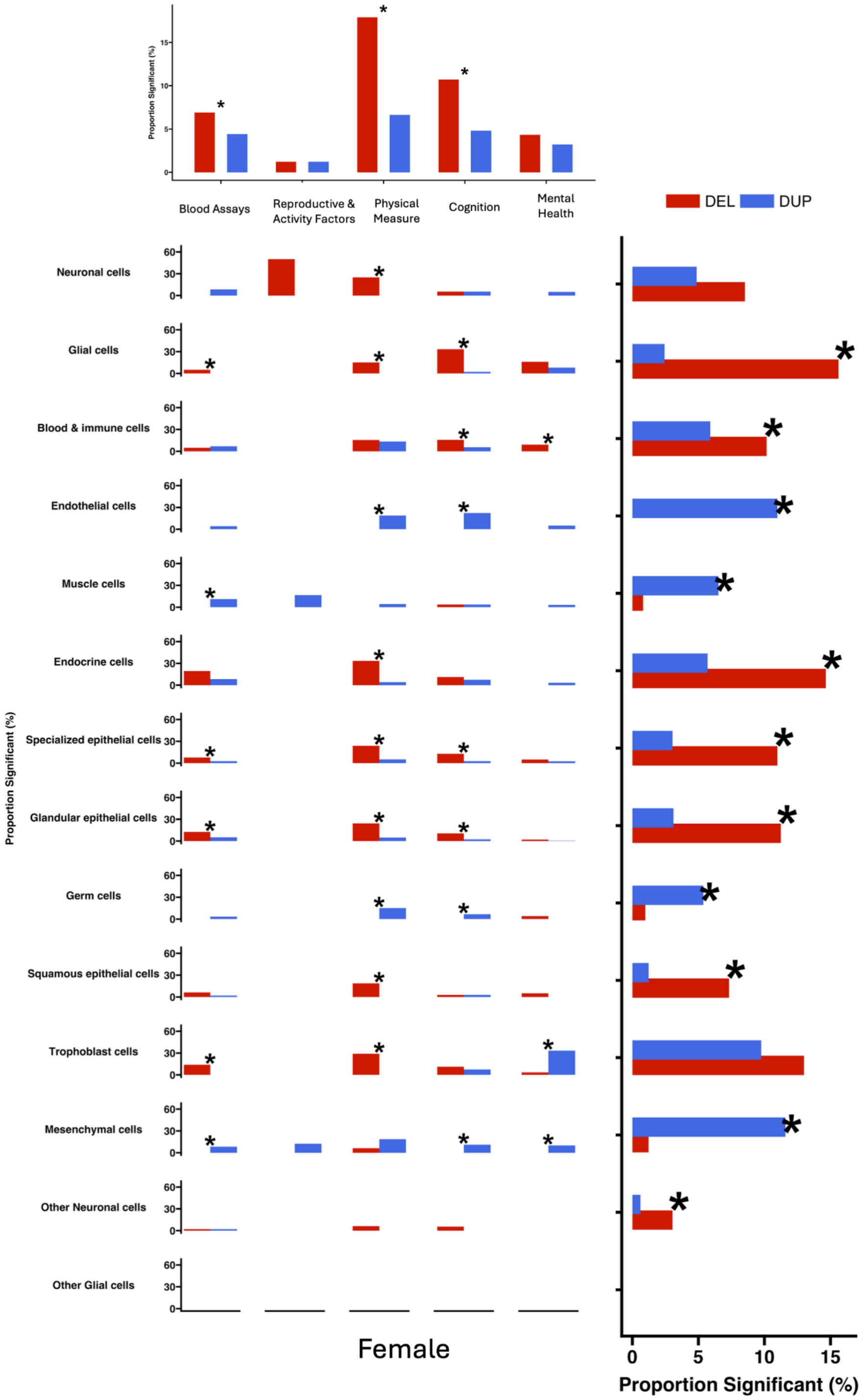


**Supplementary Figure 34: Differences between del-dup associations across trait and cell-type gene-set categories for Female**

Bar plots summarizing the differences in the level of association between deletions and duplications (proportion of significant association) for five trait categories and functional gene set categories/annotations. Marginal plots show the differences per trait category (top) or gene-set category (right). *: statistically significant proportion differences (FDR-adjusted).

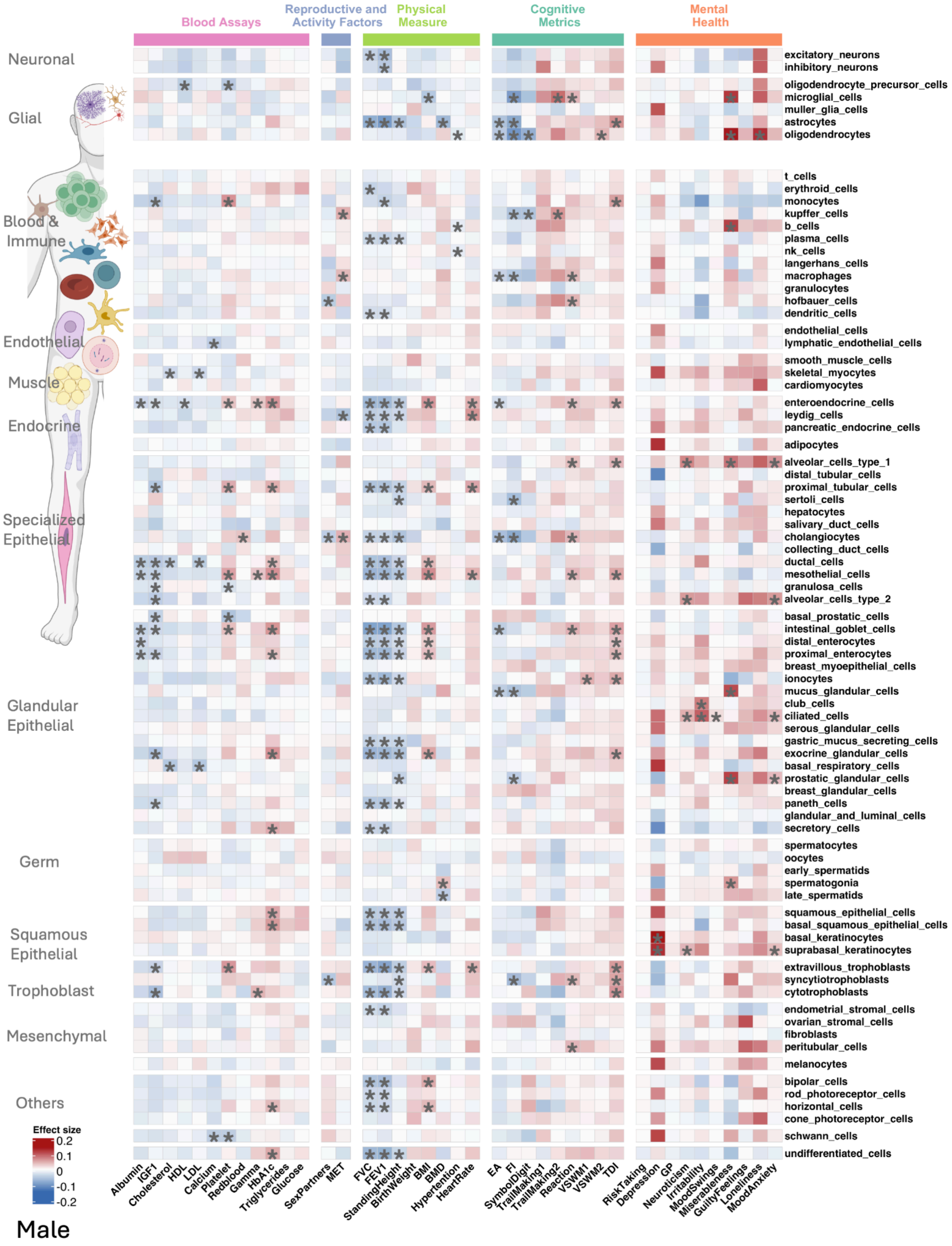

**Supplementary Figure 35: Heatmap of deletion effect sizes for whole-body cells across traits for Male**

Sex stratified (Male) deletion burden association effect sizes between five categories of traits (x-axis) and cell-type-specific gene sets (y-axis, with their categories/annotations indicated on the right side). * indicate statistically significant associations between traits and genes.

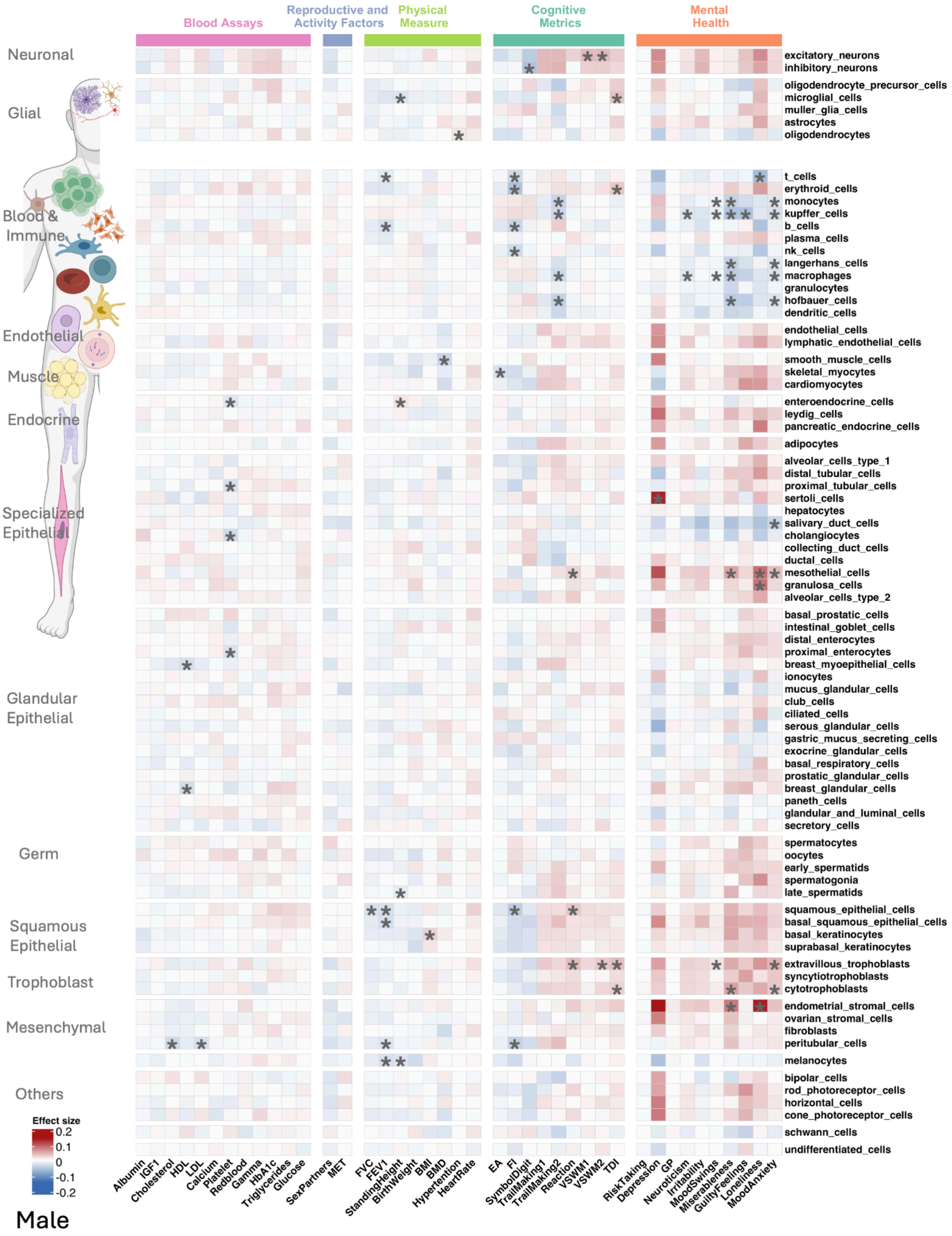

**Supplementary Figure 36: Heatmap of duplication effect sizes for whole-body cells across traits for Male**

Sex stratified (Male) duplication burden association effect sizes between five categories of traits (x-axis) and cell-type-specific gene sets (y-axis, with their categories/annotations indicated on the right side). * indicate statistically significant associations between traits and genes.

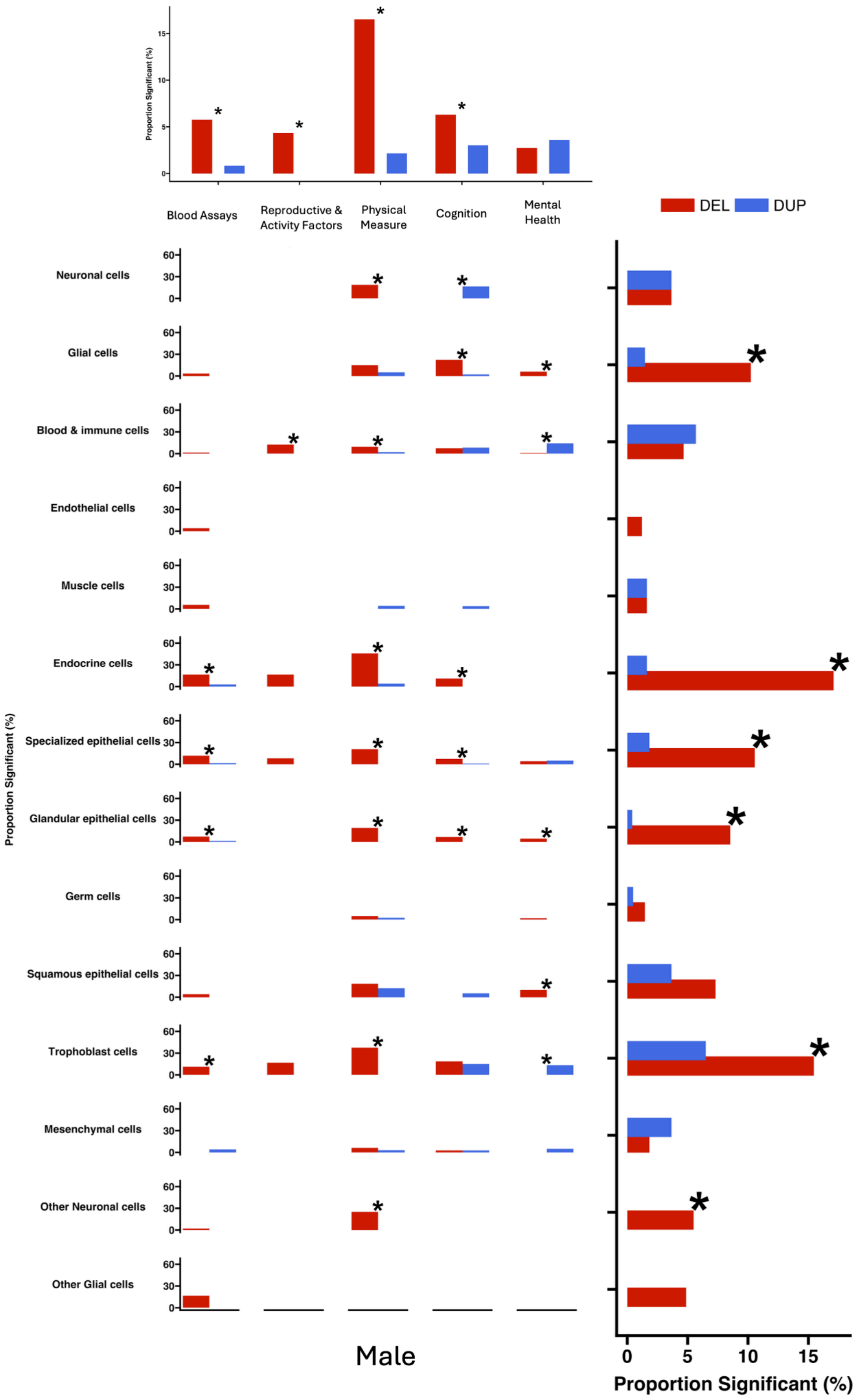

**Supplementary Figure 37: Differences between del-dup associations across trait and gene-set categories for Male**

Bar plots summarizing the differences in the level of association between deletions and duplications (proportion of significant association) for five trait categories and functional gene set categories/annotations. Marginal plots show the differences per trait category (top) or gene-set category (right). *: statistically significant proportion differences (FDR-adjusted).

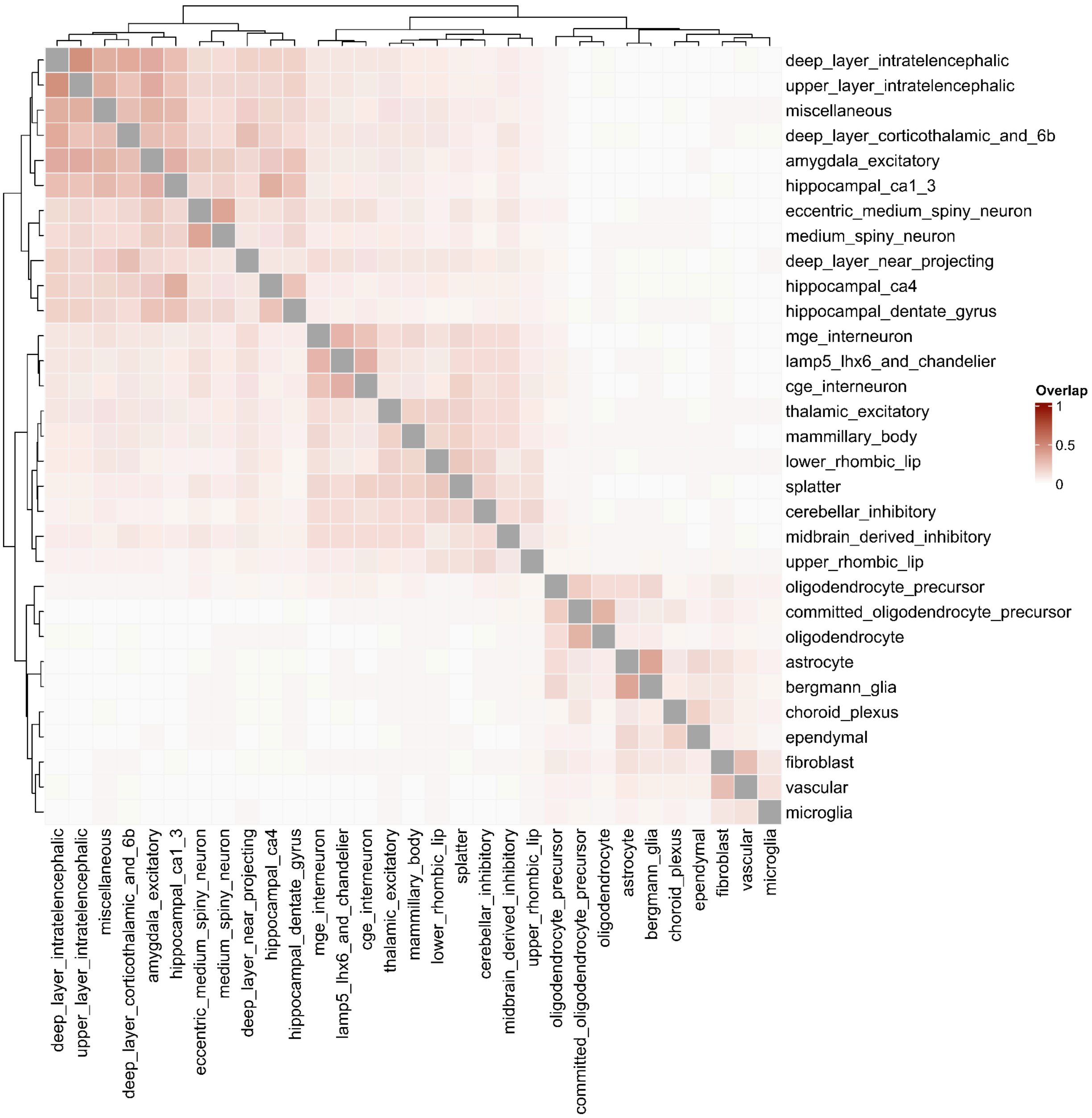

**Supplementary Figure 38: Pairwise overlap matrix of whole brain cell gene sets**

The heatmap displays the pairwise overlap between whole-brain gene sets. The maximum, minimum, mean, and median values of the overlap are: 46.7%, 0.98%, 7.5% and 4.9%.

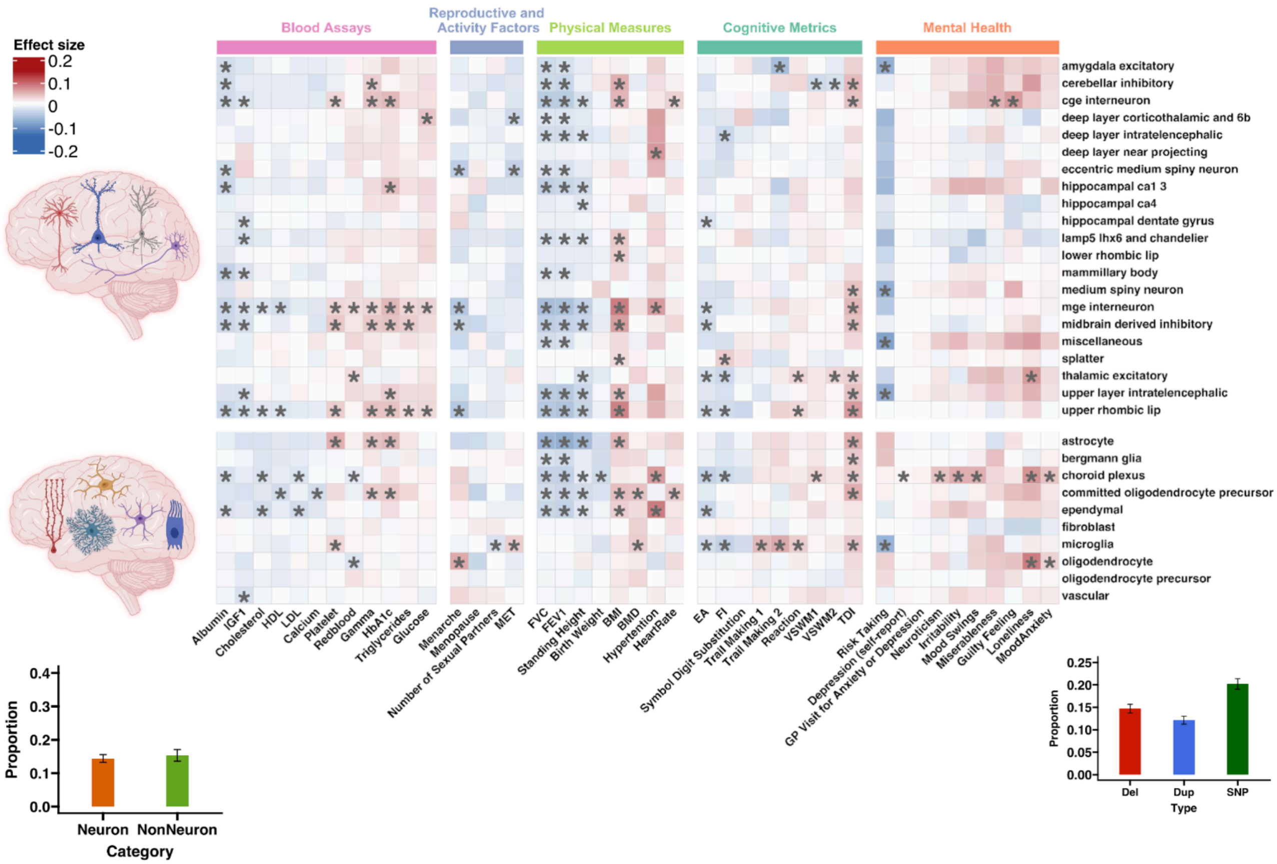

**Supplementary Figure 39: Heatmap of deletion effect sizes for whole-brain cells across traits**

The heatmap displays deletion burden association effect sizes between five categories of traits (x-axis) and whole-brain cell-type-specific gene sets (y-axis). blue = negative effect size; red = positive effect size. * indicate statistically significant associations between traits and genes (FDR correction across all gene sets, traits, and CNV type, 172 ✕ 43 ✕ 2 =14,792 tests).

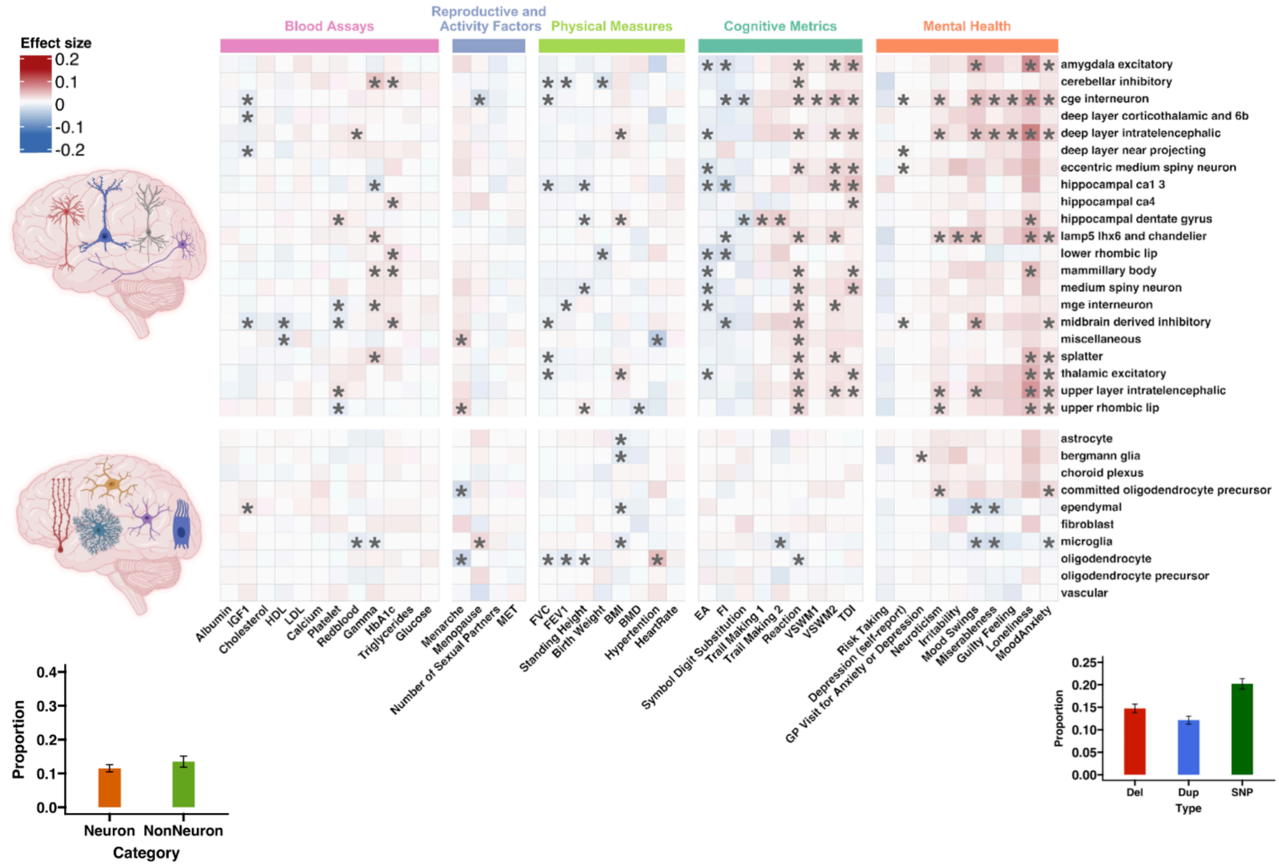


**Supplementary Figure 40: Heatmap of duplication effect sizes for whole-brain cells across traits**

The heatmap displays duplication burden association effect sizes between five categories of traits (x-axis) and whole-brain cell-type-specific gene sets (y-axis). blue = negative effect size; red = positive effect size. * indicate statistically significant associations between traits and genes (FDR correction across all gene sets, traits, and CNV type, 172 ✕ 43 ✕ 2 =14,792 tests).

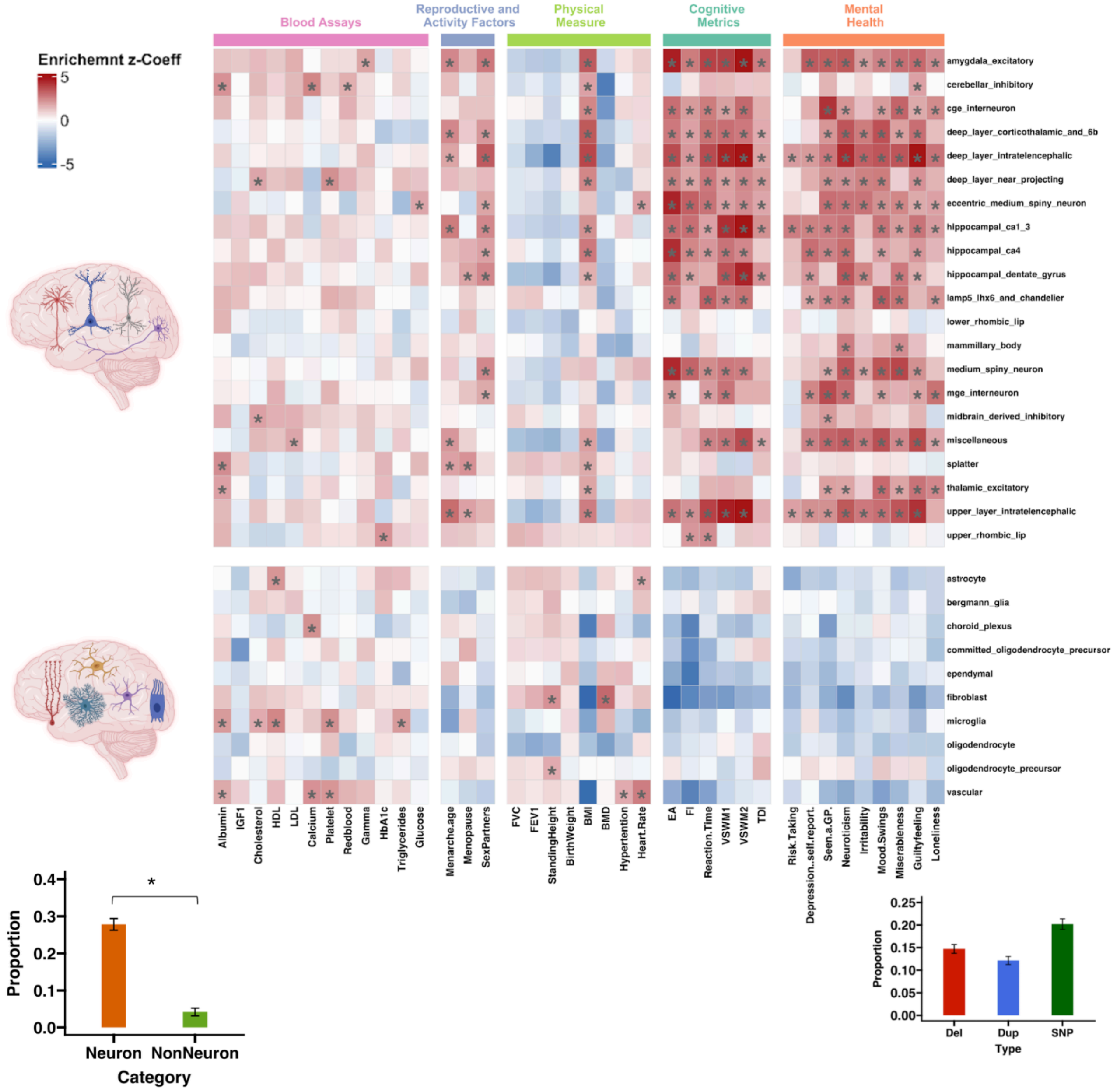


**Supplementary Figure 41: Heatmap of GWAS enrichments for whole brain cell types across traits using S-LDSC.**

The heatmap displays S-LDSC-derived GWAS enrichments for five categories of traits (x-axis) across gene sets (y-axis). The color intensity reflects the enrichment Z coefficient. Black asterisks (*) indicate statistically significant associations between traits and genes (nominal).

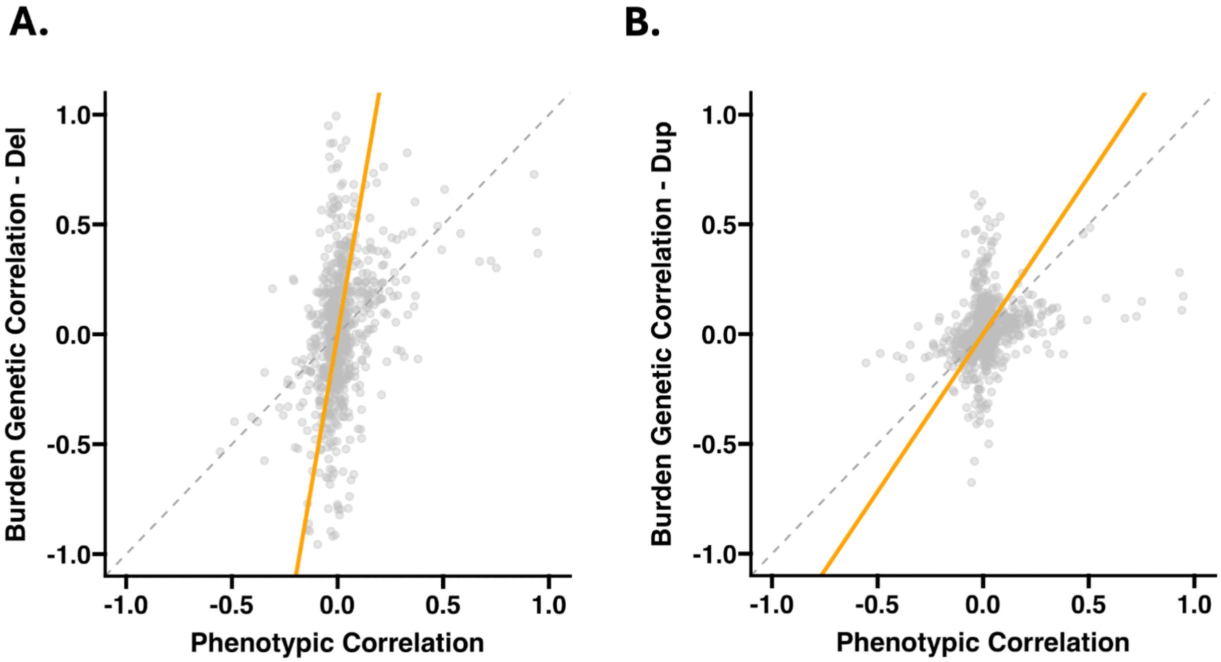


**Supplementary Figure 42: Comparing between-trait CNV burden correlations with phenotypic correlations**

Concordance scatterplots comparing between-trait A) deletion and B) duplication burden correlations with between-trait phenotypic correlations.

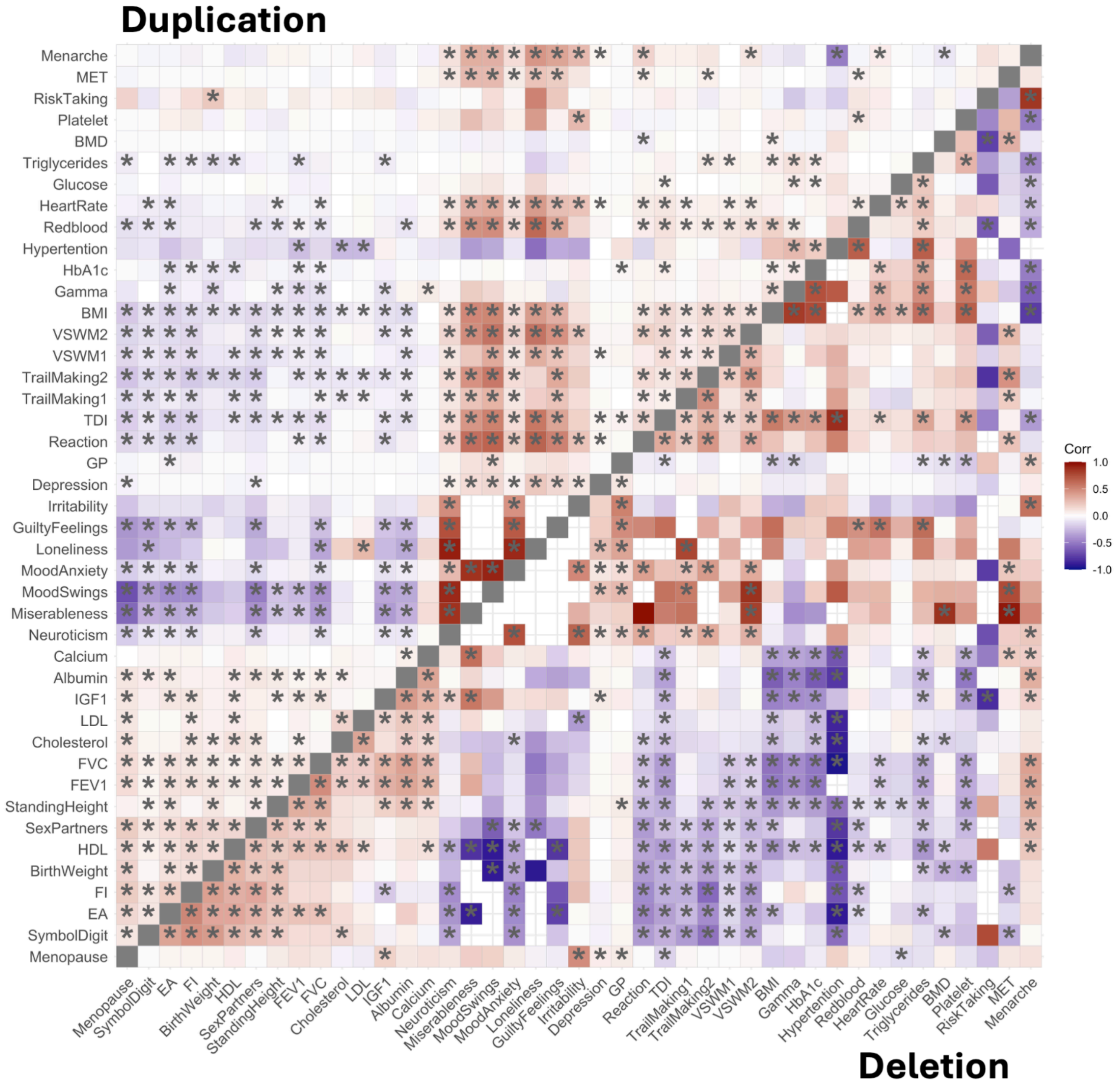


**Supplementary Figure 43: Pairwise CNV burden genetic correlations across 43 traits**

Heatmap of between-trait CNV burden correlations. Deletion correlations are in the lower triangle matrix, and duplications are in the upper triangle matrix. Negative correlations are shown in purple, and positive correlations are in red. Correlations with an FDR-corrected p-Jaccard significance are marked with an asterisk (*). White spaces indicate CNV burden correlations that exceeded 1 after adjusting for CNV heritability (R2, variance explained, **Methods, Table ST17**).

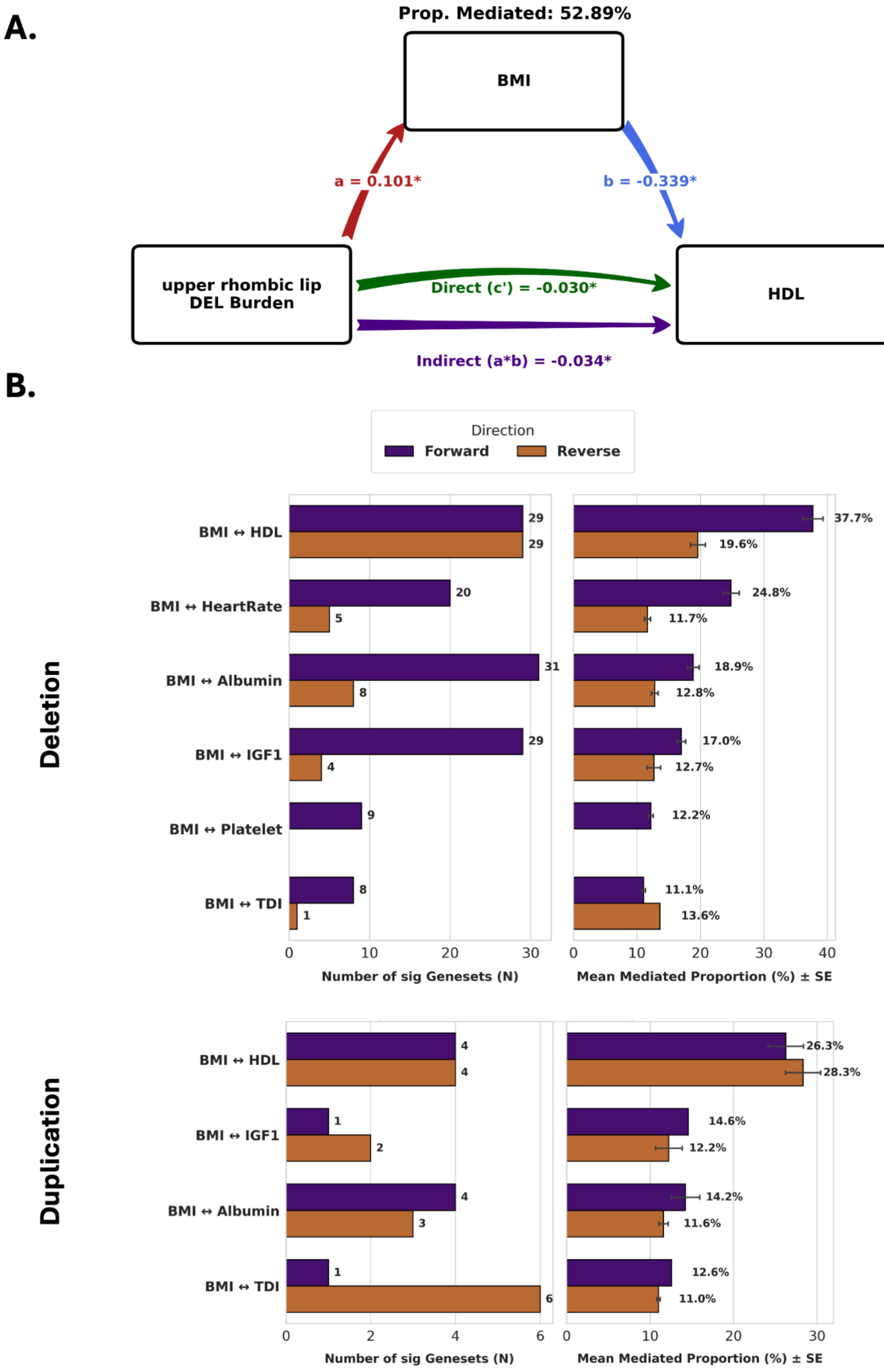


**Supplementary Figure 44: Mediation analysis to distinguish biological and mediated pleiotropy**

(A) Representative Deletion Mediation Model. Path diagram illustrating the decomposition of the deletion burden effect in the "upper rhombic lip" gene set on HDL cholesterol, using BMI as the mediator. The model partitions the total effect into a direct genetic effect (c' = -0.030, representing biological pleiotropy) and an indirect effect (a * b = -0.034, representing mediated pleiotropy). In this specific model, 52.89% of the association is mediated through BMI. Path a (0.101) represents the burden effect on the mediator; path b (-0.339) represents the effect of the mediator on the outcome. Asterisks * denote significant path coefficients ($p < 0.05$). (B) Summary of Bidirectional Mediation Across Correlated Trait Pairs. Results of mediation analysis for six trait pairs with CNV burden correlations > 0.3, analyzed across all 172 gene sets for deletions and duplications. Left panels indicate the number of significant gene sets (N) identifying a mediation path (a * b), where both the exposure-to-mediator (path a) and the direct effect (path c') were also statistically significant. Right panels display the mean mediated proportion (%) and standard error (SE) across significant gene sets. Analysis was performed in two directions: forward (BMI → Trait, purple) and reverse (BMI ← Trait, brown). While the direct genetic effect explained the majority of the shared genetic architecture (mean mediation 16%), deletions for BMI–HDL showed a higher mediated proportion (37.7%), suggesting specific metabolic dependencies for deletion-driven associations.

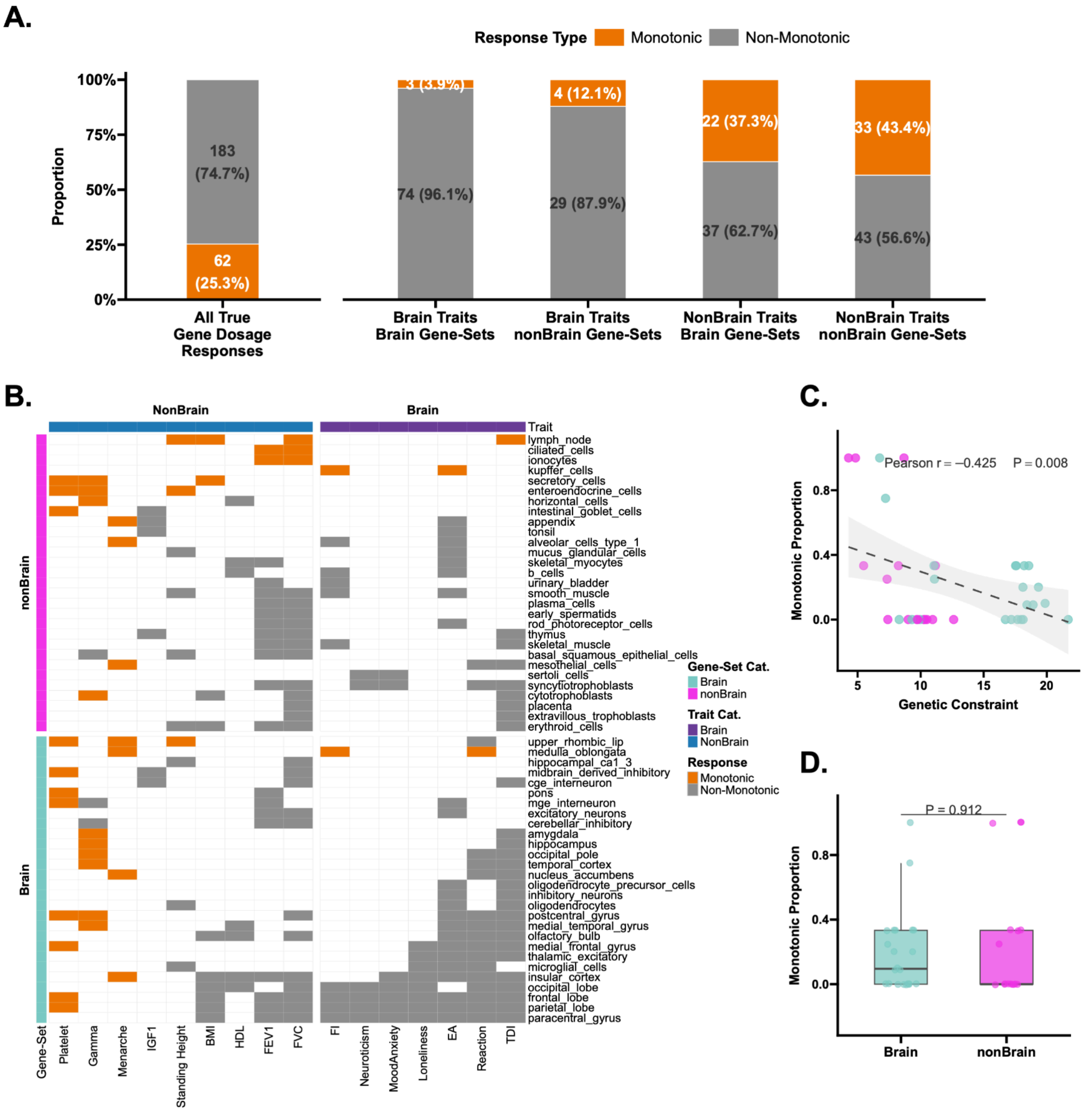


**Supplementary Figure 45: Characteristics of true gene dosage responses across associated traits and gene-sets**

Analysis is restricted to gene-set–trait pairs showing FDR-significant effects for both deletions and duplications. To ensure robust statistics, the data is further restricted to include only traits associated with at least 5 significant gene-sets, and only gene-sets associated with at least 2 traits (N = 245 pairs across 16 traits and 88 gene sets). (A) Stacked bar plots detailing the proportion of monotonic (opposite-direction, orange) versus non-monotonic (same-direction, gray) responses. Overall, 62 pairs (25.3%) show monotonic responses, while 183 pairs (74.7%) are non-monotonic (Binomial test against null proportion of 0.50, p = 4.8E-15). Stratification reveals that brain traits paired with brain gene sets are 96.1% non-monotonic, whereas non-brain traits display higher monotonic proportions (37.3%–43.4%). (B) Heatmap mapping the specific distribution of monotonic and non-monotonic responses across 16 traits and 57 out of 88 genesets (visualizing only those with >2 associations) combinations. (C) Scatter plot demonstrating a significant negative correlation between genetic constraint and the proportion of monotonic responses at the gene-set level (Pearson r = -0.425, p = 0.008). (D) Box plot comparing the proportion of monotonic responses between brain and non-brain gene sets, showing no significant difference between the categories (p = 0.912).

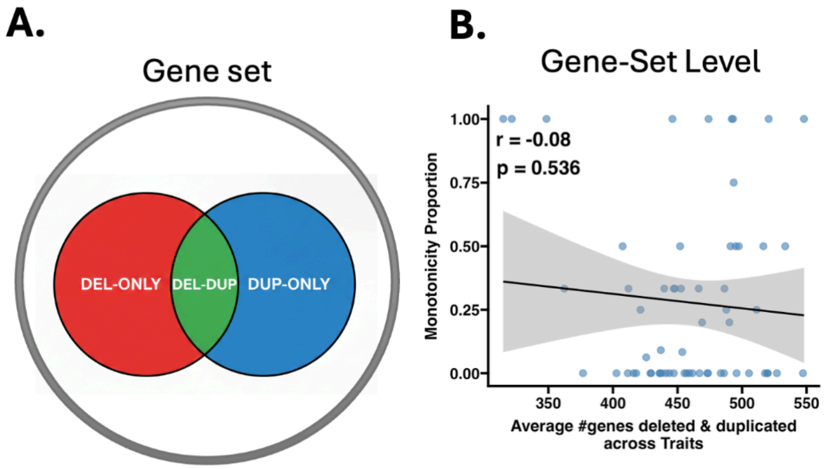


**Supplementary Figure 46: Ability to detect monotonicity is not driven by different genes in a gene set affected by deletions and duplications**

(A) Partitioning of CNV-Impacted Genes Within Gene Sets. Venn diagram illustrating the schematic proportion of genes within a gene set hit exclusively by deletions (DEL-ONLY, red), exclusively by duplications (DUP-ONLY, blue), or by both CNV types (DEL-DUP, green). On average, 35% of genes per gene set are disrupted by both deletions and duplications. (B) Scatter plot showing the number of genes encompassed in both deletions and duplications (x-axis) versus the proportion of monotonic responses (y-axis). Each point is a gene set trait association. No significant correlation was observed (r = −0.08, p = 0.54). Monotonicity is computed exclusively among true gene dosage responses: gene-set–trait pairs where both deletion and duplication associations reach FDR significance (57 out of 172 gene sets and 16 out of 43 traits remained). Black lines represent linear regression fits with shaded 95% confidence intervals.

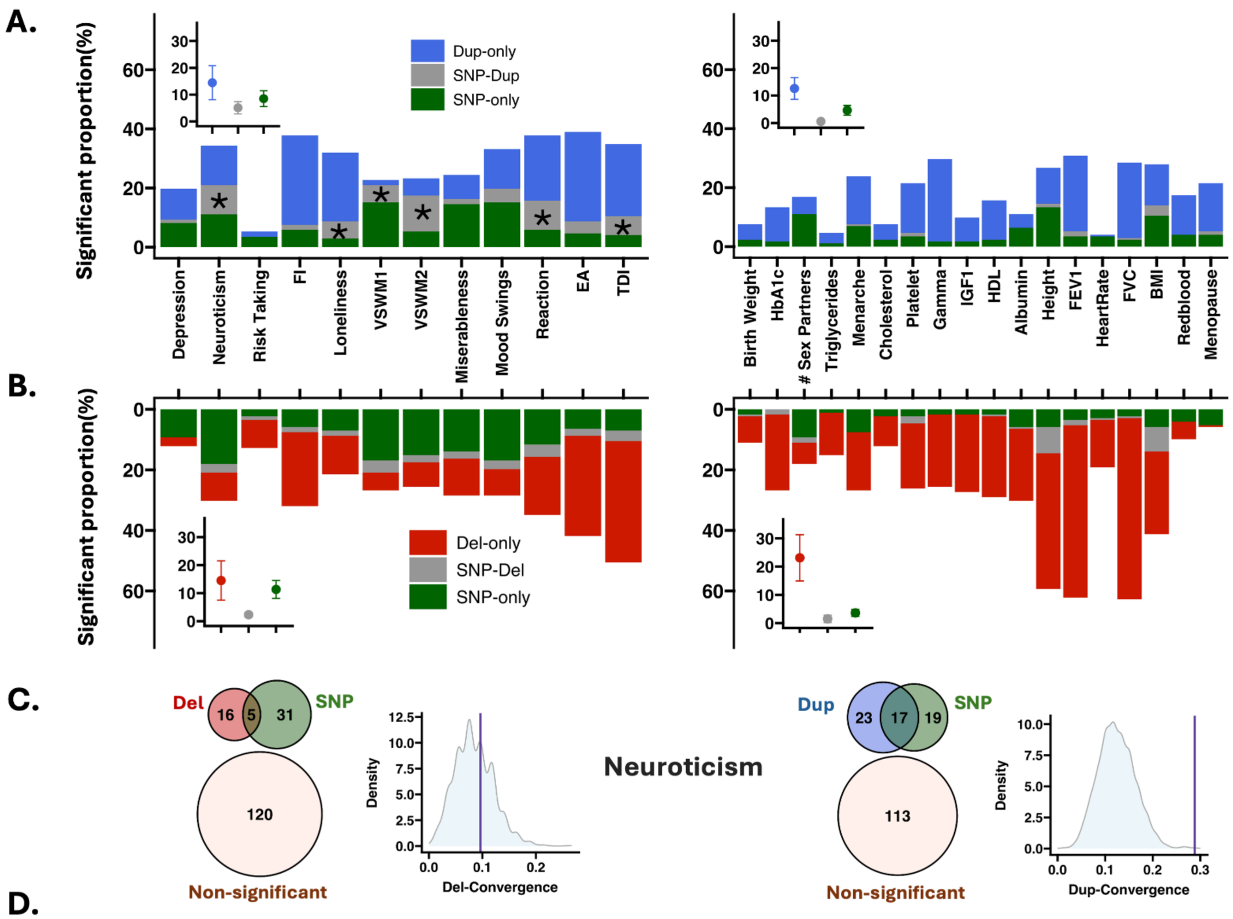


**Supplementary Figure 47: Functional overlap between common variants and CNVs**

**A)** The stack bar plot represents the proportion of functional genes set overlapping between common variants and duplications for each trait. Overlapping functional gene sets (enriched in common variant and CNV signal) are gray, and non-overlapping functional gene sets are represented in blue (associations only significant for duplications or deletions). Stars represent significant functional overlap (FDR permutation p-value < 0.05). Y-axis: proportion of associated gene sets. X-axis: Brain and non-brain traits. The inserts represent the average convergence (between common variants and duplications) and average duplication-specific associations across traits. The same information on functional convergence is shown in **(B)** between deletions and common variants (SNPs). **C)** illustrates the example of the functional overlap between SNPs and CNVs for Neuroticism using a Venn diagram. The density plot represents the null distribution of overlap significance (1000 permutations), with the observed overlap shown by the purple vertical line.

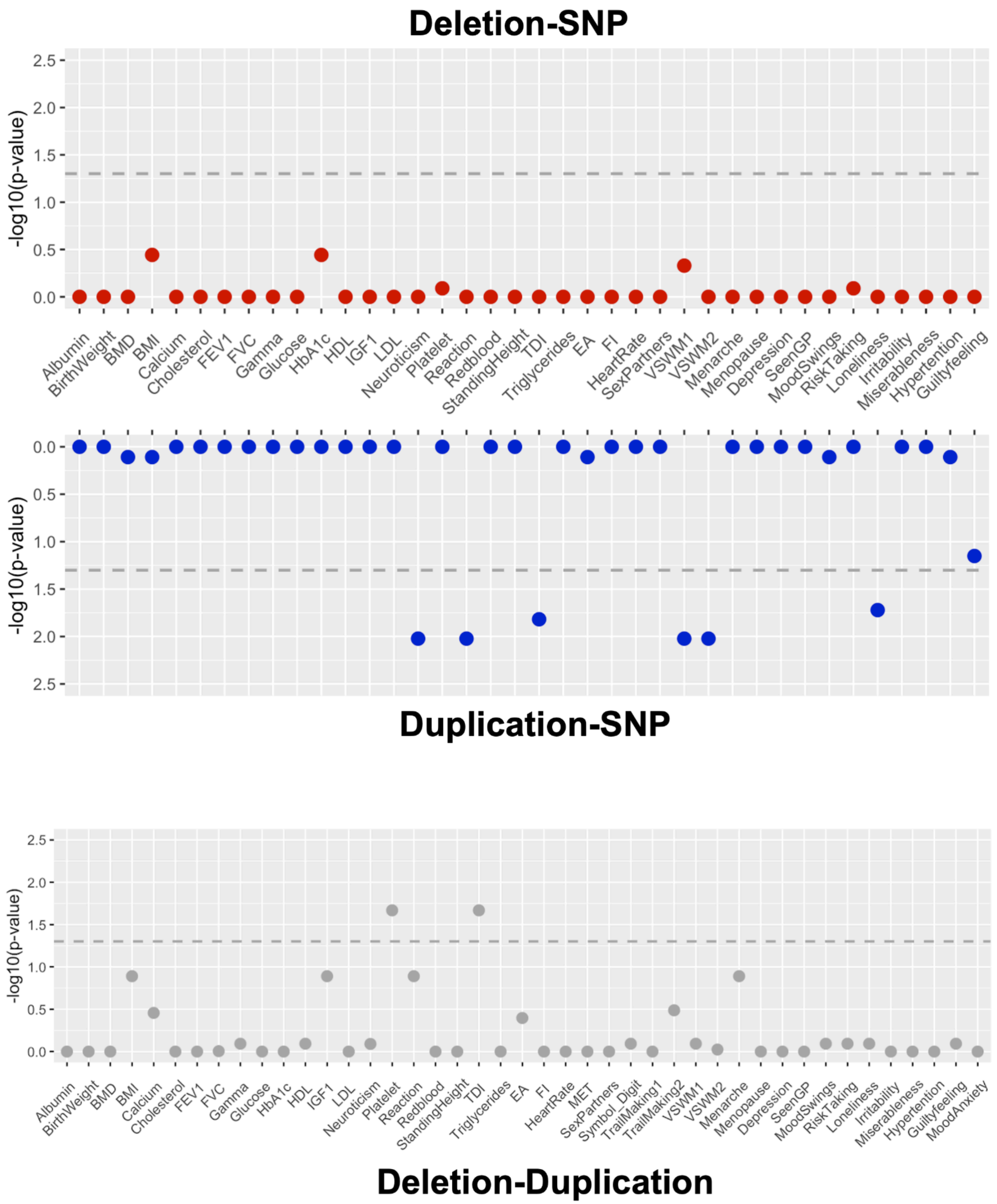


**Supplementary Figure 48: Permutation test P-Values (1000 nulls) for functional overlap between CNV associations and GWAS enrichments, and between deletion and duplication.**

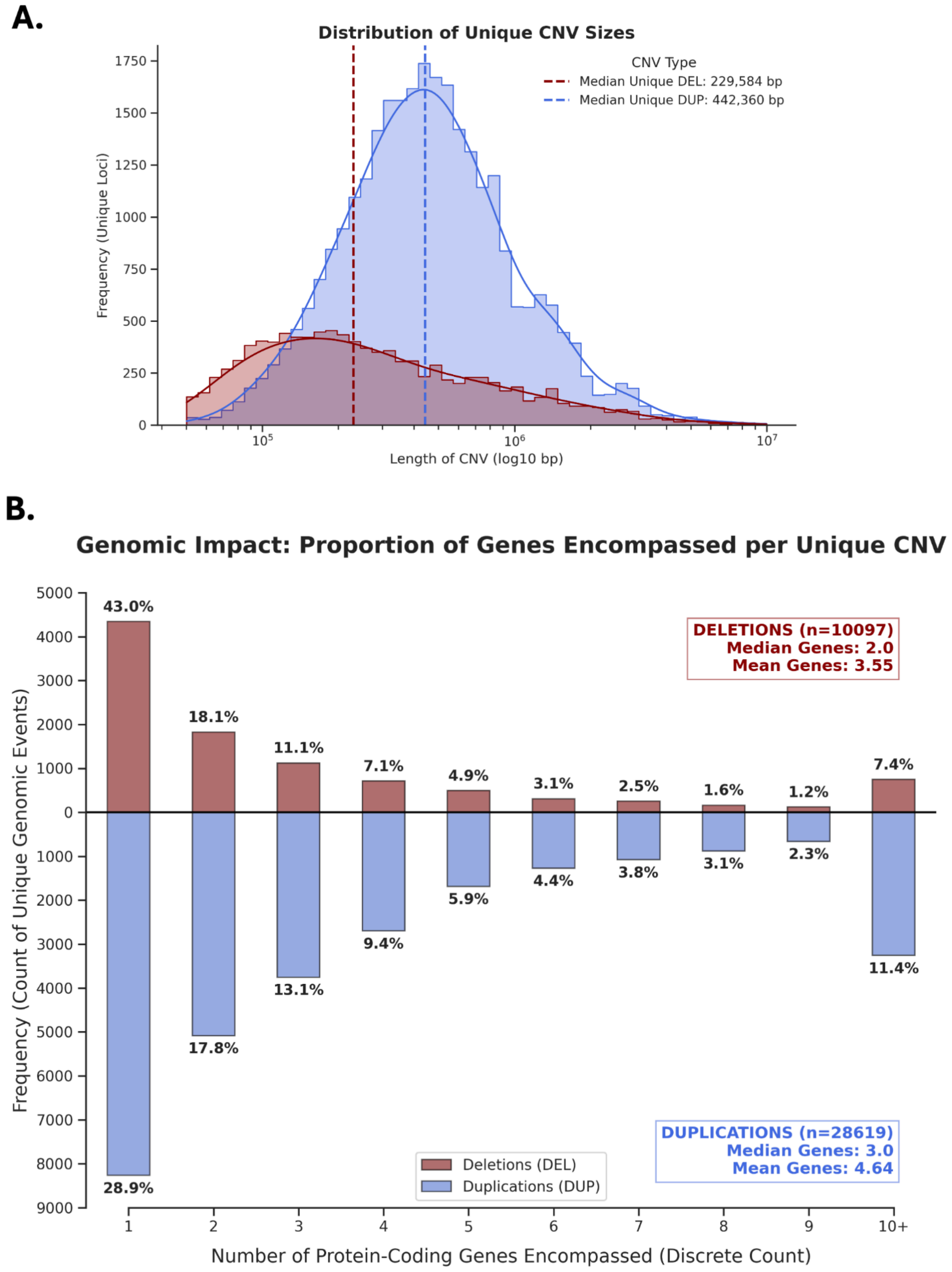


**Supplementary Figure 49: Genomic characteristics and gene content of unique deletions and duplications**

(A) Distribution of Unique CNV Sizes. Histogram and density plot showing the length distribution (log10 base pairs) for unique deletions (DEL, red) and duplications (DUP, blue). Vertical dashed lines indicate the median size for each CNV type: 229,584 bp for deletions and 442,360 bp for duplications. (B) Proportion of Genes Encompassed per Unique CNV. Mirror bar chart displaying the frequency and percentage of unique genomic events categorized by the number of genes (protein-coding) they encompass (ranging from 1 to 10+). Deletions (n = 10,097) show a median of 2.0 genes and a mean of 3.55 genes per event. Duplications (n = 28,619) show a higher gene density with a median of 3.0 genes and a mean of 4.64 genes per event.

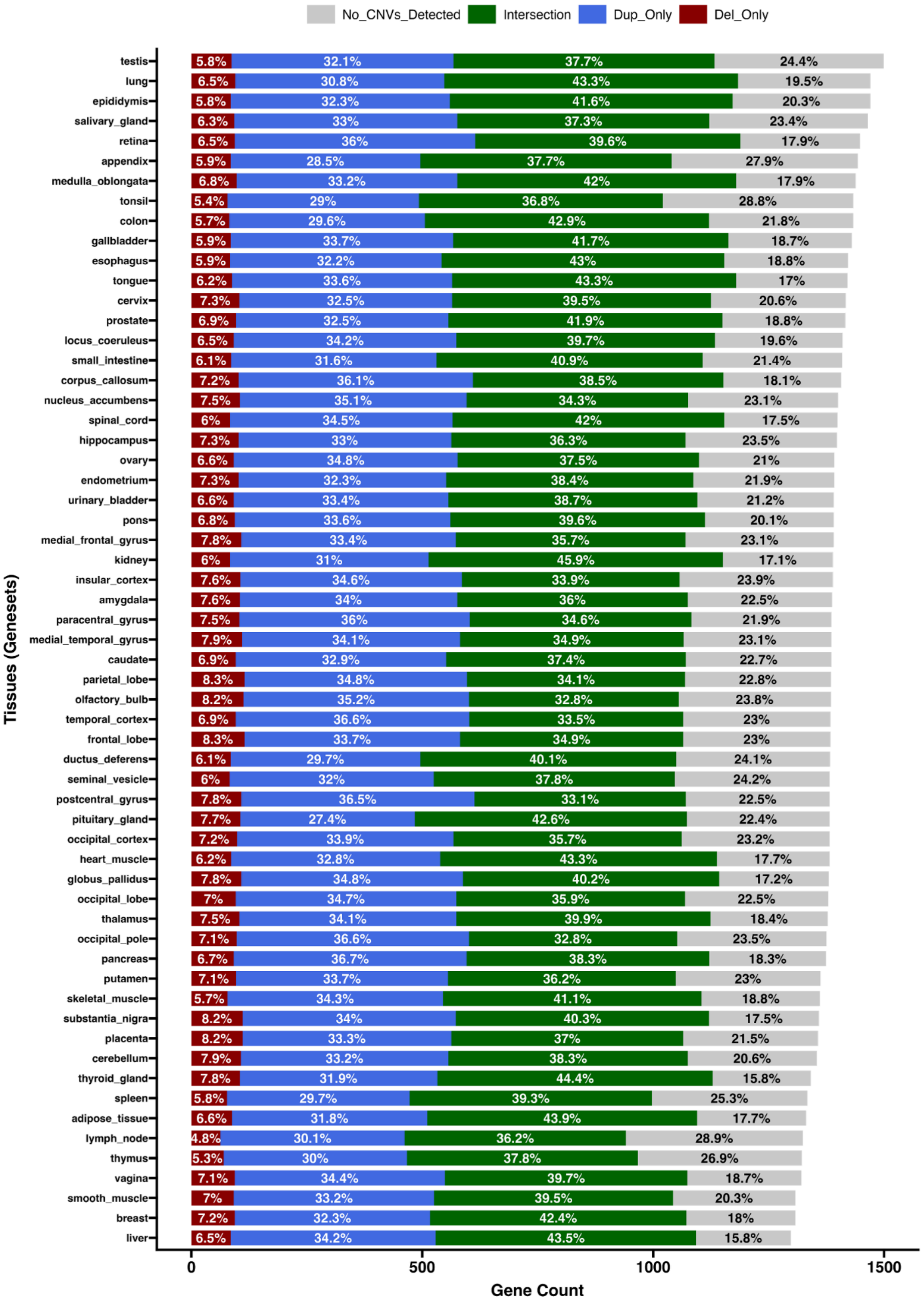


**Supplementary Figure 50: Proportion of genes in tissue-specific gene sets hit by observed CNVs (for BMI).**

Horizontal stacked bar chart depicting the intersection between 60 FANTOM5 tissue-specific gene sets and the CNVs observed in our dataset. Each bar represents the total gene count for a defined gene set, partitioned by the specific type of CNV hit: genes hit exclusively by observed deletions (Del_Only, red), genes hit exclusively by observed duplications (Dup_Only, blue), and genes hit by both CNV types (Intersection, green). The grey segment (No_CNVs_Detected) accounts for genes within each defined set that were not hit by any CNVs in our dataset. Internal labels indicate the percentage of the total gene count within each defined gene set that is hit by observed CNVs in each respective category.

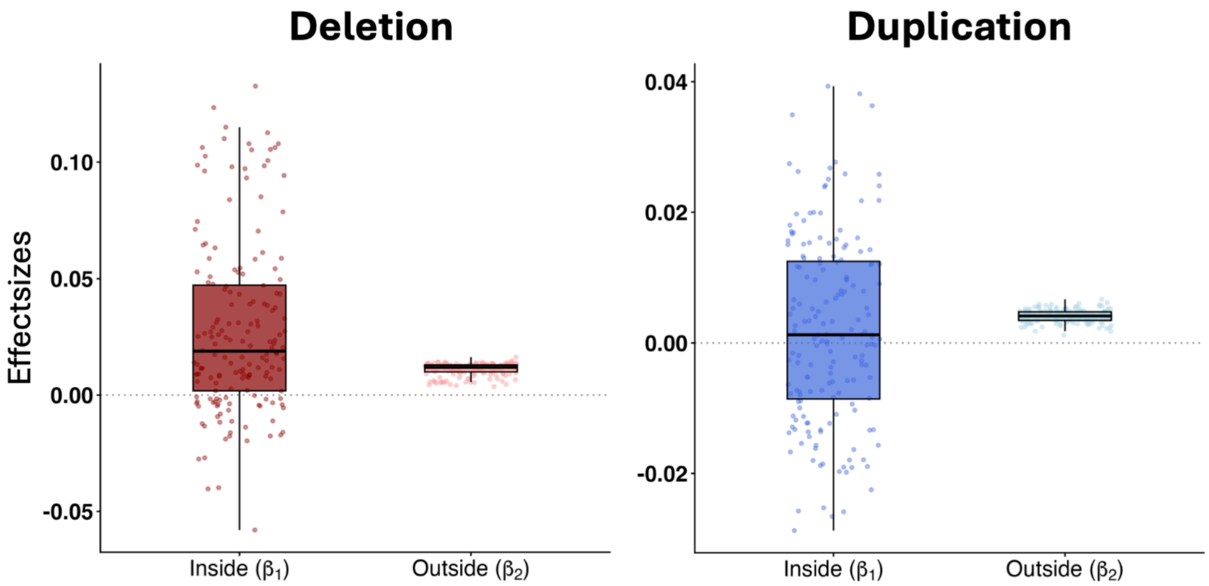


**Supplementary Figure 51: Comparison of burden effect sizes for gene set membership inside (β1) and outside (β2) for BMI**

Box plots comparing the effect size estimates for genes inside the target gene set (β1) versus genes outside the set (β2) across all 172 gene sets, shown separately for deletions (left) and duplications (right). Inside effects show greater variance than outside-set effects for both CNV types, consistent with gene-set-specific signal enrichment. The center line denotes the median, the box represents the interquartile range (IQR), and data points are shown as a jittered overlay to illustrate the full distribution of the gene sets' effect sizes. Full statistics for all traits are provided in **Table ST4**.

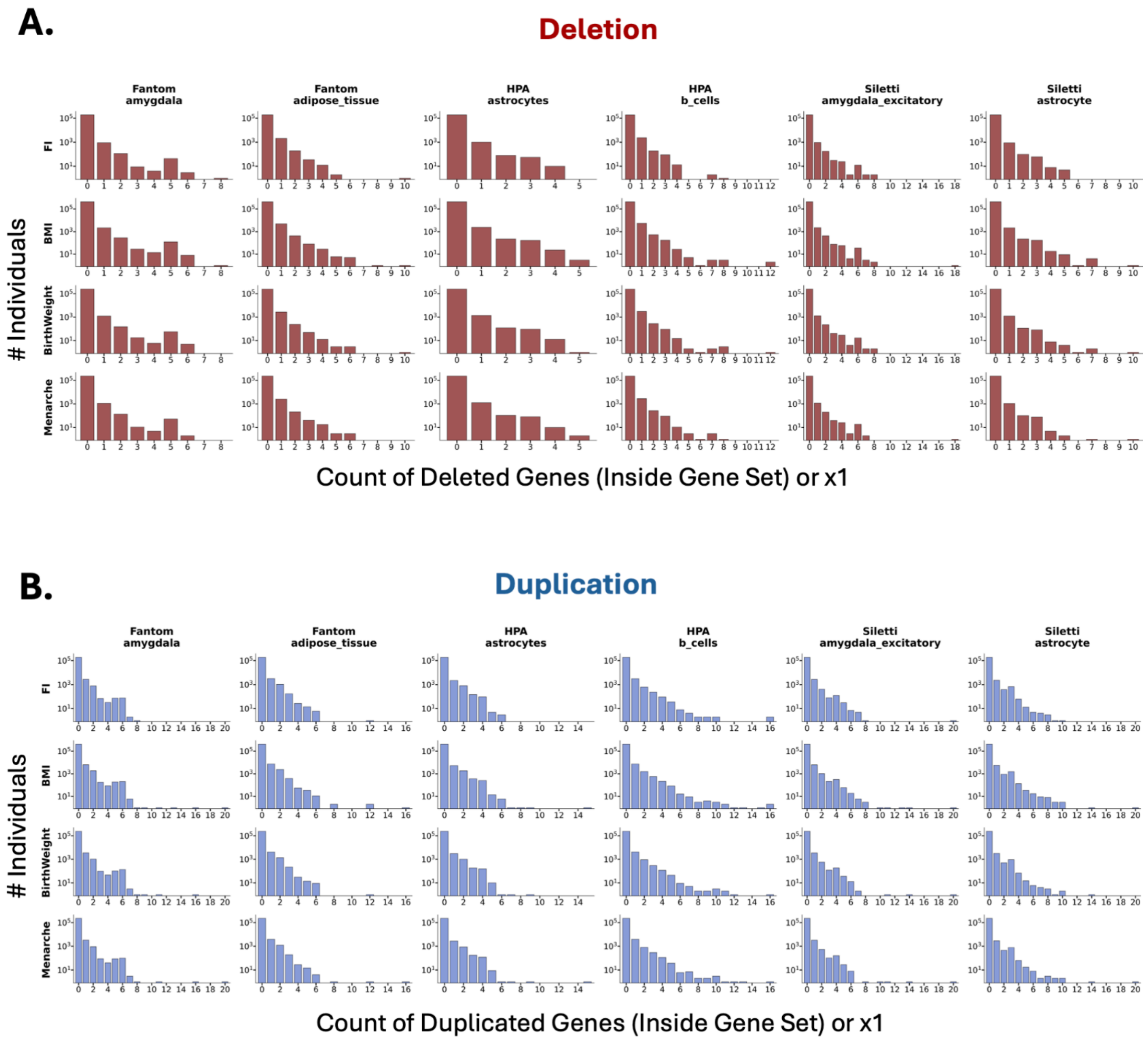


**Supplementary Figure 52: Distribution of the number of genes hit by CNVs per individual across representative gene sets and traits**

(A) Deletion Count Distributions. Grid of histograms showing the frequency of individuals (y-axis) categorized by the discrete count of deleted genes (x-axis; x1) within six representative gene sets across four traits. (B) Duplication Count Distributions. The observed distributions follow a Poisson-like decay, where the majority of individuals carry zero hits within a specific gene set, and a small fraction of the population carries multiple disrupted genes.

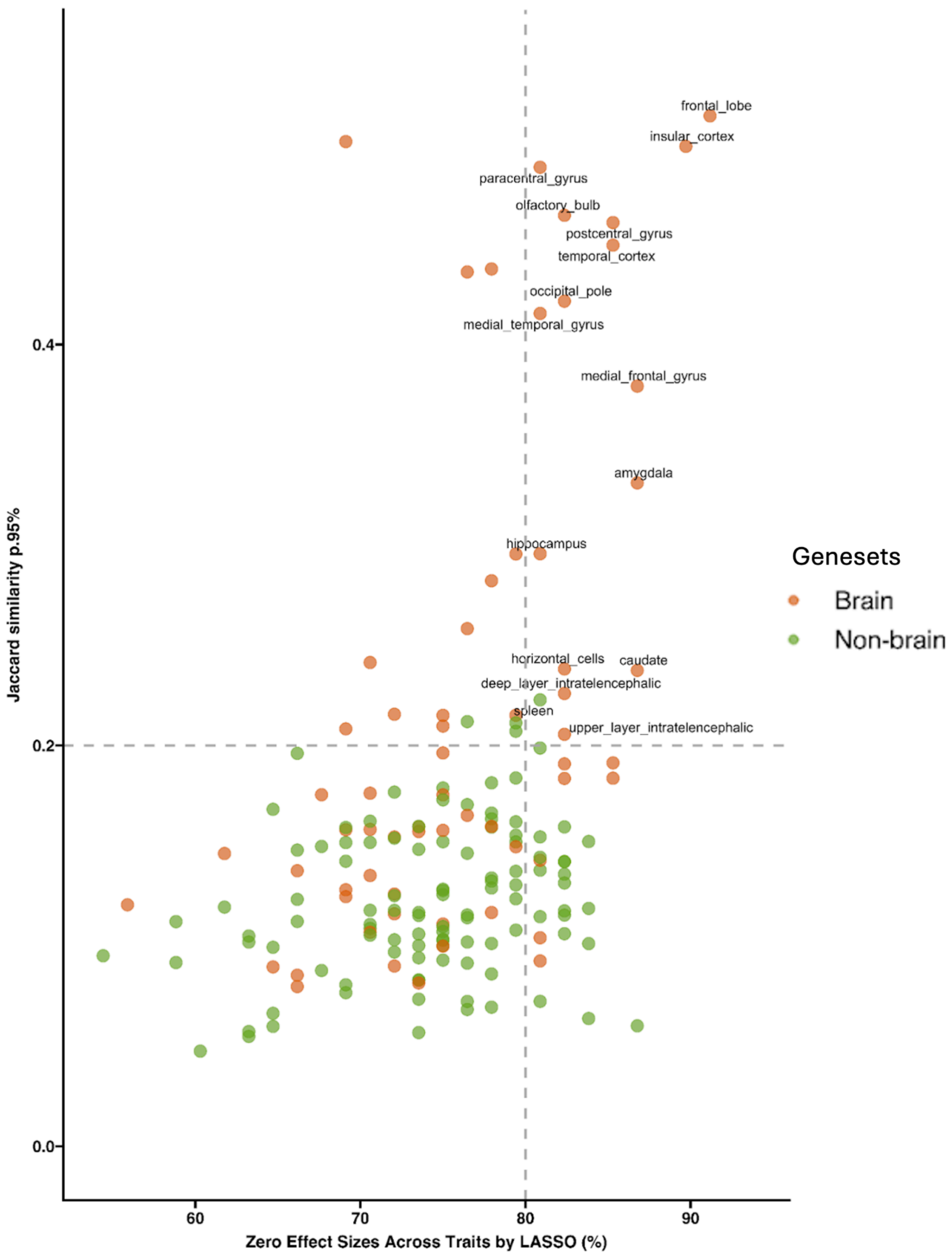


**Supplementary Figure 53: Scatterplot of geneset Jaccard similarity (95th percentile) vs geneset zero effect sizes across traits excluded by LASSO (%)**

Genesets with Jaccard similarity > 0.2 and > 80% zero effect sizes across traits were excluded. In total, 16 gene sets were removed from the genetic correlation analysis**.**

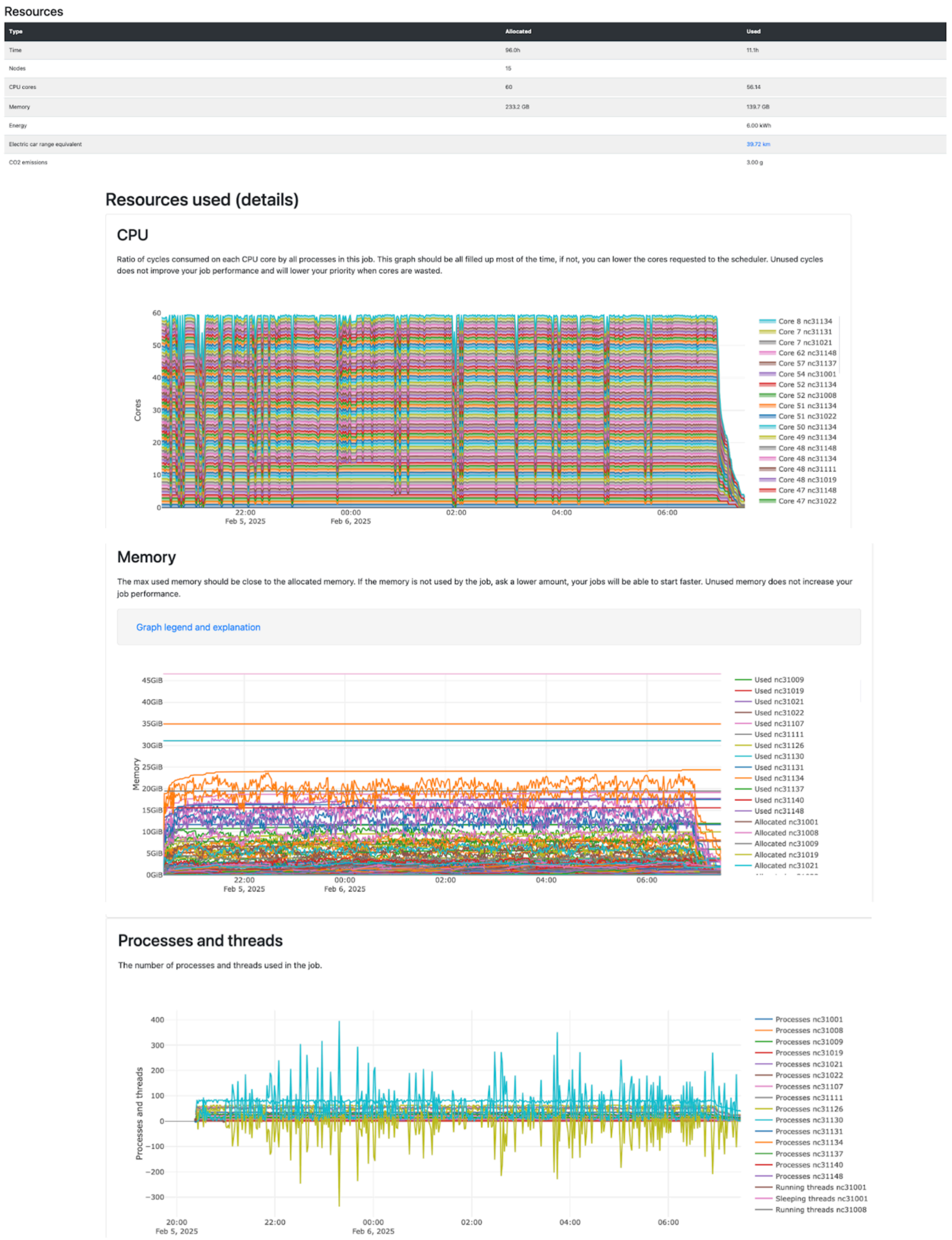


**Supplementary Figure 54: Performance of FunBurd running in parallel for constraint nulls genesets and traits (43,000 Jobs)**

This hybrid computation was executed in parallel across 1000 constraint-based null gene sets and 43 trait combinations. Within each run, the Funburd function utilized multithreading to evaluate 20 levels of constraint percentages ranging from 5% to 100%. In total, the computation involved 43 traits × 1000 constraint nulls × 20 constraint levels × 2 CNV types (deletions and duplications), resulting in 1,720,000 individual Funburd associations.

*Statistics of the performance of hybrid jobs derived from Compute Canada.*
